# Data-driven cardiometabolic phenogroups reveal distinct subclinical cardiac and proteomic profiles

**DOI:** 10.64898/2026.06.15.26355645

**Authors:** Jiayu Feng, Jocelyn Han Shi Chew, Hangkuan Liu, Liang Zhao, Nadja Mikulic, Abhinit Kumar Ambastha, Melvin Khee Shing Leow, Derek John Hausenloy, Mark Dhinesh Muthiah, Wenru Wang, Michael Wei Liang Chee, Ju Lynn Ong, Calvin Woon Loong Chin, Dianbo Liu, Weiqiang Seow, Reshma Aziz Merchant, Xin Zhou, Qing Yang, Josip Car, Lieng Hsi Ling, Roger Foo, the RESET Consortium

## Abstract

**Background:** Cardiometabolic disease is heterogeneous and incompletely resolved by conventional classification. We aimed to use expanded variables to identify data-driven phenogroups and characterise their echocardiographic and proteomic features.

**Methods:** Latent class analysis was applied to a discovery cohort (RESET; n=1,034) using fourteen cardiometabolic variables. A decision tree model was constructed using the most important variables to enable practical phenogroup classification and facilitate external validation. External validation used three cohorts: PICMAN (n = 120, proteomics), the UK Biobank (n = 344,817, proteomics and outcomes) and CHARLS (n = 12,145, outcomes).

**Results:** Five latent phenogroups were identified: Metabolically Preserved (lowest burden of adiposity, insulin resistance and hyperglycemia), with and without hypertension (each n=244; 23.6%); Lean-Insulin Resistant (IR) (n=140; 13.5%); Obese-Insulin Sensitive (IS) (n=211; 20.4%); and Obese-IR (n=195; 18.9%). Although Lean-IR and Obese-IS showed discordant adiposity and insulin/glycemic status, both exhibited greater subclinical diastolic dysfunction (36.7% and 38.8%, respectively) than the Metabolically Preserved group (16.5%; P<0.001), with impaired global longitudinal strain more prominent in Obese-IS (22.5% vs 13.6%; P<0.001). A decision tree model nominated four variables as most discriminative for the phenogroups (visceral adiposity, IR, elevated SBP, and HbA1c), classifying individuals with an AUC of 0.84 (0.81-0.86), and was used for external validation. Validation in the PICMAN cohort using plasma proteomes showed signatures for the two discordant phenotypes: an inflammatory emphasis in Lean-IR (IL-6, PLCB2) and a stronger hepatic metabolic/injury signature in Obese-IS (ADH4, KRT8/KRT18), upon a shared insulin–endocrine core in both (LEP, IGFBP1, FGF21). In the UK Biobank, phenogroups assigned using surrogates for visceral adiposity and IR (waist circumference and TG:HDL-C ratio) replicated the proteomic signature, and were associated with incident myocardial infarction and stroke. Hazard ratios for the composite outcome after adjusting for age and sex were 1.86 (1.75-1.98) for Lean-IR, 1.85 (1.75-1.97) for Obese-IS, and 2.75 (2.56-2.95) for Obese-IR, compared with the Metabolically Preserved group.

**Conclusion:** Data-driven classification with expanded variables reveals underlying heterogeneity. Phenogroups with discordant adiposity and insulin/glycemic status are associated with subclinical cardiac dysfunction, but differ in their proteomic pathways. The inferred divergent biology implies that individuals of different phenogroups may benefit from differing targeted prevention.

**Condensed Abstract:** Cardiometabolic disease is heterogeneous and incompletely resolved by conventional classification. Using latent class analysis, we identified five phenogroups, including two discordant phenotypes: lean insulin-resistant (Lean-IR) and obese insulin-sensitive (Obese-IS). Despite opposite adiposity, both had a higher prevalence of subclinical cardiac dysfunction than metabolically preserved individuals. Plasma proteomics, validated in the PICMAN cohort, showed an inflammatory signature in Lean-IR and a stronger hepatic metabolic/injury signature in Obese-IS, on a shared insulin–endocrine core. In the UK Biobank, both phenotypes carried a similarly elevated CVD risk, showing that this convergent risk can arise from divergent biology, with implications for precision prevention.

**Graphic Abstract:** BMI, body mass index; WC, Waist circumference; TG, Triglyceride; LDL-C, Low density lipoprotein cholesterol; HDL-C, High density lipoprotein cholesterol; FPG, Fasting plasma glucose; HbA1c, Glycated Hemoglobin A1c; HOMA-IR, Homeostatic Model Assessment of Insulin Resistance; SBP, systolic blood pressure; DBP, diastolic blood pressure; DD, Diastolic dysfunction; GLS, Global longitudinal strain; MI, myocardial infarction

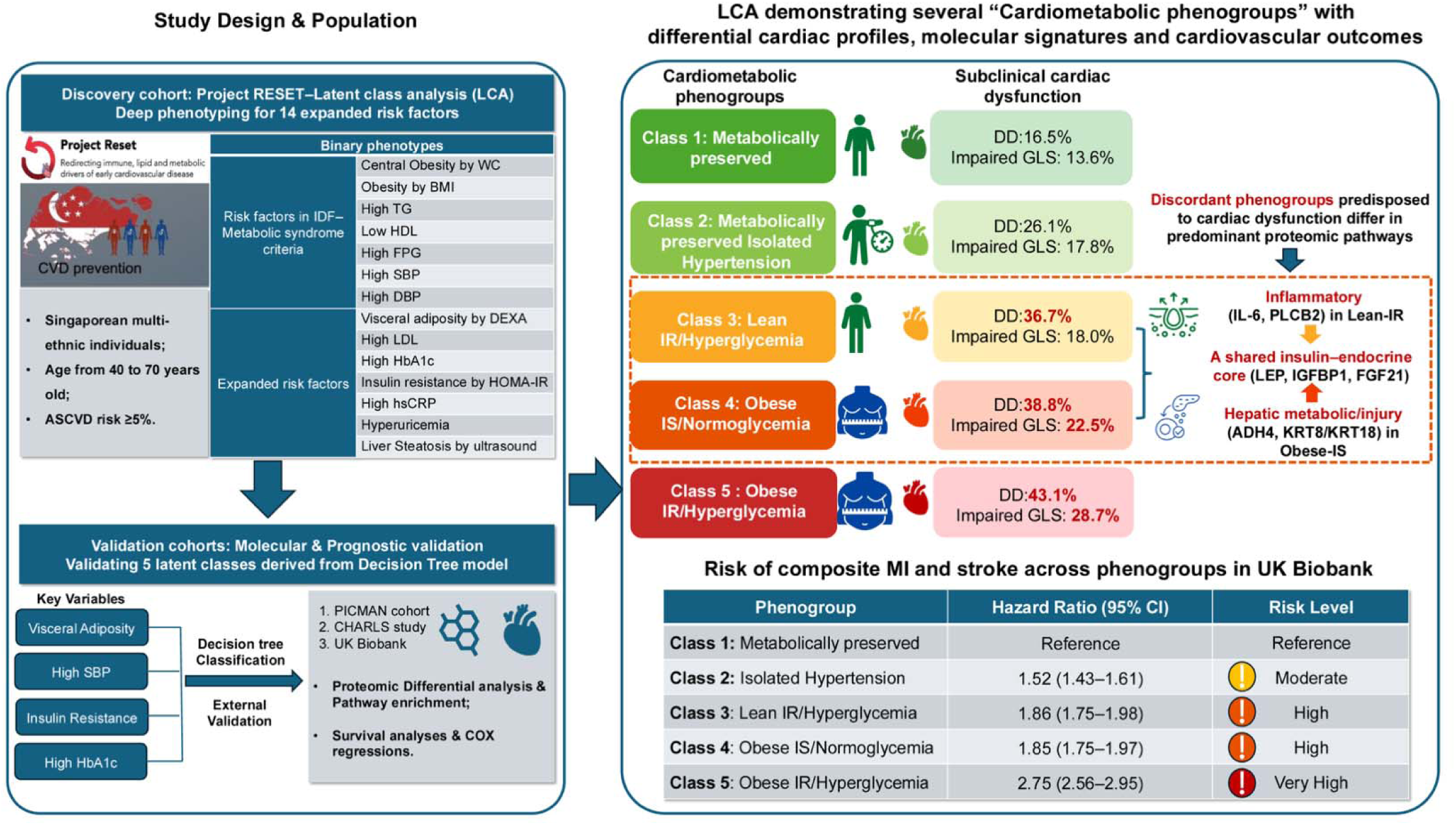

## INTRODUCTION

Cardiovascular disease (CVD) represents a growing global health burden, with metabolic dysfunction increasingly recognised as a key driver of adverse outcomes.^1^ The current framework, evolving from metabolic syndrome (MetS)^2,3^ to the recently proposed Cardiovascular–Kidney–Metabolic (CKM) syndrome,^4^ provides clinically practical, stage-based categorisation. Yet individuals within the same category can differ markedly in underlying biology and subclinical organ dysfunction, as seen in the discordance between adiposity and metabolic status (e.g., metabolically healthy obesity [MHO], metabolically unhealthy normal weight [MUNW]).^5^ Genetic analyses further indicate that adiposity and cardiometabolic risk are at least partially uncoupled.^6^ Resolving this cardiometabolic heterogeneity requires approaches that do not presuppose known categories. Unsupervised clustering has recently been applied at scale to cardiometabolic phenotyping: data-driven analysis identified body mass index (BMI)-discordant profiles whose cardiometabolic risk diverged from that expected for their BMI, with divergent risks of major adverse cardiovascular events. ^7^

While BMI and waist circumference (WC) are foundational screening tools, they are not enough to quantify ectopic fat, identify early impairment in adipose tissue expandability, or capture coordinated deterioration across hepatic, adipose, and cardiometabolic pathways.^8^ “Silent” signs of metabolic dysfunction such as insulin resistance (IR), hepatic fat accumulation, visceral adiposity, and systemic inflammation,^9,10^ are not detected using conventional clinical screening tools, and yet may independently increase the cardiovascular risk.^11,12^ These parameters were also not incorporated in previous unsupervised efforts to define adiposity–metabolic health discordance, which relied largely on BMI and routine biomarkers.^7^ Additional parameters for visceral adiposity and IR should therefore facilitate more precise identification of latent cardiometabolic phenotypes.^13^ Critically, metabolic dysfunction manifests in the heart before overt disease. Subclinical cardiac dysfunction, such as impaired left ventricular global longitudinal strain (GLS) or diastolic dysfunction with preserved ejection fraction links metabolic derangement to clinical cardiovascular events^14–16^. However, this remains largely unexplored across the data-driven unsupervised cardiometabolic phenotypes, as does the systematic proteomic characterisation which may reveal distinct biological signatures.

In this study, we undertook a data-driven approach to identify latent cardiometabolic phenogroups using expanded variables, and evaluated their subclinical cardiac dysfunction by echocardiographic features in the discovery cohort. We then derived a parsimonious classifier from the most discriminative variables and validated the phenogroups in three independent cohorts, characterising their plasma proteomic signatures and long-term cardiovascular risk. We focused particularly on discordant phenotypes, testing whether comparable cardiovascular risk can arise through distinct biological and cardiac pathways.

## METHODS

### Study populations

We used data from 4 different cohorts. (1) A discovery cohort was formed from individuals (n=1,034) recruited thus far in RESET (Redirecting immune, lipid, and metabolic drivers of early cardiovascular disease), an ongoing study of multi-ethnic Singaporeans (Malay, Indian and Chinese) aged 40 to 70 years old with a 10-year Atherosclerotic Cardiovascular Disease (ASCVD) risk of ≥ 5% without prior major adverse cardiovascular events (MACE).^17^ (2) An external validation cohort study PICMAN (Platform for the interdisciplinary study of cardiovascular, metabolic and neurovascular diseases, n=120), which recruited individuals participants aged 21–72 years with a 10-year ASCVD risk ≥ 5% but no established CVD and type 2 diabetes, ^18^ which was used to validate the proteomic signature across phenogroups. Two publicly available longitudinal datasets were used for external prognostic validation, UK Biobank and The China Health and Retirement Longitudinal Study (CHARLS). (3) In the UK Biobank, individuals of self-reported White ethnicity and aged 40–69 years were included. Participants with prevalent myocardial infarction (MI), stroke, cancer, or missing follow-up data and cardiometabolic metrics were excluded. (4) In CHARLS, individuals with prevalent CVD, stroke, or cancer, as well as those outside the target age range (40–70 years) or missing data at baseline were excluded. For every individual, the baseline visit was defined as the first survey wave at which all cardiometabolic variables were simultaneously available and not missing. The flowcharts for inclusion and exclusion are shown in **Figure S1**.

The discovery cohort–RESET was approved by the Ethics Committee of the National University Health System on September 2023, and is registered at clinicaltrials.gov (NCT06211868). The validation cohort–PICMAN was approved by the National Healthcare Group Domain Specific Review Board (2021/00003). The validation cohort–UK Biobank access was approved under project number: 94477. This cohort study was reported in accordance with the STROBE statement.

### Measurements

In the RESET cohort, 14 binary indicators aligned with clinical severity thresholds were used for phenotyping, incorporating 7 conventional MetS measures^2^: obesity defined by BMI, central obesity (waist circumference), elevated triglycerides, SBP, DBP, fasting glucose and reduced HDL cholesterol, and 7 additional expanded metabolic risk factors: elevated LDL-C, HbA1c, hsCRP, hyperuricemia, IR defined by HOMA-IR, visceral adiposity defined by Dual-Energy X-ray Absorptiometry [DEXA] – visceral adiposity tissue area [VAT cm^2^], and liver steatosis defined by liver ultrasound – Controlled Attenuation Parameter [CAP db/m]). Thresholds used to dichotomize the clinical indicators and MetS definition are detailed **Supplementary Methods 1**. Participants were classified into Cardiovascular-Kidney-Metabolic Syndrome (CKM) stages according to 2026 ACC guideline.^19^ Echocardiography was performed in the discovery cohort, global longitudinal strain (GLS) was measured by two-dimensional speckle-tracking. Impaired GLS was defined as |GLS| < 18% (borderline or abnormal GLS).^20^ Left ventricular diastolic function was graded (normal, grade 1–3) according to the 2025 ASE/EACVI recommendations.^21^

In the PICMAN study, considering the sample size limitation and difference in VAT measurements, IR was defined as HOMA-IR≥66.6th percentile, and visceral adiposity was defined as VAT volume (cm^3^) by MRI≥ 66.6th percentile. Plasma proteomic profiling was performed using the Olink Explore HT platform (5,416 proteins, details in **Supplementary Methods 2**).

In the UK Biobank, IR was replaced by the surrogate indicator high TG:HDL ratio (≥3.5 mmol/L in male and 2.5 mmol/L in female),^22^ and visceral adiposity was replaced by the surrogate indicator central obesity (WC≥102cm in male and ≥88cm in female)^2^. Plasma proteomes were profiled using Olink (2,923 proteins) as a part of the UK Biobank Pharma Proteomics Project (UKB-PPP).^23^ Similarly, in CHARLS, IR was replaced by the surrogate indicator high TG:HDL ratio (≥1.51 mmol/L in male and 0.84 mmol/L in female),^22^ and visceral adiposity was replaced by the surrogate indicator central obesity (WC≥90cm in male and ≥80cm in female),^2^ as Asian-specific thresholds.

### Outcomes

CHARLS and the UK Biobank were used for prognostic validation in Chinese and white population, respectively. In CHARLS, incident CVD or stroke were defined as the first self-reported physician diagnosis occurring after baseline, consistent with previous CHARLS-based studies.^24^ In the UK Biobank, incident MI or stroke were defined by ICD-10 codes.

### Cardiometabolic phenotyping in the discovery cohort

We implemented latent class analysis (LCA) to define cardiometabolic phenogroups, using a model-based classification method for categorical or binary indicators by providing probabilistic class membership. To evaluate the local independence assumption, residual correlations between pairs of indicators were calculated. To determine the optimal number of latent classes, models specifying 2 to 6 classes were fitted and evaluated using the Bayesian Information Criterion (BIC). The model with minimum BIC was selected as the optimal solution (**Supplementary Method 3 and Table S1**). LCA was conducted using R package *poLCA.*^25^ The conditional probabilities (P[phenotype | class]) of each binary-defined metabolic phenotype across the latent classes were shown by R package *ComplexHeatmap*.

Sensitivity analyses were conducted to assess the performance, stability and reproducibility of the LCA model: (1) a sub-group analysis stratified by gender, (2) a LCA model after excluding highly correlated variables (residual correlation |r| > 0.10, **Table S2**), (3) a simple LCA model was fitted by only MetS variables, plus BMI derived obesity, (4) another LCA model with the number of latent classes equal to optimal number-1, (5) varying each threshold by ±5% to re-fit the LCA models, (6) Latent profile analysis (LPA) and Hierarchical Clustering on Principal Components (HCPC) using continuous data.

### Development of supervised model for identifying latent classes and external validation

To facilitate clinical translation of the LCA-derived phenotypes, a supervised decision tree classifier was constructed to approximate latent class membership using a parsimonious set of 14 variables which were used for LCA in the discovery cohort. This similar strategy of distilling data-driven clusters into a parsimonious supervised classifier for external portability has been used in prior unsupervised clustering studies.^26,27^ The model was implemented using the *rpart* package in R with latent class membership as the outcome and selected cardiometabolic indicators as predictors. Tree complexity was constrained to improve generalizability and clinical interpretability by limiting the maximum depth to three levels. For sensitivity analysis, a multinomial logistic classifier was used to rank feature importance via SHAP (SHapley Additive exPlanations) values to test the consistency of top driving variables between supervised decision tree classifier and the multinomial model. Latent classes identified by supervised decision tree model were externally validated in PICMAN, CHARLS and the UK Biobank, following which differential molecular profiles and cardiovascular outcomes were analysed.

### Statistical analysis

Sociodemographic and clinical characteristics were summarized using count (%) for categorical variables, and mean (Standard deviation: SD) for continuous variables. Continuous variables were compared across classes using analysis of variance (ANOVA). Categorical variables were compared using the chi-square test. Differential expression analyses for proteomes were conducted using R package *limma* across decision tree identified latent classes. Latent classes were annotated with putative biological modules according to top differential proteins and the dominant pathways enriched within each class (details in **Supplementary Method 4**). Protein-disease Mendelian randomization estimates for the signature proteins were retrieved from the publicly available proteome-phenome-atlas (https://proteome-phenome-atlas.com).^28^ Kaplan– Meier survival analyses were performed to estimate the cumulative incidence of outcomes. Cox proportional hazards regression models were applied to evaluate the association between decision tree identified latent classes and incident outcomes, with Class 1 specified as the reference group. Hazard ratios (HRs) and 95% confidence intervals (CIs) were estimated after adjustment for Model 1: age and sex; Model 2: age, sex, smoking status, WC, TG, HDL cholesterol, fasting glucose, HbA1c, anti-hypertensive, diabetic and dyslipidemic drugs. As several Model 2 covariates overlap with class-defining variables, Model 1 is considered the primary analysis, with Model 2 serving as a sensitivity analysis to assess the robustness of associations under full adjustment. All statistical tests were two-sided, and a p-value <0.05 was considered statistically significant.

## RESULTS

### Baseline characteristics in the discovery RESET cohort

A total of 1,034 participants from the discovery cohort–RESET were studied, with sociodemographic and clinical characteristics in **Table 1** and **Table S3-S7**. Mean (SD) age was 63 (5.3) years, and 703 participants (68.0%) were male. Most participants were of Chinese ethnicity (937 [91.0%]), followed by Indian (54 [5.2%]) and Malay (15 [1.5%]). Mean BMI was 24.9 kg/m², with 46.3% of participants classified as overweight and 19.1% obese. The prevalence of diabetes, hypertension and dyslipidemia was 17.5%, 65.7% and 66.5%, respectively. 24.7% of participants had MetS according to IDF criteria and 32.4% had MetS based on AHA/NHLBI statement (≥3 of 5 components). The proportion of participants classified into CKM stages 1, 2 and 3 was 8.5%, 47.6% and 43.9%, respectively. Mean DEXA-measured VAT area was 92.6 cm²; 37.3% had visceral adiposity defined by VAT ≥100 cm², 37.9% had liver steatosis defined by liver ultrasound CAP≥254 dB/m, and 36.4% showed IR defined by HOMA-IR≥2.5. Notably, among 790 (76.4%) participants with normal FPG (<5.6mmol/L), 25.7% had IR and 53.9% had high HbA1c (≥5.7%, **Table S8**), revealing metabolic abnormalities that conventional fasting-glucose screening overlooks. A correlation matrix of all continuous cardiometabolic traits are shown in **Figure S2.** For the echocardiographic characteristics, 68.9% of participants had normal diastolic function, 23.8% had Grade 1 and 7.2% had Grade 2 diastolic dysfunction; 19.9% had impaired GLS (|GLS| < 18%).

**Table 1.**
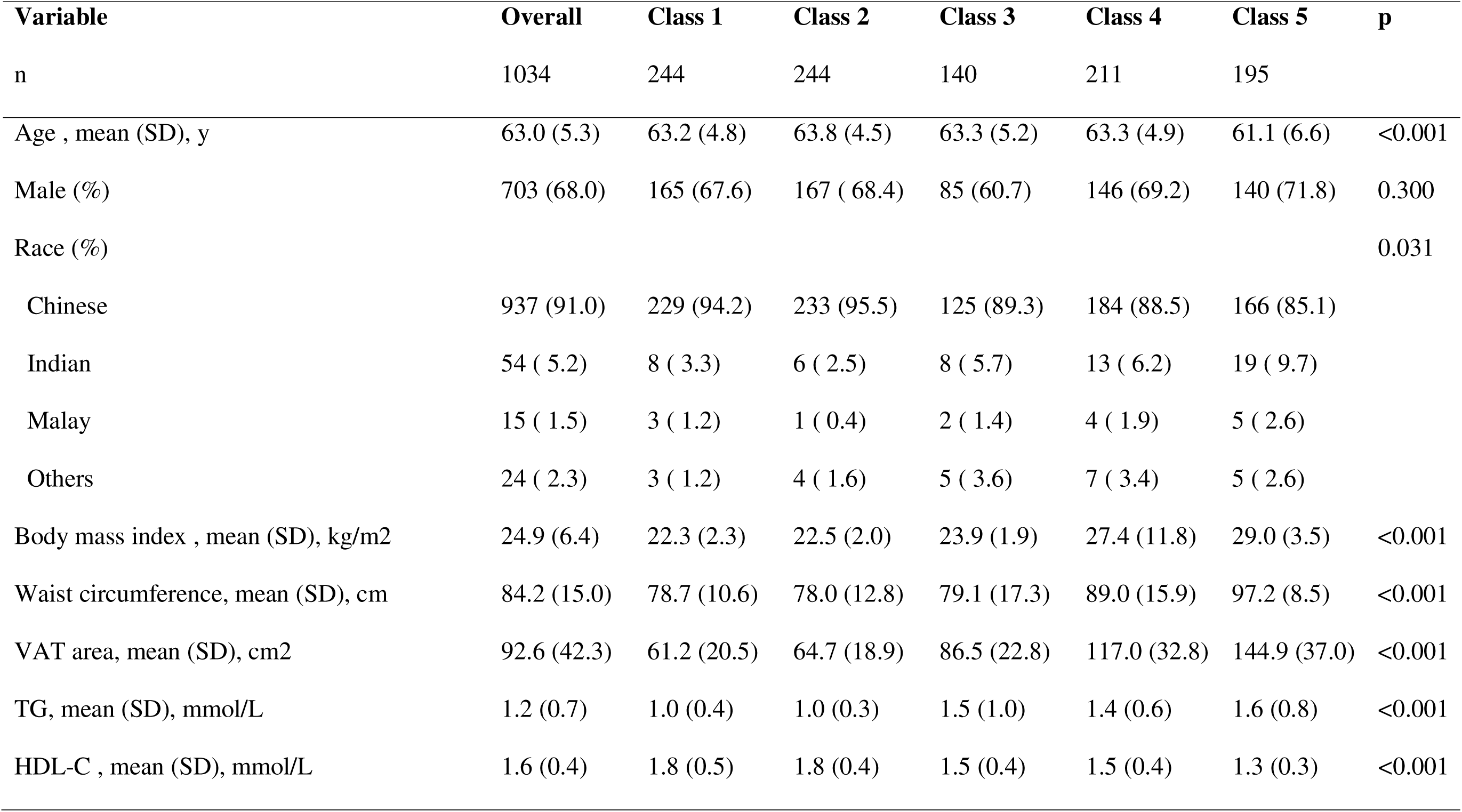
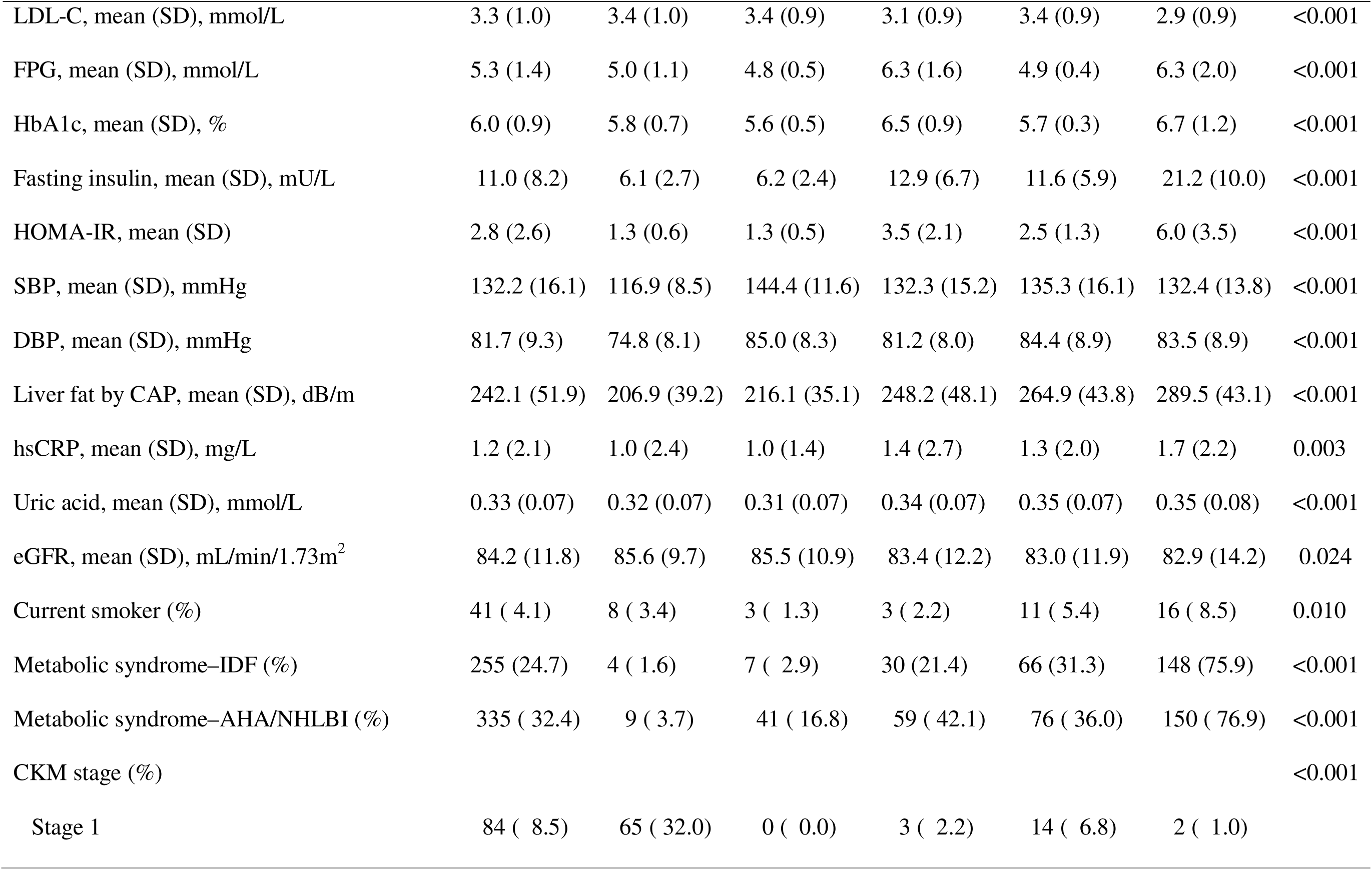
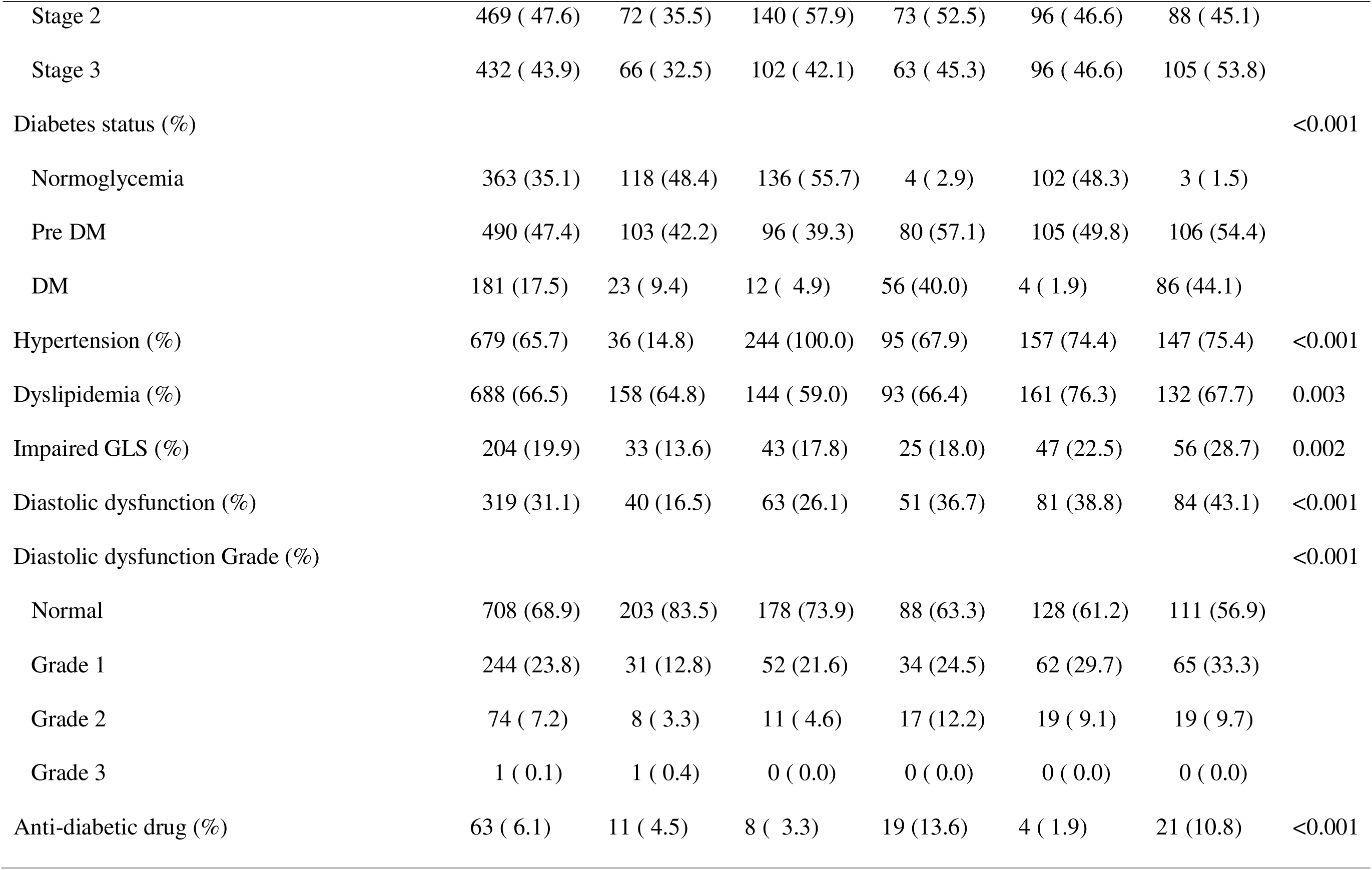
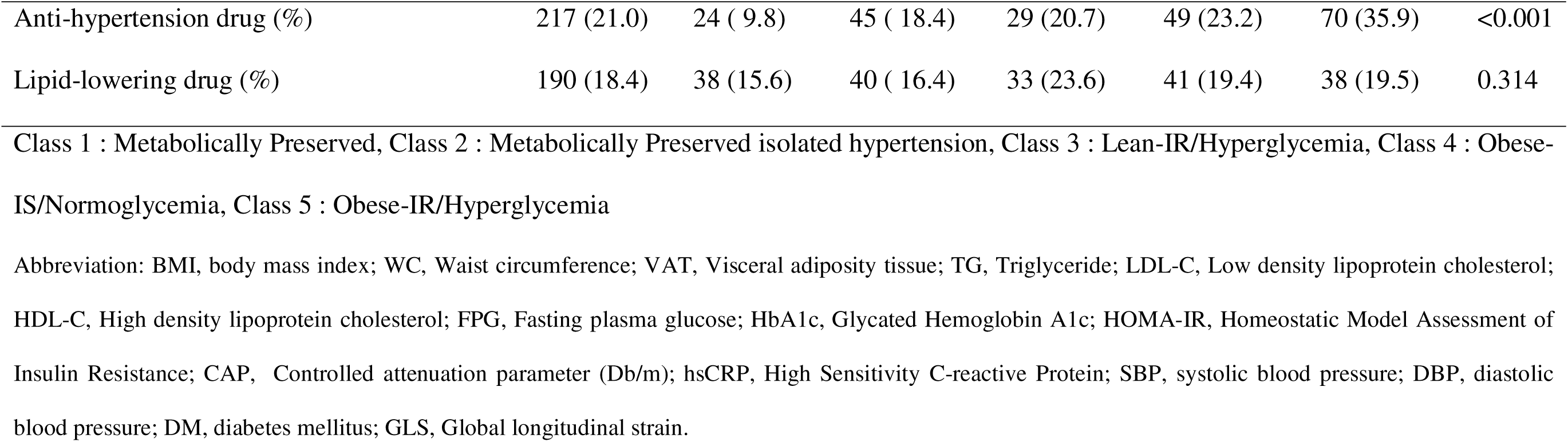
Characteristics according to 5 latent classes in discovery cohort–RESET study.

### Five latent phenotype classes in the discovery RESET cohort

Class 1 comprised individuals with a Metabolically Preserved phenotype (n=244; 23.6%). This group exhibited the lowest prevalence of metabolic multi-comorbidities, with low probability/prevalence of obesity/adiposity, IR, high FPG, high TG, hyperuricemia and high SBP/DBP (**Figure 1A-B**). Class 2 was also metabolically preserved, but had isolated hypertension (n=244; 23.6%). A distinguishing feature of this class was the highest probability of elevated SBP (100%), in the absence of other metabolic risk factors.

**Figure 1.**
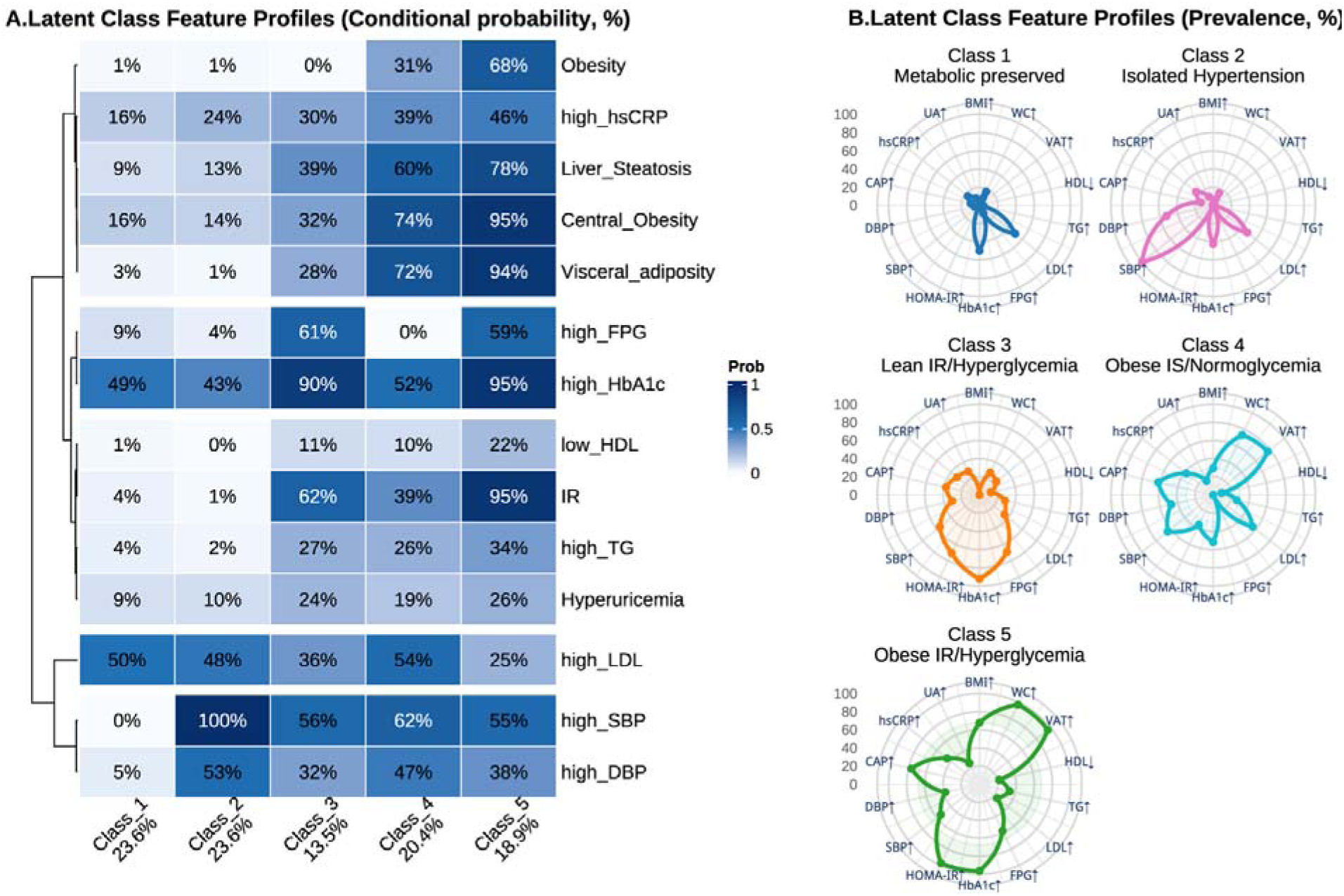
Phenotypes using expanded metabolic risk factors identified by latent class analysis (LCA) in discovery cohort RESET. **A.** Heatmap showing the conditional probabilities (P[phenotype | class]) of each binary-defined metabolic phenotype in the 5 latent classes identified by LCA. Class 1 : Metabolically Preserved, Class 2 : Metabolically Preserved isolated hypertension, Class 3 : Lean-IR/Hyperglycemia, Class 4 : Obese-IS/Normoglycemia, Class 5 : Obese-IR/Hyperglycemia. **B.** Radar plots illustrating the within-class prevalence of each metabolic risk factor in the 5 latent classes. LCA, latent class analysis; LDL, Low density lipoprotein cholesterol; HDL, High density lipoprotein cholesterol; SBP, systolic blood pressure; DBP, diastolic blood pressure; FPG, Fasting plasma glucose; HbA1c, Glycated Hemoglobin A1c; TG, triglyceride; HOMA-IR, Homeostatic Model Assessment of Insulin Resistance; CAP, Controlled attenuation parameter (Db/m); hsCRP, High Sensitivity C-reactive Protein; UA, Uric acid; WC, Waist circumference; BMI, Body mass index; VAT, Visceral adiposity tissue area by DEXA; IR, insulin resistance; IS, insulin sensitive.

**Figure 2.**
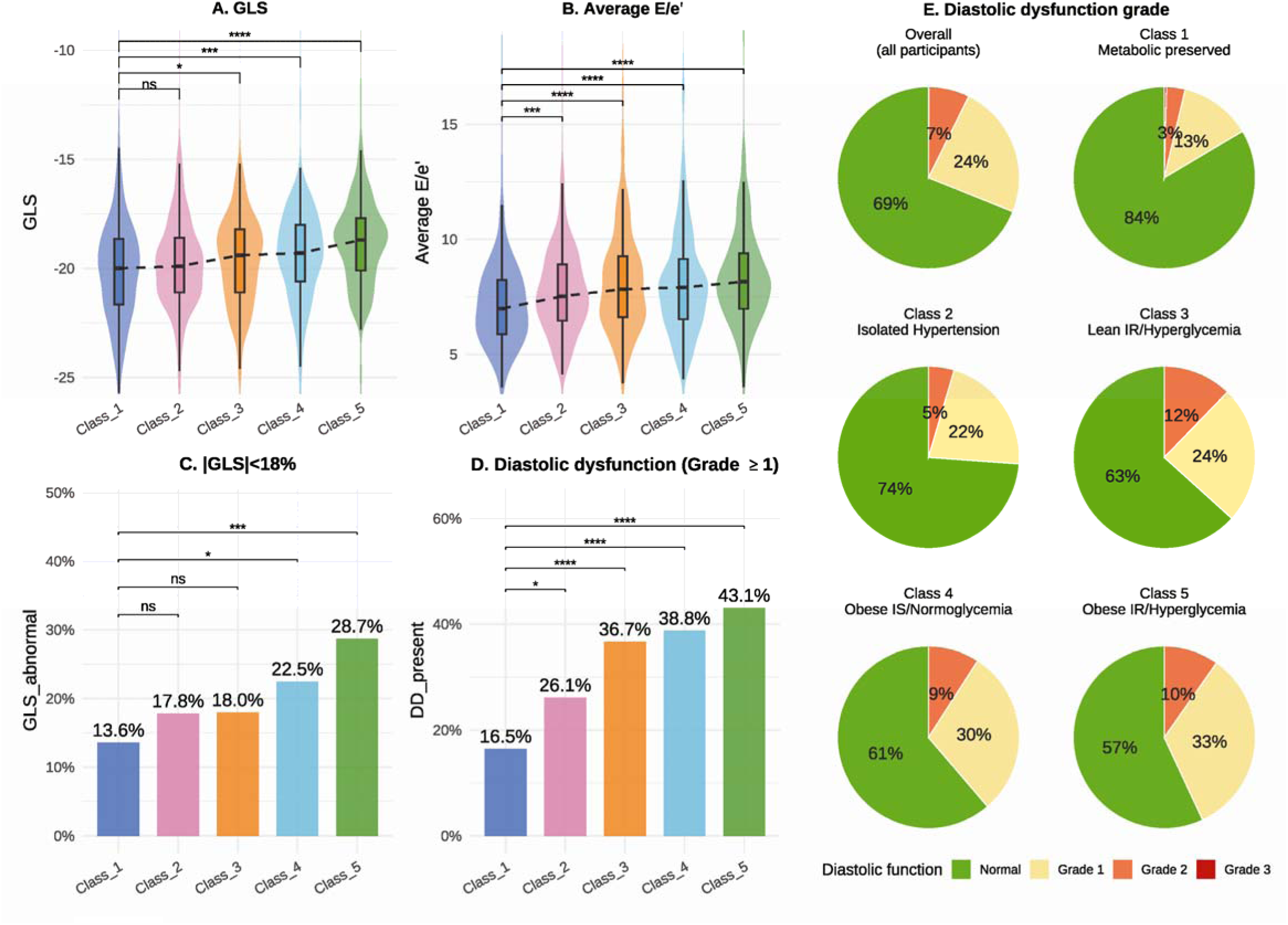
Subclinical cardiac dysfunction across latent classes in discovery cohort RESET. GLS, Global longitudinal strain; E/e_′_, ratio of early mitral inflow velocity to average early diastolic mitral annular velocity.

Class 3 (n=140; 13.5%) and Class 4 (n=211; 20.4%) represented intermediate cardiometabolic risk phenotypes, characterized by discordant adiposity and IR/hyperglycemia. Specifically, Class 3 was lean with IR and hyperglycemia, showing lowest conditional probability of BMI–obesity (0%, mean BMI 23.9 kg/m²) , lower probability of central obesity (32%) and visceral adiposity (28%, mean VAT 86.5 cm^2^), but highly elevated HbA1c (90%) and IR (62%). In contrast, Class 4 was obese but normoglycemic and relatively insulin sensitive (IS): central obesity (74%) and visceral adiposity (72%, mean VAT 117.0 cm^2^) were elevated compared to Classes 1, 2 and 3, although BMI–obesity was not dominant. In Class 3 and Class 4, MetS prevalence was only 21.4% and 31.3% by IDF criteria, and 42.1% and 36.0% by AHA/NHLBI criteria, respectively (**Table 1**), implying that conventional MetS criteria miss the majority of these discordant phenotypes.

Class 5 (n=195; 18.9%) displayed the highest risk burden across all metabolic markers, for BMI– obesity (68%), central obesity (95%), visceral adiposity (94%) and liver steatosis (78%), together with the highest rates of IR (95%) and high HbA1c (95%). 75.9% of individuals in this class satisfied the IDF MetS criteria, and 76.9% by the AHA/NHLBI MetS criteria. Nearly half (47.8%) of individuals labelled metabolically healthy obese (MHO) by BMI and conventional criteria (i.e. obese with fewer than two of elevated triglycerides, low HDL-C, elevated fasting glucose or elevated blood pressure) were in fact assigned to Class 5 (**Figure S3**), indicating that many MHO individuals are not truly healthy, carrying hidden IR and visceral fat. Although the latent classes were defined using categorical variables, their corresponding continuous metabolic profiles (**Figure S4**) showed concordant class-level differences, supporting the robustness of the phenotyping. The mean number of abnormal variables among the 14 risk factors was 5 in Class 3, 6 in Class 4 and 8 in Class 5, the highest of all classes (**Figure S5**).

We carried out a series of sensitivity analysis (**Figure S6-S11** and **Table S9-S12**) to validate our LCA approach by confirming that (1) the distribution of clinical indicators across all five latent classes were consistent when stratified by sex. (2) Applying the LCA model to simplified metabolic risk factors (variables in the MetS criteria) derived only 3 classes at best, and using fewer risk factors failed to recapitulate the 5 latent phenotypes that we had found. (3) Selecting 4 latent classes when using the 14 expanded metabolic risk factors also coherently revealed the phenotypes of Lean-IR/Hyperglycemia and Obese-IS/Normoglycemia. (4) After varying each binary threshold by ±5%, the overall feature profiles of latent classes remained highly consistent across analyses. (5) Continuous-variable analyses (LPA and HCPC) demonstrated the same concordant phenotype structures, with overlapping distributions of HOMA-IR between classes of lean with hyperglycemia, and obese with relatively normoglycemia.

### Subclinical cardiac dysfunction and CKM stages across latent classes in the discovery RESET cohort

Left ventricular ejection fraction (LVEF) was preserved across all five phenogroups (median 63.6–64.6%, p=0.265; LVEF<50% in only 0.4%), yet more sensitive indices deteriorated in a graded fashion: GLS was progressively less negative (median −20.0% in Class 1 to −18.7% in Class 5; p<0.001), average E/e′ ranged from 7.0 to 8.2 (**Table S5**), and the prevalence of diastolic dysfunction (≥grade 1) was more than doubled, from 16.5% in Class 1 to 43.1% in Class 5 (p<0.001, **Figure2**). Critically, the two discordant phenogroups carried a comparably elevated subclinical burden despite opposing adiposity. Lean-IR/Hyperglycaemia (Class 3), despite lower BMI and visceral fat, and Obese-IS/Normoglycaemia (Class 4) had near-identical diastolic dysfunction prevalence (36.7% and 38.8%), both higher than Class 1 (16.5%; p<0.001), although impaired GLS (|GLS|<18%) reached significance only in Class 4 (22.5% vs 13.6%, p<0.001), and not in Class 3 (18.0% vs 13.6%, p=0.213).

A subclinical cardiac phenotype is precisely what the CKM framework captures at Stage 3, which incorporates subclinical heart failure signalled by elevated NT-proBNP / hs-cTnI or by echocardiographic abnormalities. Consistent with their comparable cardiac dysfunction, Classes 3 and 4 fell into CKM Stage 3 at similar rates (45.3% and 46.6%; **Figure S12**). Although CKM Stage 1 was composed almost entirely of the Metabolically Preserved class (77.4%), Stages 2 and 3 pooled the discordant phenotypes together: Class 3 and Class 4 jointly accounted for 36.1% of Stage 2 (15.6% and 20.5%) and 36.8% of Stage 3 (14.6% and 22.2%, **Figure S12**).

### Supervised decision tree model for predicting unsupervised latent classes in discovery RESET cohort

To facilitate an implementation of LCA phenotyping for generalizable cardiometabolic risk-stratification, we constructed a decision tree model based on the most important node variables– visceral adiposity (by VAT area ≥100 cm^2^), high SBP (≥130mmHg), IR (by HOMA-IR≥2.5) and high HbA1c (≥ 5.7%, **Figure 3**). Overall predictive accuracy for the decision tree model in identifying unsupervised latent classes was assessed by area under the curve (AUC): 0.84, 95% CI:(0.81, 0.86) with 5-fold internal cross-validation. Other within-class performance metrics are shown in **Table S13**. In addition, multinomial model and SHAP also confirmed that high SBP, IR and visceral adiposity were the top 3 most important binary variables for supervised identification of latent classes (**Figure S13**).

**Figure 3.**
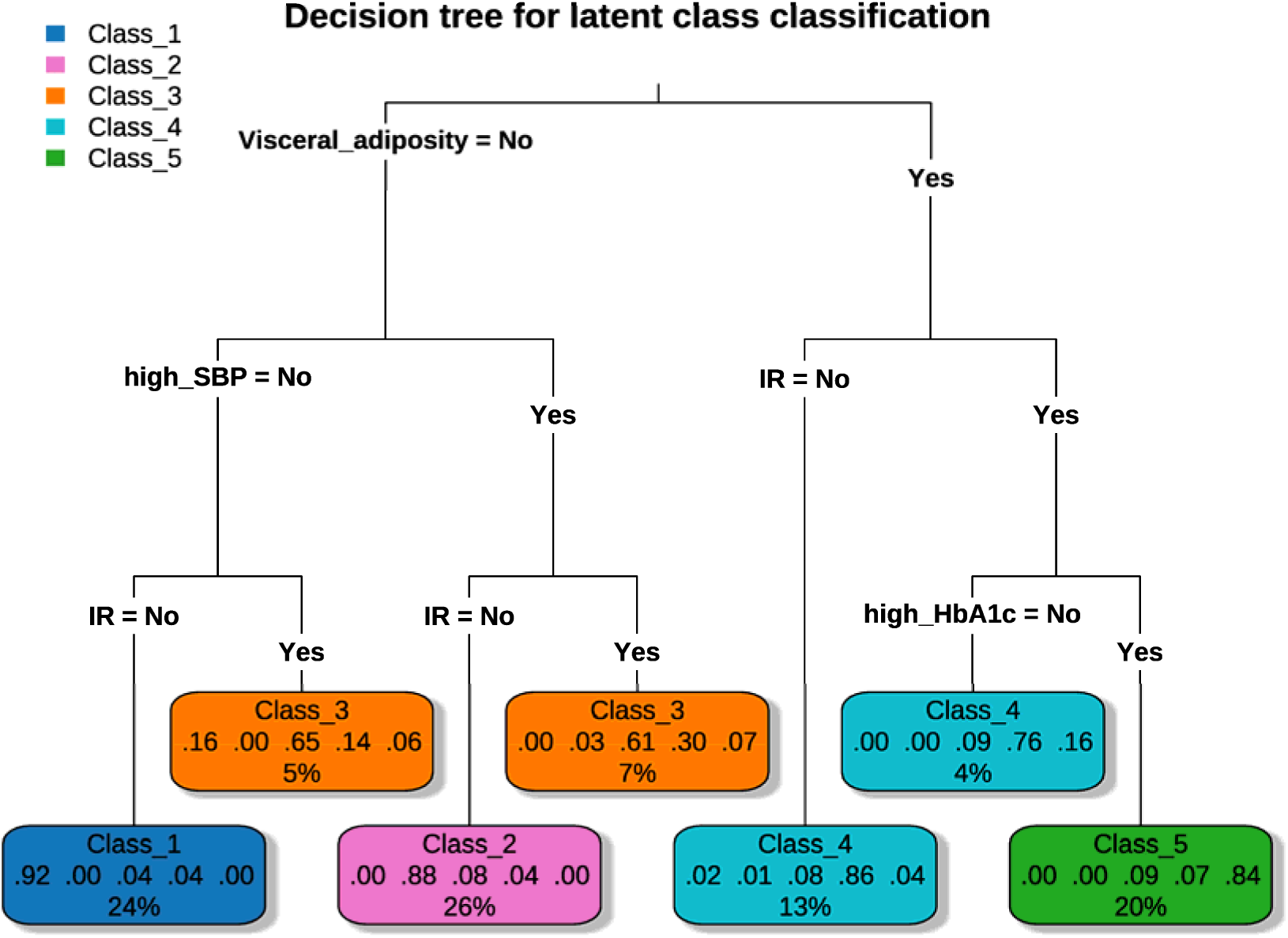
An interpretable decision tree model for supervised prediction of latent classes. The decision tree illustrating an interpretable classification model for assigning individuals to the 5 latent classes identified in Figure 1. Terminal nodes display the predicted class along with the class membership probabilities and the proportion of individuals classified into each leaf. Class 1 : Metabolically Preserved, Class 2 : Metabolically Preserved isolated hypertension, Class 3 : Lean-IR/Hyperglycemia, Class 4 : Obese-IS/Normoglycemia, Class 5 : Obese-IR/Hyperglycemia.

### Molecular characterisation of the phenotypic classes in the PICMAN cohort and the UK Biobank

Thereafter, we validated the decision tree–derived cardiometabolic classification in an independent cohort (PICMAN, n=120). Coherently, the 5 distinct classes with comparable distributions to those observed in the discovery cohort were validated (**Figure S14**). We analysed the plasma proteomes in PICMAN for differentially expressed proteins (DEPs). Importantly, 70.5% of DEPs identified from pairwise comparisons across latent classes in PICMAN were successfully validated in the UK Biobank (n=36,505 with proteomic data).

Class 2 with Metabolically Preserved isolated hypertension showed an activation of innate immune and inflammatory pathways, including NOD-like receptor signaling. Beyond this, the two discordant phenotypes (Classes 3 and 4) displayed predominantly distinct proteomic signatures. Biological annotation of the DEPs (|log fold-change| > 0.8 in PICMAN and replicated in the UK Biobank at FDR-adjusted P<0.05) is provided in **Table S14**. Specifically, Lean-IR/Hyperglycaemia (Class 3) was characterised by an inflammatory emphasis, represented by IL-6 and PLCB2 and an enrichment of cytokine and AGE–RAGE signalling, consistent with an endocrine–inflammatory metabolic-stress signature. In contrast, Obese-IS/Normoglycaemia (Class 4) showed a stronger and broader hepatic metabolic/injury signature: a core of hepatic enzymes (HAO1, GSTA1, ADH1B, FTCD) was abundant in both phenotypes but more markedly so in Obese-IS, including additional hepatic enzymes (ADH4, CA5A, DCXR) and hepatocyte-injury markers (KRT8, KRT18). Moreover, the enrichment of tyrosine and pyruvate metabolism KEGG pathways in Class 4 is further consistent with a greater hepatic metabolic load progressing toward hepatocyte injury (**Figure 4**).

**Figure 4.**
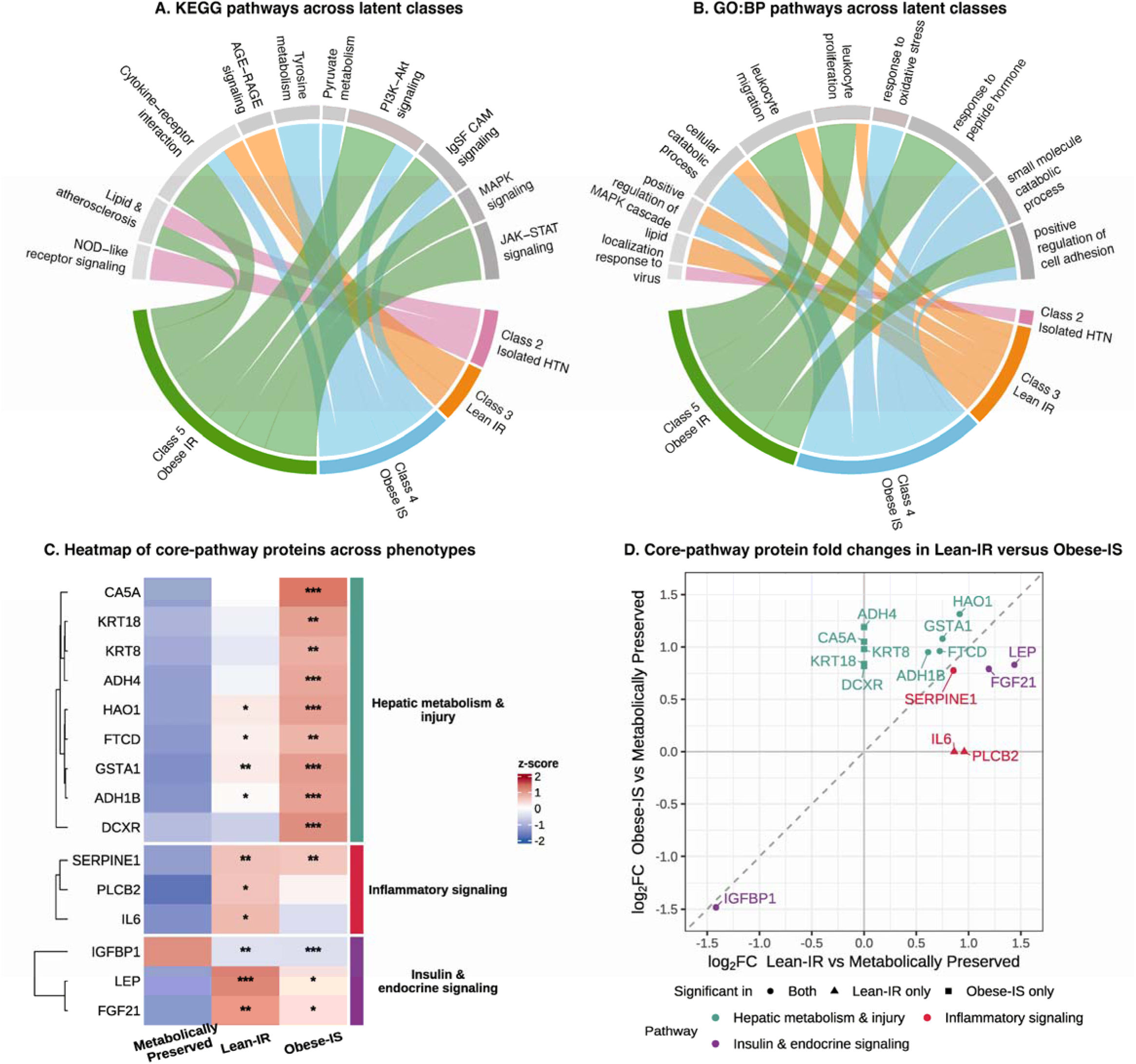
**Proteomic signatures of the latent classes A-B**: Top enriched KEGG and Gene Ontology (GO) pathways based on Differentially expressed proteins (DEPs) for Classes 2–5, compared with Class 1. **C**: Heatmap of z-scored protein abundance for core-pathway DEPs across the Metabolically Preserved (Class 1), Lean-IR/hyperglycemia (Class 3) and Obese-IS/normoglycemia (Class 4) groups. Core-pathway proteins meeting significance thresholds in PICMAN (P<0.05 and |log fold change| >0.8) and replicated in the UK Biobank (FDR-adjusted P<0.05) are shown. Asterisks denote significance versus the Metabolically Preserved group (*P<0.05, **P<0.01, ***P<0.001). **D**: log fold changes of the same core-pathway proteins in Lean-IR (x-axis) versus Obese-IS (y-axis), each relative to the Metabolically Preserved group. Point shape indicates the phenotype(s) in which a protein was significant. A log fold change of 0 on either axis denotes a protein that was not significantly differentially expressed in that phenotype.

Despite these divergent, phenotype-specific profiles, both discordant phenotypes shared a common insulin–endocrine core, with up-regulated LEP (leptin) and FGF21, alongside down-regulated IGFBP1, indicating a convergent IR hormonal state; a shared prothrombotic adipokine, SERPINE1 (PAI-1), consistent with adipose dysfunction accompanying IR, independent of the adiposity burden. In an exploratory two-sample Mendelian randomisation analysis (**Supplementary Data 1**), genetically predicted FGF21 was associated with metabolic disorders (cis-MR), and higher LEP with heart failure and peripheral artery disease (trans-MR).

Class 5 (Obese-IR/hyperglycemia) showed extensive enrichment of cytokine signaling and lipid-atherosclerosis related pathways, together with those related to leukocyte migration, adhesion, and proliferation, indicating an advanced inflammatory–metabolic remodelling (**Figure 4A-B).** Moreover, the proteome characterising the two discordant phenotypes: IL-6 (Lean-IR-specific), ADH4 and KRT8/KRT18 (Obese-IS-specific), and IGFBP1 (shared), reached their greatest dysregulation in Class 5, reflecting Class 5 as highest-risk group, and representing a convergence of both Classes 3 and 4 discordant signatures (**Table S15**). Details of proteomic pathway analyses and their associations with latent classes and are in **Figures S15-S21** and Supplementary Data 2-5.

### Prognostic validation of the decision tree model in CHARLS and the UK Biobank

Finally, to link our classification to prognostic outcomes, we applied the decision tree–derived 5 classes to CHARLS (n=12,145) and the UK Biobank (n= 344,817). Using surrogate indicators for IR (elevated TG:HDL ratio) and visceral adiposity (central obesity by WC), together with high SBP and high HbA1c, the model reproducibly identified 5 cardiometabolic classes (**Table S16-S17 and Figure S21-S22**). Although Class 3 (Lean-IR) had significantly higher abundance of IR measured by TG:HDL ratio compared to Class 4, FPG and HbA1c were only modestly higher in Class 3 than in Class 4. These likely confirm that as a surrogate IR measure, TG:HDL ratio does not fully replace HOMA-IR (correlation of 0.52 in the discovery cohort) and shows a weaker correlation to hyperglycemia.

For longitudinal follow-up in both the UK Biobank and CHARLS, Kaplan–Meier and multivariable Cox analyses showed lowest event-free survival in Metabolically Preserved Class 1 and highest rates CVD events in Obese-IR (Class 5, **Figure 5**). In the UK Biobank, after the adjustment for age and sex, HR for the composite outcome of MI and stroke were 1.52 (95% CI, 1.43-1.61) for metabolically preserved isolated hypertension, 1.86 (1.75-1.98) for Lean-IR, 1.85 (1.75-1.97) for Obese-IS, and 2.75 (2.56-2.95) for Obese-IR, compared with metabolically preserved group. Notably, Lean-IR and Obese-IS conferred comparable risk (HR 1.86 vs 1.85; Wald test p=0.902), despite their distinct metabolic profiles, whereas Obese-IR carried significantly higher risk than all other phenogroups (all Wald test p<0.001). Event rates per 1,000 person-years and the HR and 95% CI when adjusted for Model 2 are shown in **Table S18-S19.** The same prognostic significance was also confirmed in the CHARLS cohort, using self-reported CVD and stroke outcomes (**Table S18-S19**).

**Figure 5.**
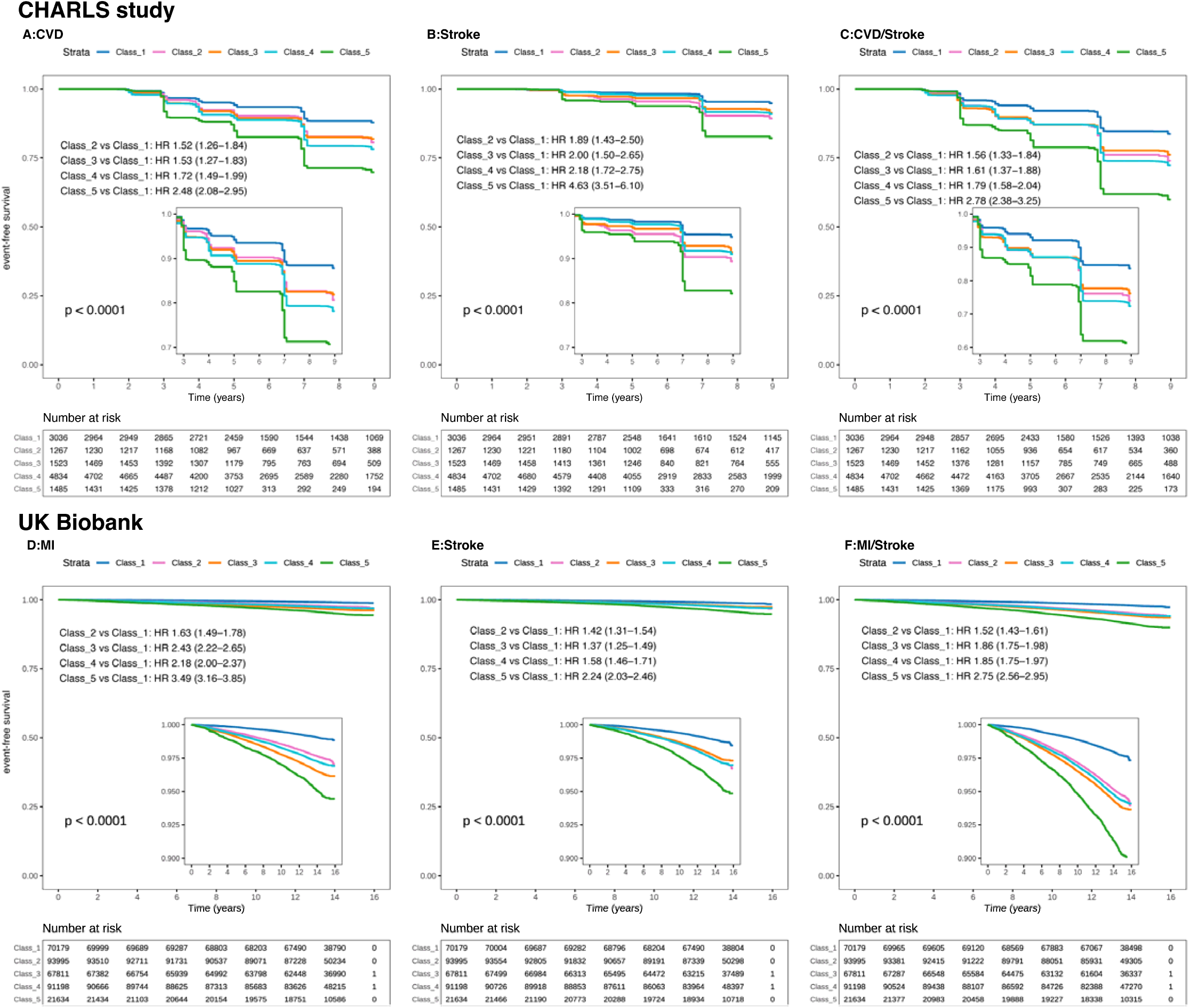
Longitudinal outcomes of the 5 latent classes in the CHARLS study and the UK Biobank. **A-C**. Kaplan-Meier curves of incident cardiovascular disease (CVD), stroke and CVD/stroke outcomes in 5 latent classes in CHARLS (n=12,145). Hazard ratio (HR) and 95% confidence interval (CI) were derived from COX regression and adjusted for age and sex. In CHARLS, IR was replaced by the surrogate indicator high TG:HDL ratio (≥1.51 mmol/L in men and 0.84 mmol/L in women), and visceral adiposity was replaced by the surrogate indicator central obesity (waist circumference≥80cm for female and ≥90cm for male). **D-F**. Kaplan-Meier curves of incident myocardial infarction (MI), stroke and MI/stroke outcomes in 5 latent classes in the UK Biobank (n= 34,4817). In the UK Biobank, IR was replaced by the surrogate indicator high TG:HDL ratio (≥3.5 mmol/L in male and 2.5 mmol/L in female), and visceral adiposity was replaced by the surrogate indicator central obesity (WC≥88cm for female and ≥102cm for male).

## DISCUSSION

Data-driven classification with expanded cardiometabolic traits identified early phenogroups that are characterised by distinct metabolic profiles rather than a single adiposity–driven continuum. Lean-IR and Obese-IS showed discordant adiposity and insulin/glycemic status, despite this, both exhibited a similar prevalence of subclinical diastolic dysfunction, significantly higher than in the Metabolically Preserved group. A parsimonious four-variable classifier (visceral adiposity, insulin resistance, elevated SBP and HbA1c) reproduced the 5 phenogroups, where plasma proteomics (validated in PICMAN and the UK Biobank) revealed an inflammatory emphasis in Lean-IR and a stronger hepatic metabolic/injury signature in Obese-IS, upon a shared insulin– endocrine core proteomic signature in both. These findings point to how convergent cardiac risk may arise from divergent biology, conceptualising how different individuals may benefit from earlier, targeted prevention.

### Data-driven phenotypes reveal underlying heterogeneity and subclinical cardiac dysfunction

Although MetS is used to assess cardiometabolic risk, its binary, threshold-based definition oversimplifies a biologically heterogeneous process.^29,30^ The CKM syndrome refines this by staging progression from dysfunctional adiposity to overt disease, enabling earlier, stage-guided prevention.^4^ Nonetheless, CKM staging captures the extent of cardiometabolic progression more than its underlying biology, and phenogroups with differing driver pathophysiology may fall within the same stage. In this study, a data-driven classification with expanded cardiometabolic traits revealed phenotypes not captured by MetS or conventional metabolic-health/BMI definitions, nor distinguished by CKM staging. This was explained in part by individual risk factors, particularly insulin resistance, conferring substantial risk even in the absence of other overt abnormalities.^11,13^ In the RESET cohort, CKM staging distinguished only the metabolically preserved group (Stage 1 was 77.4% Class 1), whereas higher stages grouped the opposing Lean-IR and Obese-IS phenotypes together (**Figure S12**). Nearly half (47.8%, **Figure S3**) of individuals labelled MHO by BMI and conventional risk factors^5^, are more appropriately assigned as Obese-IR (Class 5), characterised by pronounced visceral adiposity, hepatic steatosis, and insulin resistance.

The data-driven phenogroups also differed in subclinical cardiac function, in line with established evidence that IR and adiposity each impair the heart. IR reduces GLS even at normal weight,^15^ and MHO individuals show impaired strain and diastolic function despite a favourable metabolic profile.^31,32^ Published studies, however, stratified risk using BMI and conventional metabolic criteria, and stopped at phenotypic description. We advance this in two ways. First, our phenotypes emerge from the data-driven integration of multiple expanded risk factors. On this basis, the opposing phenotypes Lean-IR and Obese-IS converge on a similar elevated burden of subclinical cardiac dysfunction. Second, unlike prior echocardiographic studies, we link this shared cardiac phenotype to its molecular basis, which we examined in the plasma proteome.

### Distinct and shared proteomic signatures of the discordant phenotypes

At the molecular level, the two discordant phenotypes carried predominantly distinct proteomic signatures upon a shared endocrine core, mirroring their divergent physiology. In Lean-IR, inflammatory signalling centred on interleukin-6 (IL6), a cytokine that impairs insulin signalling and drives coronary microvascular endothelial inflammation, the proposed initiating step in heart failure with preserved ejection fraction (HFpEF).^33,34^ The accompanying AGE–RAGE signalling is consistent with this, as advanced glycation end-products fuel myocardial inflammation and cross-link the extracellular matrix, stiffening the myocardium and impairing diastolic function.^35^ Together these implicate inflammation as the route from IR to early cardiac injury in lean individuals, independent of adiposity.

The up-regulation of liver-enriched enzymes (HAO1, GSTA1, ADH1B and FTCD) was shared by both discordant phenotypes but more pronounced in Obese-IS, pointing to a shared subclinical hepatic stress that intensifies with adiposity.^36^ Because these are intracellular enzymes, their circulating rise most likely reflects hepatocyte leakage rather than increased hepatic activity, in keeping with MASLD proteomic maps that trace circulating changes to hepatic cell-type origin. ^37^ Most notably, the Obese-IS-specific marker KRT18 (cytokeratin-18), released during hepatocyte apoptosis and an established circulating marker of steatohepatitis,^38^ marks a continuum from shared hepatic stress toward overt hepatocyte injury in the obese phenotype. Both discordant phenotypes also shared an endocrine core. Leptin and FGF21 rise in obesity and IR, where target tissues become resistant to their actions, so high circulating levels reflect hormonal resistance rather than a healthy compensatory response.^39^ These inferences remain hypothesis-generating, as plasma proteomics cannot localise the tissue of origin.

### Clinical and translational implications

The data-driven phenogroups and their molecular difference have translational implications. The discordant phenotypes and their circulating signatures may enable earlier and more precise recognition of individuals who are overlooked by conventional criteria. Two of the 4 variables in our classifier, HOMA-IR and DEXA-derived visceral adipose tissue, are not yet routine but are increasingly accessible.^40^ In the UK Biobank, Lean-IR and Obese-IS carried a higher risk of incident myocardial infarction and stroke than the metabolically preserved group (hazard ratios 1.86 and 1.85), indicating that these phenogroups are not benign but are at an early, potentially modifiable stage of disease. Their divergent biology also points to actionable pathways. The inflammatory route of Lean-IR centres on IL-6, which may imply a means for precision targeting with ziltivekimab.^41^ The shared metabolic axis marked by FGF21 calls to FGF21 analogues that improve hepatic fibrosis, insulin sensitivity and lipids in trials.^42,43^ Overall, our report raises the possibility that these pathways may be engaged earlier and more precisely, in phenotype-matched individuals identified before overt disease manifests. We do not claim improved risk stratification over existing scores, which would require formal head-to-head comparison, but rather that the data-driven phenogroups are outcome-relevant and mechanistically informative.

### Limitation

A notable limitation of our study is the cross-sectional design of the discovery cohort, which does not allow for the confirmation of longitudinal progression and direct prognostic comparison. Dichotomisation of continuous variables enhances clinical interpretability but may introduce information loss and threshold-dependent misclassification.The proteomic validation in PICMAN is limited in statistical power, although we were able to validate 70.5% of the proteomic profiles successfully in the UK Biobank cohort after multiple testing correction. In the CHARLS and UK Biobank validation cohorts, the TG:HDL-C surrogate may misclassify IR phenotypes. Our analysis showed a correlation of 0.52 between TG:HDL-C ratio and HOMA-IR in the discovery cohort, indicating that direct IR measurement is preferred for future validation in large-scale prognostic cohorts. Finally, the decision tree model is more appropriately interpreted as a clinically implementable approximation of latent phenotypes rather than an independent validation of their biological discreteness.

## CONCLUSION

In conclusion, deep-phenotyping based on expanded cardiometabolic variables reveals marked heterogeneity. Latent classes include adiposity–IR/dysglycemia discordant subgroups with higher prevalence of subclinical cardiac dysfunction. A supervised model successfully confirms the 5 phenogroups in validation cohorts of both Asian and white participants. Proteomic analyses demonstrate an inflammatory emphasis in Lean-IR and a stronger hepatic metabolic/injury signature in Obese-IS, upon a shared insulin–endocrine core in both. By linking distinct phenogroups to divergent underlying biology, this work provides a rationale for precision cardiometabolic prevention, in which different phenotypes may benefit from mechanism-targeted strategies. This hypothesis will require testing in interventional studies.

## Author Contributions

JF, JC, and RF contributed to the conception of the work. JF, JC, HL, RF, NM, AA, ML, DJH, MM, WW, LL, DL, contributed to the data collection. JF, JC, HL and LZ contributed to the data analysis. JF, JC, and RF drafted the article. All authors contributed to the interpretation of data and critical revision of the Article. All authors gave final approval of the version to be published.

## Data Availability

The discovery cohort is part of the RESET cohort study: the dataset used for analysis will be made available upon reasonable request. Singaporean-based external cohort: Platform for the interdisciplinary study of cardiovascular, metabolic and neurovascular diseases (PICMAN): the dataset used for analysis will be made available upon reasonable request. The China Health and Retirement Longitudinal Study (CHARLS) is a publicly available cohort in: https://charls.pku.edu.cn/en/. The UK Biobank is accessible to approved researchers worldwide for health-related research in the public interest in: https://www.ukbiobank.ac.uk/. The raw code used for analysis will be made available upon reasonable request.

## Data Availability

https://www.ukbiobank.ac.uk/

## Acknowledgement and Group Information

Project RESET Investigators are listed in the Supplement Appendix. We acknowledge the RESET HQ Clinical Team comprising of Beverly Wong Jingmei, Amirul Sufyan Bin Mohd Salleh, Amisha Sharma D/O Rajender Kumar, Mohamed Sultan Aynul Marliya, Chi Dam, Benjamin Loh Jia-Hui, Phylicia Aw Pei Chin, Suhanni Nigam, Tharisini D/O Soorianarayanan, and Zachariah Seet Zhong En, who supported the recruitment of the RESET cohort.

