## Appendix Methods, Tables and Figures for "Data-driven cardiometabolic phenogroups reveal distinct subclinical cardiac and proteomic profiles"

1. **Supplementary Methods**

Supplementary Methods 1: Criteria used to dichotomize the clinical indicators and definition of MetS
Supplementary Methods 2: The Olink Proteomic quality control protocol in PICMAN study
Supplementary Methods 3: latent class analysis (LCA) approach and parameters
Supplementary Methods 4: Proteomic validation of decision tree characterized latent phenotypes in the PICMAN study and the UK Biobank cohort

2. **Supplementary Tables**

Table S1 Latent class analysis (LCA) model metrics for determining best classes
Table S2 Residual Correlation Matrix for Assessing Local Independence in LCA
Table S3 Baseline socioeconomic characteristics of discovery cohort–RESET study
Table S4 Detail of baseline variables according to 5 Latent classes in discovery cohort–RESET study
Table S5 Detail of echocardiographic variables according to 5 Latent classes in discovery cohort–RESET study
Table S6 Detail of baseline variables according to 5 Latent classes in male in discovery cohort–RESET study
Table S7 Detail of baseline variables according to 5 Latent classes in female in discovery cohort–RESET study
Table S8 Prevalence of metabolic risk factors and 5 latent classes according to fasting glucose in discovery cohort–RESET study
Table S9 Latent class analysis (LCA) model using simplified metabolic risk factors metrics for determining best classes
Table S10 Sensitivity analyses to vary each binary threshold by ±5% to assess the latent classification compared with original binary threshold
Table S11 Sensitivity analysis employed Latent profile analysis (LPA) on continuous data
Table S12 Sensitivity analysis employed HCPC on continuous data clustering
Table S13 The predictive accuracy for the decision tree model in identifying unsupervised latent classes with 5-fold internal cross-validation
Table S14 Biological annotation of the 31 DEPs with |$\log_2$ fold-change| > 0.8 in PICMAN across these two phenotypes
Table S15 The Dominant Biological Modules, Pathways and Proteins with top fold change in each class in PICMAN study
Table S16 Cardiometabolic traits, phenotypes and events according to 5 Latent classes in validation cohort–CHARLS
Table S17 Cardiometabolic traits, phenotypes and events according to 5 Latent classes in validation cohort–UK Biobank
Table S18 Cox proportional models for evaluating the association between latent classes and incident outcomes
Table S19 Events numbers and events rates per 1000 person-years for CVD, stroke and CVD/stroke in CHARLS and UK Biobank

3. **Supplementary Figure**

Figure S1 Inclusion and exclusion flowcharts in CHARLS and UK Biobank
Figure S2 The heatmap of correlation matrix for all related continuous cardiometabolic traits
Figure S3 Distribution of the five latent classes across categories of metabolic health and WC/BMI
Figure S4 Comparison of the metabolic traits as continuous variables among five latent classes from LCA in RESET
Figure S5 Violin plot of the numbers of abnormal binary metabolic phenotypes among five latent classes
Figure S6 Sensitivity analysis of LCA model after excluding highly correlated variables.
Figure S7 Sensitivity analysis of using simplified metabolic risk factors for clustering.
Figure S8 Sensitivity analysis of choosing four latent classes.
Figure S9 Sensitivity analyses to vary each binary threshold by ±5% to assess the latent classification
Figure S10 Sensitivity analysis of Latent profile analysis (LPA) on continuous data
Figure S11 Sensitivity analysis of Hierarchical Clustering on Principal Components (HCPC) on continuous data
Figure S12 Distribution of five latent classes within CKM stages
Figure S13 SHAP identifying the importance of features in a multinomial logistic regression model for predicting five latent classes in discovery cohort–RESET study
Figure S14 Validation of decision tree model identified five classes in PICMAN study
Figure S15 Venn plot for proteins associated with HOMA-IR, VAT, SBP and HbA1c in the validation dataset (PICMAN)
Figure S16 Proteomic signatures of the 5 classes in the PICMAN study and the UK Biobank.
Figure S17 Top differential proteomic signatures of the 5 classes in the PICMAN study and the UK Biobank.

Figure S18 KEGG over-representation analysis of differentially expressed proteins corresponding to each class compared with Metabolically Preserved (Class1).
Figure S19 Gene Ontology Biological Pathway (GO:BP) over-representation analysis of differentially expressed proteins corresponding to each class compared with Metabolically Preserved (Class1).
Figure S20 Gene Set Enrichment Analysis (GSEA) of Gene Ontology Biological Pathway (GO:BP) among five latent classes in the validation dataset (PICMAN)
Figure S21 Gene Set Enrichment Analysis (GSEA) of KEGG pathways among five latent classes in the validation dataset (PICMAN)
Figure S22 Validation of decision tree identified latent phenotypes in CHARLS
Figure S23 Validation of decision tree identified latent phenotypes in UK Biobank

**Supplementary Method 1: Criteria used to dichotomize the clinical indicators and definition of MetS**

(1) Thresholds for dichotomizing the 14 clinical indicators used for LCA

| | | |
|---|---|---|
| Risk factors in IDF–Metabolic syndrome criteria | Central Obesity | Waist Circumference ≥ 90/80(Male/Female) |
| | Obesity | BMI≥27.5kg.m$^2$ |
| | High TG | TG≥1.7 mmol/L |
| | Low HDL | HDL-C<1.03/1.29 mmol/L (Male/Female) |
| | High FPG | FPG≥5.6 mmol/L |
| | High SBP | SBP≥130 mmHg |
| | High DBP | DBP≥85 mmHg |
| Expanded Risk factors | Visceral adiposity | VAT≥100 cm$^2$ |
| | High LDL | LDL-C≥3.4 mmol/L |
| | High HbA1c | HbA1c≥5.7% |
| | Insulin resistance | HOMA-IR≥2.5 |
| | High hsCRP | hsCRP≥1mg/L |
| | Hyperuricemia | UA≥0.42/0.36 mmol/L (Male/Female) |
| | Liver Steatosis | CAP≥254 dB/m |

(2) Classification of metabolic health and BMI/WC subgroups

For description of population characteristics, participants were classified into metabolically healthy or unhealthy subgroups within each BMI- or WC-based category. BMI categories were defined according to cutoffs for Asian cohorts: RESET, PICMAN and CHARLS (normal weight: 18.5-22.9kg/m$^2$; overweight: 23.0-27.49kg/m$^2$; obesity class ≥27.5) [1], and cutoffs for western cohort: UK Biobank (normal weight: 18.5–24.9 kg/m$^2$; overweight: 25.0–29.9 kg/m$^2$; obesity class ≥30.0 kg/m$^2$). WC status was determined using sex-specific thresholds (≥90 cm for men, ≥80 cm for women in RESET, PICMAN and CHARLS ; and ≥102 cm for men, ≥88 cm for women in UK Biobank )[2].

Metabolically healthy is defined as any two of the following: elevated triglycerides (≥1.7 mmol/L or receiving treatment for hypertriglyceridemia), reduced HDL cholesterol (below 1.03 mmol/L in men or 1.29 mmol/L in women, or receiving lipid-lowering therapy), elevated blood pressure (systolic, SBP ≥130 mmHg, diastolic, DBP ≥85 mmHg, or current antihypertensive treatment), or impaired fasting glucose (≥5.6 mmol/L or current anti-diabetic therapy).

(3) Metabolic syndrome definition by IDF and AHA/NHLBI

MetS was defined according to the International Diabetes Federation criteria (IDF)[2], which require central obesity (high WC: (≥90 cm for men, ≥80 cm for women in RESET, PICMAN and CHARLS ; and ≥102 cm for men, ≥88 cm for women in UK Biobank) plus any two of the following: elevated triglycerides (≥1.7 mmol/L or receiving treatment for hypertriglyceridemia), reduced HDL cholesterol (below 1.03 mmol/L in men or 1.29 mmol/L in women, or receiving lipid-lowering therapy), elevated blood pressure (systolic, SBP ≥130 mmHg, diastolic, DBP ≥85 mmHg, or current antihypertensive treatment), or impaired fasting glucose (≥5.6 mmol/L or current anti-diabetic therapy). MetS was also defined according to the AHA/NHLBI statement[3], which require ≥3 of 5 components (high WC, elevated triglycerides, reduced HDL cholesterol, elevated blood pressure, impaired fasting glucose)

1. Appropriate body-mass index for Asian populations and its implications for policy and intervention strategies. Lancet (London, England). 2004;363(9403):157-163.
2. Alberti KGMM, Zimmet P, Shaw J. Metabolic syndrome—a new world-wide definition. A consensus statement from the international diabetes federation. Diabetic medicine. 2006;23(5):469-480.
3. Alberti KG, Eckel RH, Grundy SM, et al. Harmonizing the metabolic syndrome: a joint interim statement of the International Diabetes Federation Task Force on Epidemiology and Prevention; National Heart, Lung, and Blood Institute; American Heart Association; World Heart Federation; International Atherosclerosis

Society; and International Association for the Study of Obesity. Circulation. Oct 20 2009;120(16):1640-5. doi:10.1161/CIRCULATIONAHA.109.192644

**Supplementary Method 2: The Olink Proteomic quality control protocol in PICMAN study**
**Quality control (QC)** was performed according to Olink Explore HT standard protocols. Three internal controls were included in each sample: Incubation Control, Extension Control, and Amplification Control. The Extension Control was used for normalization to generate normalized protein expression (NPX) values, while the Incubation and Amplification Controls were used to monitor assay performance, immunoreaction efficiency, and potential technical errors.
In addition, three external controls were included in each run: Plate Control (pooled plasma from healthy individuals), Sample Control (pooled plasma from healthy individuals), and Negative Control (buffer). The Plate Control was used for normalization and overall run quality assessment. The Sample Control was used to evaluate inter-run and inter-plate variability. The Negative Control was used to detect unexpected background signals in assays.

**Plate-Level Quality Assessment:** QC parameters were evaluated to detect potential technical deviations caused by instrumental or operational errors. The following criteria were applied:

1. QC for all samples
    - Samples and external controls were flagged as failed if they exhibited low total counts within a block.
    - Samples and external controls failed if counts for any internal control were below predefined thresholds.
    - Samples received warnings if internal control counts were low but above failure thresholds.
2. Additional QC for Plate Controls and Negative Controls
    - Plate Controls failed if internal control counts deviated from expected ranges.
    - A block failed if an insufficient number of Plate Controls passed QC.
    - Negative Controls failed if excessive assay counts were detected relative to internal controls.
    - Assays received warnings if elevated counts were observed across Negative Controls.
    - A block failed if too few Negative Controls passed QC.

Only datapoints passing QC were included in downstream analyses.

**QC Summary:** All 120 samples in the Explore HT panel passed quality control (100% sample pass rate; 100% datapoint pass rate).

**Data Processing:** Protein expression data were reported as normalized protein expression (NPX) values on a $log_2$ scale. NPX values represent relative protein quantification derived from matched counts using next-generation sequencing (NGS) as the readout platform.

.

**Supplementary Method 3: latent class analysis (LCA) approach and parameters**

We implemented latent class analysis (LCA) to uncover latent cardiometabolic phenotypes, which is a model-based classification method for categorical or binary indicators by providing probabilistic class membership. To evaluate the local independence assumption, residual correlations between pairs of indicators were calculated as the difference between observed correlations and model-implied correlations under the fitted LCA model. Variable pairs with absolute residual correlations ≥0.10 were considered to indicate potential local dependence. The local independence assumption of LCA model was largely satisfied, with only modest residual correlations observed between high SBP and DBP, triglycerides and LDL cholesterol, and triglycerides and low HDL cholesterol (|r| > 0.10). The sensitivity analysis of excluding highly correlated variables (exclude high TG, high LDL and high DBP and retain low HDL and high SBP) for LCA showed that subtypes similar with the original latent classes still existed.

To determine the optimal number of latent classes, models specifying 2 to 6 classes were fitted and evaluated using the Bayesian Information Criterion (BIC). The model with the minimum BIC was selected as the optimal solution. LCA was conducted using R package poLCA, each model was fitted with 30 random initializations (nrep = 30), and the maximum number of expectation–maximization iterations was set at 5000, allowing sufficient iterations for convergence. Stability of latent class solution was evaluated using bootstrap resampling (B=500). For each bootstrap sample, LCA was refitted and class assignments were compared with the original model using adjusted Rand index (ARI) and classification agreement. As a results, the evaluation of LCA model demonstrated a classification quality with entropy of 0.79 and mean ARI of 0.68 (SD 0.13) in 500 bootstrap resampling. The conditional probabilities (P[phenotype | class]) of each binary-defined metabolic phenotype across the latent classes were shown by R package ComplexHeatmap. The within-class prevalence of each metabolic phenotype among latent classes was visualized by radar plot.

**Supplementary Method 4: Proteomic validation of decision tree characterized latent phenotypes in the PICMAN study and the UK Biobank cohort**

Latent classes identified by supervised decision tree model were first validated in another Singaporean-based external cohort: Platform for the interdisciplinary study of cardiovascular, metabolic and neurovascular diseases (PICMAN), which also included participants with ASCVD risk ≥ 5%.120 enrolled participants were clinically deep-phenotyped and underwent plasma multi-omic profiling. In the PICMAN study, considering the sample size limitation and difference in VAT measurements, IR was defined as HOMA-IR≥66.6th percentile, and visceral adiposity was defined as VAT volume ($cm^3$) by MRI≥ 66.6th percentile.

Plasma proteomic profiling was performed using the Olink Explore HT platform, quantifying 5416 proteins in 120 participants of the PICMAN cohort. Protein expression levels were processed using $\log_2$-transformed normalized protein expression (NPX) values. Given the high dimensionality and limited sample size, a two-step strategy adapted from prior multi-omics studies (*T. Takeuchi et al. Nature 2023 Vol. 621 Issue 7978 Pages 389-395*), was implemented to identify proteins associated with latent classes. First, linear models implemented in the R package limma were applied using continuous clinical traits corresponding to decision-tree node variables as outcomes to pre-screen proteins showing linear associations. Second, proteins significantly associated with these continuous traits were carried forward to pairwise differential expression analyses across latent classes using limma, with Class 1 specified as the reference group. Volcano plots were generated to visualize proteins with the largest fold changes and statistical significance across comparisons. Functional enrichment analyses were conducted for differentially expressed proteins using both over-representation analysis (ORA) and gene set enrichment analysis (GSEA) based on Gene Ontology (GO) and Kyoto Encyclopedia of Genes and Genomes (KEGG) pathways. Finally, latent classes were annotated with putative biological modules according to top differential proteins (Absolute logFC >0.8) and the dominant pathways enriched within each class.

Latent classes identified by supervised decision tree model were validated in the UK Biobank, which is a large-scale prospective cohort study comprising participants aged 40–69 years. Individuals of self-reported White ethnicity were included in this validation analysis. To match the analysis for RESET and PICMAN, participants with prevalent myocardial infarction (MI), stroke, cancer, or missing follow-up data and cardiometabolic metrics were excluded. In UK Biobank, IR was replaced by the surrogate indicator high TG:HDL ratio (≥3.5 mmol/L in male and 2.5 mmol/L in female), and visceral adiposity was replaced by the surrogate indicator central obesity (WC≥88cm for female and ≥102cm for male). Protein levels were measured using Olink (2923 proteins) as a part of UKB-PPP. Given the large sample size, no pre-screening was performed, and proteins with adjusted $P < 0.05$ using the Benjamini–Hochberg false discovery rate (FDR) were considered significant. Proteins consistently identified in both the PICMAN and UK Biobank cohorts were labelled in volcano plots.

**Table S1 Latent class analysis (LCA) model metrics for determining best classes**

| Classes | logLik | AIC | BIC | Gsq | Chisq |
|---|---|---|---|---|---|
| 2 | -7711.107 | 15480.214 | 15623.5085 | 3112.28519 | 25570.083 |
| 3 | -7630.4409 | 15348.8817 | 15566.2941 | 2950.95293 | 24870.9749 |
| 4 | -7557.2022 | 15232.4044 | 15523.9346 | 2804.47562 | 17859.1473 |
| **5** | **-7488.4457** | **15124.8915** | **15490.5395** | **2666.96267** | **17391.7546** |
| 6 | -7449.7155 | 15077.431 | 15517.1969 | 2589.50218 | 17348.4606 |

logLik = log-likelihood; AIC = Akaike Information Criterion; BIC = Bayesian Information Criterion; Gsq = Likelihood ratio chi-square statistic ($G^2$); Chisq = Pearson chi-square statistic

**Table S2 Residual Correlation Matrix for Assessing Local Independence in LCA**

| | Obesity | Central_ Obesity | Visceral_ adiposity | low_HDL | high_TG | high_LDL | high_FPG | high_ HbA1c | IR | high_SBP | high_DBP | Liver_ Steatosis | high_ hsCRP | Hyper-uricemia |
|---|---|---|---|---|---|---|---|---|---|---|---|---|---|---|
| Obesity | 0.000 | -0.001 | 0.007 | 0.005 | -0.023 | 0.000 | -0.055 | -0.037 | -0.029 | -0.035 | 0.014 | -0.003 | 0.016 | 0.020 |
| Central_ Obesity | -0.001 | 0.000 | 0.004 | 0.010 | -0.030 | 0.038 | -0.027 | -0.034 | -0.029 | 0.015 | -0.042 | 0.018 | -0.008 | -0.015 |
| Visceral_ adiposity | 0.007 | 0.004 | 0.000 | -0.055 | 0.005 | -0.005 | -0.032 | -0.023 | -0.040 | -0.005 | 0.017 | -0.020 | 0.002 | -0.020 |
| low_HDL | 0.005 | 0.010 | -0.055 | 0.000 | 0.110 | 0.015 | 0.010 | -0.001 | 0.006 | 0.023 | 0.021 | -0.001 | 0.062 | 0.003 |
| high_TG | -0.023 | -0.030 | 0.005 | 0.110 | 0.000 | 0.119 | -0.047 | -0.013 | 0.030 | 0.007 | 0.022 | 0.046 | 0.012 | 0.095 |
| high_LDL | 0.000 | 0.038 | -0.005 | 0.015 | 0.119 | 0.000 | -0.033 | -0.032 | 0.024 | 0.034 | 0.046 | 0.026 | 0.071 | 0.072 |
| high_FPG | -0.055 | -0.027 | -0.032 | 0.010 | -0.047 | -0.033 | 0.000 | 0.062 | 0.003 | 0.016 | -0.032 | -0.029 | -0.029 | -0.024 |
| high_ HbA1c | -0.037 | -0.034 | -0.023 | -0.001 | -0.013 | -0.032 | 0.062 | 0.000 | -0.065 | 0.004 | -0.027 | -0.016 | 0.006 | -0.015 |
| IR | -0.029 | -0.029 | -0.040 | 0.006 | 0.030 | 0.024 | 0.003 | -0.065 | 0.000 | 0.019 | 0.047 | 0.013 | -0.003 | 0.013 |
| high_SBP | -0.035 | 0.015 | -0.005 | 0.023 | 0.007 | 0.034 | 0.016 | 0.004 | 0.019 | 0.000 | 0.207 | 0.005 | 0.015 | -0.018 |
| high_DBP | 0.014 | -0.042 | 0.017 | 0.021 | 0.022 | 0.046 | -0.032 | -0.027 | 0.047 | 0.207 | 0.000 | -0.016 | -0.030 | 0.013 |
| Liver_ Steatosis | -0.003 | 0.018 | -0.020 | -0.001 | 0.046 | 0.026 | -0.029 | -0.016 | 0.013 | 0.005 | -0.016 | 0.000 | -0.016 | 0.027 |
| high_ hsCRP | 0.016 | -0.008 | 0.002 | 0.062 | 0.012 | 0.071 | -0.029 | 0.006 | -0.003 | 0.015 | -0.030 | -0.016 | 0.000 | 0.056 |
| Hyper uricemia | 0.020 | -0.015 | -0.020 | 0.003 | 0.095 | 0.072 | -0.024 | -0.015 | 0.013 | -0.018 | 0.013 | 0.027 | 0.056 | 0.000 |

The local independence assumption of LCA model was largely satisfied, with only modest residual correlations observed between high SBP and DBP, triglycerides and LDL cholesterol, and triglycerides and low HDL cholesterol (|r| > 0.10)

**Table S3 Baseline socioeconomic characteristics of discovery cohort–RESET study**

| Variable | Overall |
|---|---|
| n | 1034 |
| Age (mean (SD)) | 63.0 (5.3) |
| Gender = Male (%) | 703 (68.0) |
| Race (%) | |
| Chinese | 937 (91.0) |
| Indian | 54 ( 5.2) |
| Malay | 15 ( 1.5) |
| Others | 24 ( 2.3) |
| Marital status = Married (%) | 852 (82.7) |
| Monthly household income(SGD) = >$5,000 (%) | 555 (53.9) |
| Housing (%) | |
| Condominium/ Apartment | 326 (31.7) |
| HDB | 434 (42.1) |
| HDB Executive Maisonette | 66 ( 6.4) |
| Landed property | 189 (18.3) |
| Others | 15 ( 1.5) |
| Education (%) | |
| ITE/ Polytechnic/ Junior College | 251 (24.4) |
| Post Graduate Studies | 271 (26.3) |
| Primary School | 6 ( 0.6) |
| Secondary School | 121 (11.7) |
| University | 381 (37.0) |
| Employment_status (%) | |
| Full-time | 319 (31.0) |
| Homemaker | 43 ( 4.2) |
| Part-time | 172 (16.7) |
| Retired | 461 (44.8) |
| Unemployed | 35 ( 3.4) |
| Night_shift = Yes (%) | 97 ( 9.4) |
| Religion (%) | |
| Buddhist | 197 (19.1) |
| Christian/ Catholic | 492 (47.8) |
| Free thinker | 230 (22.3) |
| Hindu | 30 ( 2.9) |
| Muslim | 24 ( 2.3) |
| Others | 57 ( 5.5) |

**Table S4 Detail of baseline variables according to 5 Latent classes in discovery cohort–RESET study**

| Variable | Overall | Class 1 | Class 2 | Class 3 | Class 4 | Class 5 | p |
|---|---|---|---|---|---|---|---|
| n | 1034 | 244 | 244 | 140 | 211 | 195 | |
| Age , mean (SD), y | 63.0 (5.3) | 63.2 (4.8) | 63.8 (4.5) | 63.3 (5.2) | 63.3 (4.9) | 61.1 (6.6) | <0.001 |
| Male (%) | 703 (68.0) | 165 (67.6) | 167 ( 68.4) | 85 (60.7) | 146 (69.2) | 140 (71.8) | 0.300 |
| Race (%) | | | | | | | 0.031 |
| Chinese | 937 (91.0) | 229 (94.2) | 233 (95.5) | 125 (89.3) | 184 (88.5) | 166 (85.1) | |
| Indian | 54 ( 5.2) | 8 ( 3.3) | 6 ( 2.5) | 8 ( 5.7) | 13 ( 6.2) | 19 ( 9.7) | |
| Malay | 15 ( 1.5) | 3 ( 1.2) | 1 ( 0.4) | 2 ( 1.4) | 4 ( 1.9) | 5 ( 2.6) | |
| Others | 24 ( 2.3) | 3 ( 1.2) | 4 ( 1.6) | 5 ( 3.6) | 7 ( 3.4) | 5 ( 2.6) | |
| Body mass index , mean (SD), kg/m$^2$ | 24.9 (6.4) | 22.3 (2.3) | 22.5 (2.0) | 23.9 (1.9) | 27.4 (11.8) | 29.0 (3.5) | <0.001 |
| Waist circumference, mean (SD), cm | 84.2 (15.0) | 78.7 (10.6) | 78.0 (12.8) | 79.1 (17.3) | 89.0 (15.9) | 97.2 (8.5) | <0.001 |
| VAT area, mean (SD), cm$^2$ | 92.6 (42.3) | 61.2 (20.5) | 64.7 (18.9) | 86.5 (22.8) | 117.0 (32.8) | 144.9 (37.0) | <0.001 |
| TG, mean (SD), mmol/L | 1.2 (0.7) | 1.0 (0.4) | 1.0 (0.3) | 1.5 (1.0) | 1.4 (0.6) | 1.6 (0.8) | <0.001 |
| HDL-C , mean (SD), mmol/L | 1.6 (0.4) | 1.8 (0.5) | 1.8 (0.4) | 1.5 (0.4) | 1.5 (0.4) | 1.3 (0.3) | <0.001 |
| LDL-C, mean (SD), mmol/L | 3.3 (1.0) | 3.4 (1.0) | 3.4 (0.9) | 3.1 (0.9) | 3.4 (0.9) | 2.9 (0.9) | <0.001 |
| Lp(a), mean (SD) | 18.0 (22.9) | 19.8 (23.5) | 17.6 (23.1) | 21.7 (25.7) | 17.2 (21.7) | 14.4 (20.3) | 0.033 |
| FPG, mean (SD), mmol/L | 5.3 (1.4) | 5.0 (1.1) | 4.8 (0.5) | 6.3 (1.6) | 4.9 (0.4) | 6.3 (2.0) | <0.001 |
| HbA1c, mean (SD), % | 6.0 (0.9) | 5.8 (0.7) | 5.6 (0.5) | 6.5 (0.9) | 5.7 (0.3) | 6.7 (1.2) | <0.001 |
| Fasting insulin, mean (SD), mU/L | 11.0 (8.2) | 6.1 (2.7) | 6.2 (2.4) | 12.9 (6.7) | 11.6 (5.9) | 21.2 (10.0) | <0.001 |
| HOMA-IR, mean (SD) | 2.8 (2.6) | 1.3 (0.6) | 1.3 (0.5) | 3.5 (2.1) | 2.5 (1.3) | 6.0 (3.5) | <0.001 |
| SBP, mean (SD), mmHg | 132.2 (16.1) | 116.9 (8.5) | 144.4 (11.6) | 132.3 (15.2) | 135.3 (16.1) | 132.4 (13.8) | <0.001 |
| DBP, mean (SD), mmHg | 81.7 (9.3) | 74.8 (8.1) | 85.0 (8.3) | 81.2 (8.0) | 84.4 (8.9) | 83.5 (8.9) | <0.001 |
| Liver fat by CAP, mean (SD), dB/m | 242.1 (51.9) | 206.9 (39.2) | 216.1 (35.1) | 248.2 (48.1) | 264.9 (43.8) | 289.5 (43.1) | <0.001 |
| hsCRP, mean (SD), mg/L | 1.2 (2.1) | 1.0 (2.4) | 1.0 (1.4) | 1.4 (2.7) | 1.3 (2.0) | 1.7 (2.2) | 0.003 |
| Uric acid, mean (SD), mmol/L | 0.33 (0.07) | 0.32 (0.07) | 0.31 (0.07) | 0.34 (0.07) | 0.35 (0.07) | 0.35 (0.08) | <0.001 |
| eGFR, mean (SD), mL/min/1.73m$^2$ | 84.2 (11.8) | 85.6 (9.7) | 85.5 (10.9) | 83.4 (12.2) | 83.0 (11.9) | 82.9 (14.2) | 0.024 |
| MoCA score , mean (SD) | 25.2 (2.5) | 25.2 (2.4) | 25.2 (2.4) | 25.2 (2.3) | 25.2 (2.9) | 25.1 (2.5) | 0.981 |
| Alcohol status (%) | | | | | | | 0.274 |

| | | | | | | | |
|---|---|---|---|---|---|---|---|
| Ex-drinker | 55 ( 5.5) | 10 ( 4.2) | 11 ( 4.6) | 9 ( 6.5) | 9 ( 4.4) | 16 ( 8.5) | |
| No | 506 (50.2) | 125 (52.7) | 128 ( 53.8) | 73 (52.9) | 97 (47.3) | 83 (43.9) | |
| Current drinker | 446 (44.3) | 102 (43.0) | 99 ( 41.6) | 56 (40.6) | 99 (48.3) | 90 (47.6) | |
| Smoking status (%) | | | | | | | 0.010 |
| Ex-smoker | 114 (11.3) | 23 ( 9.7) | 25 ( 10.5) | 20 (14.5) | 21 (10.2) | 25 (13.2) | |
| No | 852 (84.6) | 206 (86.9) | 210 ( 88.2) | 115 (83.3) | 173 (84.4) | 148 (78.3) | |
| Current smoker | 41 ( 4.1) | 8 ( 3.4) | 3 ( 1.3) | 3 ( 2.2) | 11 ( 5.4) | 16 ( 8.5) | |
| Metabolic syndrome–IDF (%) | 255 (24.7) | 4 ( 1.6) | 7 ( 2.9) | 30 (21.4) | 66 (31.3) | 148 (75.9) | <0.001 |
| Metabolic syndrome–AHA/NHLBI (%) | 335 ( 32.4) | 9 ( 3.7) | 41 ( 16.8) | 59 ( 42.1) | 76 ( 36.0) | 150 ( 76.9) | <0.001 |
| Obesity classification by BMI (%) | | | | | | | <0.001 |
| Underweight | 26 ( 2.5) | 16 ( 6.6) | 9 ( 3.7) | 1 ( 0.7) | 0 ( 0.0) | 0 ( 0.0) | |
| Normal weight | 331 (32.0) | 133 (54.5) | 140 ( 57.4) | 40 (28.6) | 14 ( 6.6) | 4 ( 2.1) | |
| Overweight | 479 (46.3) | 94 (38.5) | 92 ( 37.7) | 99 (70.7) | 135 (64.0) | 59 (30.3) | |
| Obesity | 198 (19.1) | 1 ( 0.4) | 3 ( 1.2) | 0 ( 0.0) | 62 (29.4) | 132 (67.7) | |
| Obesity by BMI (%) | 198 (19.1) | 1 ( 0.4) | 3 ( 1.2) | 0 ( 0.0) | 62 (29.4) | 132 (67.7) | <0.001 |
| Central Obesity by WC (%) | 461 (44.6) | 41 (16.8) | 37 ( 15.2) | 38 (27.1) | 155 (73.5) | 190 (97.4) | <0.001 |
| Visceral adiposity by VAT (%) | 386 (37.3) | 3 ( 1.2) | 1 ( 0.4) | 33 (23.6) | 162 (76.8) | 187 (95.9) | <0.001 |
| low HDL (%) | 85 ( 8.2) | 4 ( 1.6) | 0 ( 0.0) | 18 (12.9) | 20 ( 9.5) | 43 (22.1) | <0.001 |
| high TG (%) | 178 (17.2) | 12 ( 4.9) | 4 ( 1.6) | 40 (28.6) | 56 (26.5) | 66 (33.8) | <0.001 |
| high LDL (%) | 453 (43.8) | 122 (50.0) | 117 ( 48.0) | 49 (35.0) | 117 (55.5) | 48 (24.6) | <0.001 |
| high FPG (%) | 244 (23.6) | 24 ( 9.8) | 10 ( 4.1) | 97 (69.3) | 0 ( 0.0) | 113 (57.9) | <0.001 |
| high HbA1c (%) | 652 (63.1) | 122 (50.0) | 104 ( 42.6) | 129 (92.1) | 109 (51.7) | 188 (96.4) | <0.001 |
| IR by HOMA-IR(%) | 376 (36.4) | 8 ( 3.3) | 2 ( 0.8) | 99 (70.7) | 77 (36.5) | 190 (97.4) | <0.001 |
| Diabetes status (%) | | | | | | | <0.001 |
| Normorglycemia | 363 (35.1) | 118 (48.4) | 136 ( 55.7) | 4 ( 2.9) | 102 (48.3) | 3 ( 1.5) | |
| Pre DM | 490 (47.4) | 103 (42.2) | 96 ( 39.3) | 80 (57.1) | 105 (49.8) | 106 (54.4) | |
| DM | 181 (17.5) | 23 ( 9.4) | 12 ( 4.9) | 56 (40.0) | 4 ( 1.9) | 86 (44.1) | |
| high SBP (%) | 563 (54.4) | 0 ( 0.0) | 244 (100.0) | 78 (55.7) | 135 (64.0) | 106 (54.4) | <0.001 |
| high DBP (%) | 361 (34.9) | 14 ( 5.7) | 129 ( 52.9) | 43 (30.7) | 100 (47.4) | 75 (38.5) | <0.001 |

| | | | | | | | |
|---|---|---|---|---|---|---|---|
| Hypertension (%) | 679 (65.7) | 36 (14.8) | 244 (100.0) | 95 (67.9) | 157 (74.4) | 147 (75.4) | <0.001 |
| Dyslipidemia (%) | 688 (66.5) | 158 (64.8) | 144 ( 59.0) | 93 (66.4) | 161 (76.3) | 132 (67.7) | 0.003 |
| Liver Steatosis (%) | 392 (37.9) | 24 ( 9.8) | 34 ( 13.9) | 53 (37.9) | 131 (62.1) | 150 (76.9) | <0.001 |
| high hsCRP (%) | 312 (30.2) | 40 (16.4) | 59 ( 24.2) | 44 (31.4) | 80 (37.9) | 89 (45.6) | <0.001 |
| Hyperuricemia (%) | 174 (16.8) | 22 ( 9.0) | 26 ( 10.7) | 40 (28.6) | 36 (17.1) | 50 (25.6) | <0.001 |
| MoCA Cognitive impairment (%) | 493 (50.6) | 115 (48.9) | 120 ( 52.4) | 60 (47.2) | 99 (49.3) | 99 (54.1) | 0.704 |
| Anti-diabetic drug (%) | 63 ( 6.1) | 11 ( 4.5) | 8 ( 3.3) | 19 (13.6) | 4 ( 1.9) | 21 (10.8) | <0.001 |
| Anti-hypertension drug (%) | 217 (21.0) | 24 ( 9.8) | 45 ( 18.4) | 29 (20.7) | 49 (23.2) | 70 (35.9) | <0.001 |
| Lipid-lowering drug (%) | 190 (18.4) | 38 (15.6) | 40 ( 16.4) | 33 (23.6) | 41 (19.4) | 38 (19.5) | 0.314 |
| Metabolic health–WC groups (%) | | | | | | | <0.001 |
| MHNWC | 374 (36.2) | 165 (67.6) | 156 ( 63.9) | 23 (16.4) | 29 (13.7) | 1 ( 0.5) | |
| MHHWC | 206 (19.9) | 37 (15.2) | 30 ( 12.3) | 8 ( 5.7) | 89 (42.2) | 42 (21.5) | |
| MUHNWC | 199 (19.2) | 38 (15.6) | 51 ( 20.9) | 79 (56.4) | 27 (12.8) | 4 ( 2.1) | |
| MUHHWC | 255 (24.7) | 4 ( 1.6) | 7 ( 2.9) | 30 (21.4) | 66 (31.3) | 148 (75.9) | |
| Metabolic health–BMI groups (%) | | | | | | | <0.001 |
| MHNW | 511 (49.4) | 201 (82.4) | 185 ( 75.8) | 31 (22.1) | 84 (39.8) | 10 ( 5.1) | |
| MHO | 69 ( 6.7) | 1 ( 0.4) | 1 ( 0.4) | 0 ( 0.0) | 34 (16.1) | 33 (16.9) | |
| MUNW | 325 (31.4) | 42 (17.2) | 56 ( 23.0) | 109 (77.9) | 65 (30.8) | 53 (27.2) | |
| MUO | 129 (12.5) | 0 ( 0.0) | 2 ( 0.8) | 0 ( 0.0) | 28 (13.3) | 99 (50.8) | |
| CKM stage (%) | | | | | | | |
| Stage 1 | 84 ( 8.5) | 65 ( 32.0) | 0 ( 0.0) | 3 ( 2.2) | 14 ( 6.8) | 2 ( 1.0) | |
| Stage 2 | 469 ( 47.6) | 72 ( 35.5) | 140 ( 57.9) | 73 ( 52.5) | 96 ( 46.6) | 88 ( 45.1) | |
| Stage 3 | 432 ( 43.9) | 66 ( 32.5) | 102 ( 42.1) | 63 ( 45.3) | 96 ( 46.6) | 105 ( 53.8) | |

Abbreviation: BMI, body mass index; WC, Waist circumference; VAT, Visceral adiposity tissue; TG, Triglyceride; LDL-C, Low density lipoprotein cholesterol; HDL-C, High density lipoprotein cholesterol; FPG, Fasting plasma glucose; HbA1c, Glycated Hemoglobin A1c; HOMA-IR, Homeostatic Model Assessment of Insulin Resistance; CAP, Controlled attenuation parameter (Db/m); hsCRP, High Sensitivity C-reactive Protein; SBP, systolic blood pressure; DBP, diastolic blood pressure; MHNW, Metabolically healthy normal weight; MUNW, Metabolically unhealthy normal weight; MHO, Metabolically healthy obese; MUO, Metabolically unhealthy obese; MHNWC, Metabolically healthy normal waist circumference; MUHWC, Metabolically unhealthy high waist circumference

**Table S5 Detail of echocardiographic variables according to 5 Latent classes in discovery cohort–RESET study**

| Variable | Overall | Class 1 | Class 2 | Class 3 | Class 4 | Class 5 | p |
|---|---|---|---|---|---|---|---|
| n | 1034 | 244 | 244 | 140 | 211 | 195 | |
| LVMI (median [IQR]) | 77.0 [65.0, 90.0] | 72.0 [62.0, 82.0] | 78.0 [67.0, 92.0] | 76.0 [64.0, 90.0] | 80.0 [69.0, 94.0] | 77.0 [62.0, 91.0] | <0.001 |
| RWT (median [IQR]) | 0.4 [0.4, 0.5] | 0.4 [0.3, 0.5] | 0.4 [0.4, 0.5] | 0.4 [0.4, 0.5] | 0.4 [0.4, 0.5] | 0.4 [0.4, 0.5] | <0.001 |
| LVEF (median [IQR]) | 63.9 [61.9, 67.1] | 64.0 [61.9, 67.4] | 64.3 [61.9, 67.1] | 64.6 [62.5, 67.1] | 63.7 [61.7, 67.2] | 63.6 [61.3, 66.3] | 0.265 |
| GLS (median [IQR]) | -19.5 [-21.0, -18.2] | -20.0 [-21.6, -18.6] | -19.9 [-21.1, -18.6] | -19.4 [-21.1, -18.2] | -19.3 [-20.6, -18.0] | -18.7 [-20.1, -17.7] | <0.001 |
| eprime_sep (median [IQR]) | 7.1 [6.0, 8.3] | 7.9 [6.7, 9.0] | 7.3 [6.2, 8.4] | 6.9 [5.8, 8.0] | 6.8 [5.8, 7.9] | 6.7 [5.7, 7.6] | <0.001 |
| eprime_lat (median [IQR]) | 9.3 [7.7, 10.7] | 10.0 [8.6, 11.6] | 9.6 [8.1, 11.0] | 8.9 [7.7, 10.1] | 8.9 [7.5, 10.2] | 8.3 [7.2, 10.2] | <0.001 |
| Ee_avg (median [IQR]) | 7.7 [6.4, 8.9] | 7.0 [5.9, 8.2] | 7.5 [6.5, 8.9] | 7.8 [6.6, 9.3] | 7.9 [6.5, 9.1] | 8.2 [7.0, 9.4] | <0.001 |
| EA (median [IQR]) | 0.9 [0.8, 1.1] | 1.0 [0.9, 1.3] | 1.0 [0.8, 1.2] | 0.9 [0.8, 1.1] | 0.9 [0.7, 1.0] | 0.8 [0.7, 0.9] | <0.001 |
| TRv (median [IQR]) | 2.1 [1.9, 2.3] | 2.2 [2.0, 2.3] | 2.1 [2.0, 2.4] | 2.1 [1.9, 2.3] | 2.1 [1.9, 2.3] | 2.1 [1.9, 2.3] | 0.473 |
| LAVI (median [IQR]) | 32.8 [27.8, 38.4] | 33.5 [28.8, 39.2] | 33.8 [28.4, 39.9] | 33.0 [28.1, 37.1] | 32.2 [27.6, 38.3] | 30.5 [25.9, 36.3] | 0.001 |
| PASP (median [IQR]) | 21.0 [18.0, 25.0] | 22.0 [18.5, 25.0] | 22.0 [19.0, 25.0] | 21.0 [18.0, 24.0] | 21.0 [18.0, 24.0] | 21.0 [17.0, 24.0] | 0.272 |
| LVH = 1 (%) | 60 ( 5.8) | 1 ( 0.4) | 14 ( 5.8) | 9 ( 6.5) | 19 ( 9.1) | 17 ( 8.7) | <0.001 |
| RWT_increased = 1 (%) | 568 (55.3) | 104 (42.8) | 139 (57.7) | 80 (57.6) | 119 (56.9) | 126 (64.6) | <0.001 |
| LV_geometry (%) | | | | | | | <0.001 |
| Concentric hypertrophy | 55 ( 5.4) | 1 ( 0.4) | 14 ( 5.8) | 9 ( 6.5) | 17 ( 8.1) | 14 ( 7.2) | |
| Concentric remodeling | 513 (50.0) | 103 (42.4) | 125 (51.9) | 71 (51.1) | 102 (48.8) | 112 (57.4) | |
| Eccentric hypertrophy | 5 ( 0.5) | 0 ( 0.0) | 0 ( 0.0) | 0 ( 0.0) | 2 ( 1.0) | 3 ( 1.5) | |
| Normal | 454 (44.2) | 139 (57.2) | 102 (42.3) | 59 (42.4) | 88 (42.1) | 66 (33.8) | |
| LVEF_reduced = 1 (%) | 4 ( 0.4) | 1 ( 0.4) | 0 ( 0.0) | 0 ( 0.0) | 2 ( 1.0) | 1 ( 0.5) | 0.509 |
| GLS_abnormal = 1 (%) | 204 (19.9) | 33 (13.6) | 43 (17.8) | 25 (18.0) | 47 (22.5) | 56 (28.7) | 0.002 |
| DD_present = Diastolic dysfunction (%) | 319 (31.1) | 40 (16.5) | 63 (26.1) | 51 (36.7) | 81 (38.8) | 84 (43.1) | <0.001 |
| DD_grade_2025 (%) | | | | | | | <0.001 |
| Normal | 708 (68.9) | 203 (83.5) | 178 (73.9) | 88 (63.3) | 128 (61.2) | 111 (56.9) | |
| Grade 1 | 244 (23.8) | 31 (12.8) | 52 (21.6) | 34 (24.5) | 62 (29.7) | 65 (33.3) | |
| Grade 2 | 74 ( 7.2) | 8 ( 3.3) | 11 ( 4.6) | 17 (12.2) | 19 ( 9.1) | 19 ( 9.7) | |
| Grade 3 | 1 ( 0.1) | 1 ( 0.4) | 0 ( 0.0) | 0 ( 0.0) | 0 ( 0.0) | 0 ( 0.0) | |
| LA_enlarged = 1 (%) | 453 (44.1) | 118 (48.6) | 116 (48.1) | 59 (42.4) | 91 (43.5) | 69 (35.4) | 0.045 |

LVMI, left ventricular mass index; RWT, relative wall thickness; LVEF, left ventricular ejection fraction; GLS, global longitudinal strain; eprime_sep (e′ sep), septal early diastolic mitral annular velocity; eprime_lat (e′ lat), lateral early diastolic mitral annular velocity'; Ee_avg (E/e′), ratio of early mitral inflow velocity to average early diastolic mitral annular velocity; EA (E/A), ratio of early to late (atrial) mitral inflow velocity; TRv, peak tricuspid regurgitation velocity; LAVI, left atrial volume index; PASP, pulmonary artery systolic pressure; LVH, left ventricular hypertrophy; DD_present, diastolic dysfunction (present); DD_grade_2025, diastolic dysfunction grade (2025 ASE/EACVI criteria); LA_enlarged, left atrial enlargement

**Table S6 Detail of baseline variables according to 5 Latent classes in male in discovery cohort–RESET study**

| Variable | Overall | Class 1 | Class 2 | Class 3 | Class 4 | Class 5 | p |
|---|---|---|---|---|---|---|---|
| n | 703 | 165 | 167 | 85 | 146 | 140 | |
| Age , mean (SD), y | 62.2 (5.5) | 62.2 (5.1) | 62.9 (4.7) | 62.1 (5.6) | 62.8 (5.0) | 60.7 (6.8) | 0.005 |
| Male (%) | 703 (100.0) | 165 (100.0) | 167 (100.0) | 85 (100.0) | 146 (100.0) | 140 (100.0) | NA |
| Race (%) | | | | | | | 0.101 |
| Chinese | 633 ( 90.3) | 153 ( 92.7) | 160 ( 95.8) | 77 ( 90.6) | 125 ( 86.8) | 118 ( 84.3) | |
| Indian | 40 ( 5.7) | 7 ( 4.2) | 4 ( 2.4) | 5 ( 5.9) | 10 ( 6.9) | 14 ( 10.0) | |
| Malay | 11 ( 1.6) | 3 ( 1.8) | 1 ( 0.6) | 1 ( 1.2) | 2 ( 1.4) | 4 ( 2.9) | |
| Others | 17 ( 2.4) | 2 ( 1.2) | 2 ( 1.2) | 2 ( 2.4) | 7 ( 4.9) | 4 ( 2.9) | |
| Body mass index , mean (SD), kg/m$^2$ | 25.0 (3.7) | 22.5 (2.4) | 22.8 (1.9) | 24.3 (1.6) | 26.7 (2.9) | 29.3 (3.4) | <0.001 |
| Waist circumference, mean (SD), cm | 85.6 (14.9) | 79.5 (10.5) | 79.0 (13.0) | 80.0 (19.3) | 90.9 (13.8) | 98.5 (7.7) | <0.001 |
| VAT area, mean (SD), cm$^2$ | 98.2 (41.9) | 66.6 (17.0) | 68.8 (15.3) | 92.7 (21.7) | 121.0 (31.9) | 150.0 (37.8) | <0.001 |
| TG, mean (SD), mmol/L | 1.5 (0.4) | 1.7 (0.4) | 1.7 (0.3) | 1.4 (0.3) | 1.4 (0.3) | 1.3 (0.3) | <0.001 |
| HDL-C , mean (SD), mmol/L | 1.3 (0.7) | 1.0 (0.4) | 1.0 (0.3) | 1.4 (1.1) | 1.5 (0.7) | 1.6 (0.9) | <0.001 |
| LDL-C, mean (SD), mmol/L | 3.2 (1.0) | 3.3 (1.0) | 3.4 (1.0) | 3.0 (0.9) | 3.4 (1.0) | 2.8 (0.9) | <0.001 |
| Lp(a), mean (SD) | 16.7 (21.6) | 18.6 (23.8) | 14.2 (18.4) | 20.1 (24.1) | 16.3 (20.3) | 15.8 (21.9) | 0.203 |
| FPG, mean (SD), mmol/L | 5.3 (1.3) | 5.0 (1.1) | 4.9 (0.6) | 6.2 (1.8) | 4.9 (0.3) | 6.2 (1.8) | <0.001 |
| HbA1c, mean (SD), % | 5.9 (0.9) | 5.8 (0.8) | 5.6 (0.5) | 6.4 (0.8) | 5.6 (0.3) | 6.6 (1.2) | <0.001 |
| Fasting insulin, mean (SD), mU/L | 11.1 (8.2) | 6.1 (2.9) | 6.1 (2.4) | 12.7 (6.8) | 11.8 (6.1) | 21.0 (9.9) | <0.001 |
| HOMA-IR, mean (SD) | 2.7 (2.4) | 1.4 (0.6) | 1.3 (0.5) | 3.5 (2.2) | 2.6 (1.4) | 5.7 (3.2) | <0.001 |
| SBP, mean (SD), mmHg | 131.6 (15.3) | 116.7 (8.4) | 143.5 (10.8) | 130.9 (15.6) | 134.8 (14.9) | 131.9 (12.0) | <0.001 |
| DBP, mean (SD), mmHg | 82.9 (8.9) | 76.2 (7.7) | 86.2 (7.7) | 82.2 (7.9) | 85.6 (8.2) | 84.6 (8.8) | <0.001 |
| Liver Fat by CAP, mean (SD), dB/m | 243.0 (51.4) | 207.4 (43.5) | 220.0 (32.9) | 247.0 (47.6) | 265.3 (44.2) | 286.7 (42.4) | <0.001 |
| hsCRP, mean (SD), mg/L | 1.2 (2.1) | 1.0 (2.4) | 0.9 (1.4) | 1.4 (3.2) | 1.2 (1.5) | 1.4 (2.0) | 0.163 |
| Uric acid, mean (SD), mmol/L | 0.35 (0.07) | 0.34 (0.06) | 0.33 (0.07) | 0.36 (0.07) | 0.37 (0.06) | 0.37 (0.08) | <0.001 |
| eGFR, mean (SD), mL/min/1.73m$^2$ | 82.6 (11.8) | 83.8 (9.9) | 84.5 (10.7) | 82.4 (12.9) | 80.0 (11.3) | 81.9 (14.4) | 0.008 |
| MoCA score , mean (SD) | 24.9 (2.4) | 25.1 (2.4) | 24.7 (2.4) | 24.7 (2.2) | 25.2 (2.7) | 24.9 (2.4) | 0.413 |
| Alcohol status (%) | | | | | | | 0.583 |

| | | | | | | | |
|---|---|---|---|---|---|---|---|
| Ex-drinker | 49 ( 7.2) | 9 ( 5.6) | 10 ( 6.2) | 8 ( 9.4) | 9 ( 6.4) | 13 ( 9.6) | |
| No | 280 ( 41.0) | 71 ( 44.4) | 74 ( 46.0) | 33 ( 38.8) | 52 ( 36.9) | 50 ( 36.8) | |
| Current drinker | 354 ( 51.8) | 80 ( 50.0) | 77 ( 47.8) | 44 ( 51.8) | 80 ( 56.7) | 73 ( 53.7) | |
| Smoking status (%) | | | | | | | 0.011 |
| Ex-smoker | 108 ( 15.8) | 20 ( 12.5) | 24 ( 14.9) | 20 ( 23.5) | 20 ( 14.2) | 24 ( 17.6) | |
| No | 538 ( 78.8) | 133 ( 83.1) | 134 ( 83.2) | 62 ( 72.9) | 112 ( 79.4) | 97 ( 71.3) | |
| Current smoker (%) | 37 ( 5.4) | 7 ( 4.4) | 3 ( 1.9) | 3 ( 3.5) | 9 ( 6.4) | 15 ( 11.0) | |
| Metabolic syndrome–IDF (%) | 159 ( 22.6) | 0 ( 0.0) | 1 ( 0.6) | 11 ( 12.9) | 40 ( 27.4) | 107 ( 76.4) | <0.001 |
| Obesity classification by BMI (%) | | | | | | | <0.001 |
| Underweight | 12 ( 1.7) | 9 ( 5.5) | 3 ( 1.8) | 0 ( 0.0) | 0 ( 0.0) | 0 ( 0.0) | |
| Normal weight | 197 ( 28.0) | 85 ( 51.5) | 88 ( 52.7) | 17 ( 20.0) | 7 ( 4.8) | 0 ( 0.0) | |
| Overweight | 348 ( 49.5) | 70 ( 42.4) | 73 ( 43.7) | 68 ( 80.0) | 98 ( 67.1) | 39 ( 27.9) | |
| Obesity | 146 ( 20.8) | 1 ( 0.6) | 3 ( 1.8) | 0 ( 0.0) | 41 ( 28.1) | 101 ( 72.1) | |
| Obesity (%) | 146 ( 20.8) | 1 ( 0.6) | 3 ( 1.8) | 0 ( 0.0) | 41 ( 28.1) | 101 ( 72.1) | <0.001 |
| Central Obesity by WC (%) | 272 ( 38.7) | 15 ( 9.1) | 9 ( 5.4) | 16 ( 18.8) | 97 ( 66.4) | 135 ( 96.4) | <0.001 |
| Visceral adiposity by VAT (%) | 289 ( 41.1) | 3 ( 1.8) | 1 ( 0.6) | 27 ( 31.8) | 122 ( 83.6) | 136 ( 97.1) | <0.001 |
| low HDL (%) | 46 ( 6.5) | 1 ( 0.6) | 0 ( 0.0) | 9 ( 10.6) | 11 ( 7.5) | 25 ( 17.9) | <0.001 |
| high TG (%) | 120 ( 17.1) | 8 ( 4.8) | 4 ( 2.4) | 20 ( 23.5) | 43 ( 29.5) | 45 ( 32.1) | <0.001 |
| high LDL (%) | 292 ( 41.5) | 80 ( 48.5) | 84 ( 50.3) | 25 ( 29.4) | 76 ( 52.1) | 27 ( 19.3) | <0.001 |
| high FPG (%) | 162 ( 23.0) | 17 ( 10.3) | 9 ( 5.4) | 58 ( 68.2) | 0 ( 0.0) | 78 ( 55.7) | <0.001 |
| high HbA1c (%) | 441 ( 62.7) | 83 ( 50.3) | 74 ( 44.3) | 76 ( 89.4) | 74 ( 50.7) | 134 ( 95.7) | <0.001 |
| IR by HOMA-IR(%) | 255 ( 36.3) | 7 ( 4.2) | 1 ( 0.6) | 58 ( 68.2) | 54 ( 37.0) | 135 ( 96.4) | <0.001 |
| Diabetes status (%) | | | | | | | <0.001 |
| Normorglycemia | 247 ( 35.1) | 79 ( 47.9) | 91 ( 54.5) | 3 ( 3.5) | 72 ( 49.3) | 2 ( 1.4) | |
| Pre DM | 345 ( 49.1) | 72 ( 43.6) | 66 ( 39.5) | 51 ( 60.0) | 73 ( 50.0) | 83 ( 59.3) | |
| DM | 111 ( 15.8) | 14 ( 8.5) | 10 ( 6.0) | 31 ( 36.5) | 1 ( 0.7) | 55 ( 39.3) | |
| high SBP (%) | 380 ( 54.1) | 0 ( 0.0) | 167 (100.0) | 42 ( 49.4) | 93 ( 63.7) | 78 ( 55.7) | <0.001 |
| high DBP (%) | 274 ( 39.0) | 12 ( 7.3) | 100 ( 59.9) | 27 ( 31.8) | 77 ( 52.7) | 58 ( 41.4) | <0.001 |
| Hypertension (%) | 473 ( 67.3) | 27 ( 16.4) | 167 (100.0) | 54 ( 63.5) | 115 ( 78.8) | 110 ( 78.6) | <0.001 |

| | | | | | | | |
|---|---|---|---|---|---|---|---|
| Dyslipidemia (%) | 455 ( 64.7) | 105 ( 63.6) | 102 ( 61.1) | 51 ( 60.0) | 107 ( 73.3) | 90 ( 64.3) | 0.159 |
| Liver Steatosis (%) | 269 ( 38.3) | 19 ( 11.5) | 25 ( 15.0) | 32 ( 37.6) | 88 ( 60.3) | 105 ( 75.0) | <0.001 |
| high hsCRP (%) | 202 ( 28.7) | 29 ( 17.6) | 38 ( 22.8) | 25 ( 29.4) | 52 ( 35.6) | 58 ( 41.4) | <0.001 |
| Hyperuricemia (%) | 125 ( 17.8) | 19 ( 11.5) | 20 ( 12.0) | 24 ( 28.2) | 28 ( 19.2) | 34 ( 24.3) | 0.001 |
| MoCA Cognitive impairment (%) | 373 ( 55.8) | 80 ( 50.3) | 96 ( 61.1) | 48 ( 59.3) | 73 ( 52.1) | 76 ( 57.6) | 0.279 |
| Anti-diabetic drug (%) | 37 ( 5.3) | 5 ( 3.0) | 7 ( 4.2) | 11 ( 12.9) | 1 ( 0.7) | 13 ( 9.3) | <0.001 |
| Anti-hypertension drug (%) | 156 ( 22.2) | 17 ( 10.3) | 32 ( 19.2) | 20 ( 23.5) | 35 ( 24.0) | 52 ( 37.1) | <0.001 |
| Lipid-lowering drug (%) | 130 ( 18.5) | 26 ( 15.8) | 28 ( 16.8) | 19 ( 22.4) | 25 ( 17.1) | 32 ( 22.9) | 0.414 |
| Metabolic health–WC groups (%) | | | | | | | <0.001 |
| MHNWC | 276 ( 39.3) | 120 ( 72.7) | 115 ( 68.9) | 14 ( 16.5) | 26 ( 17.8) | 1 ( 0.7) | |
| MHHWC | 113 ( 16.1) | 15 ( 9.1) | 8 ( 4.8) | 5 ( 5.9) | 57 ( 39.0) | 28 ( 20.0) | |
| MUHNWC | 155 ( 22.0) | 30 ( 18.2) | 43 ( 25.7) | 55 ( 64.7) | 23 ( 15.8) | 4 ( 2.9) | |
| MUHHWC | 159 ( 22.6) | 0 ( 0.0) | 1 ( 0.6) | 11 ( 12.9) | 40 ( 27.4) | 107 ( 76.4) | |
| Metabolic health–BMI groups (%) | | | | | | | <0.001 |
| MHNW | 340 ( 48.4) | 134 ( 81.2) | 122 ( 73.1) | 19 ( 22.4) | 60 ( 41.1) | 5 ( 3.6) | |
| MHO | 49 ( 7.0) | 1 ( 0.6) | 1 ( 0.6) | 0 ( 0.0) | 23 ( 15.8) | 24 ( 17.1) | |
| MUNW | 217 ( 30.9) | 30 ( 18.2) | 42 ( 25.1) | 66 ( 77.6) | 45 ( 30.8) | 34 ( 24.3) | |
| MUO | 97 ( 13.8) | 0 ( 0.0) | 2 ( 1.2) | 0 ( 0.0) | 18 ( 12.3) | 77 ( 55.0) | |

Abbreviation: BMI, body mass index; WC, Waist circumference; VAT, Visceral adiposity tissue; TG, Triglyceride; LDL-C, Low density lipoprotein cholesterol; HDL-C, High density lipoprotein cholesterol; FPG, Fasting plasma glucose; HbA1c, Glycated Hemoglobin A1c; HOMA-IR, Homeostatic Model Assessment of Insulin Resistance; CAP, Controlled attenuation parameter (Db/m); hsCRP, High Sensitivity C-reactive Protein; SBP, systolic blood pressure; DBP, diastolic blood pressure; MHNW, Metabolically healthy normal weight; MUNW, Metabolically unhealthy normal weight; MHO, Metabolically healthy obese; MUO, Metabolically unhealthy obese; MHNWC, Metabolically healthy normal waist circumference; MUHWC, Metabolically unhealthy high waist circumference

**Table S7 Detail of baseline variables according to 5 Latent classes in female in discovery cohort–RESET study**

| Variable | Overall | Class 1 | Class 2 | Class 3 | Class 4 | Class 5 | p |
|---|---|---|---|---|---|---|---|
| n | 331 | 79 | 77 | 55 | 65 | 55 | |
| Age , mean (SD), y | 64.7 (4.4) | 65.4 (3.3) | 65.7 (3.4) | 65.2 (4.0) | 64.5 (4.5) | 62.2 (6.0) | <0.001 |
| female (%) | 331 (100.0) | 79 (100.0) | 77 (100.0) | 55 (100.0) | 65 (100.0) | 55 (100.0) | NA |
| Race (%) | | | | | | | 0.261 |
| Chinese | 304 ( 92.4) | 76 ( 97.4) | 73 ( 94.8) | 48 ( 87.3) | 59 ( 92.2) | 48 ( 87.3) | |
| Indian | 14 ( 4.3) | 1 ( 1.3) | 2 ( 2.6) | 3 ( 5.5) | 3 ( 4.7) | 5 ( 9.1) | |
| Malay | 4 ( 1.2) | 0 ( 0.0) | 0 ( 0.0) | 1 ( 1.8) | 2 ( 3.1) | 1 ( 1.8) | |
| Others | 7 ( 2.1) | 1 ( 1.3) | 2 ( 2.6) | 3 ( 5.5) | 0 ( 0.0) | 1 ( 1.8) | |
| Body mass index , mean (SD), kg/m$^2$ | 24.5 (9.9) | 21.8 (2.2) | 21.7 (2.1) | 23.4 (2.2) | 29.1 (20.7) | 28.2 (3.7) | <0.001 |
| Waist circumference, mean (SD), cm | 81.1 (14.8) | 76.9 (10.6) | 75.7 (12.0) | 77.6 (13.7) | 84.9 (19.3) | 93.6 (9.2) | <0.001 |
| VAT area, mean (SD), cm$^2$ | 80.8 (40.7) | 49.9 (22.5) | 55.7 (22.5) | 76.8 (21.3) | 108.2 (33.4) | 132.1 (31.8) | <0.001 |
| TG, mean (SD), mmol/L | 1.8 (0.5) | 2.0 (0.4) | 2.0 (0.4) | 1.6 (0.4) | 1.7 (0.5) | 1.5 (0.3) | <0.001 |
| HDL-C , mean (SD), mmol/L | 1.2 (0.6) | 0.9 (0.3) | 0.9 (0.3) | 1.5 (0.6) | 1.3 (0.5) | 1.6 (0.7) | <0.001 |
| LDL-C, mean (SD), mmol/L | 3.4 (0.9) | 3.5 (1.0) | 3.3 (0.8) | 3.3 (1.0) | 3.5 (0.9) | 3.3 (1.0) | 0.373 |
| Lp(a), mean (SD) | 20.7 (25.1) | 22.2 (22.8) | 25.1 (29.7) | 24.3 (28.0) | 19.2 (24.5) | 10.8 (15.2) | 0.013 |
| FPG, mean (SD), mmol/L | 5.4 (1.4) | 4.9 (1.0) | 4.7 (0.4) | 6.3 (1.3) | 4.8 (0.4) | 6.8 (2.2) | <0.001 |
| HbA1c, mean (SD), % | 6.0 (0.9) | 5.7 (0.6) | 5.6 (0.3) | 6.7 (1.0) | 5.7 (0.3) | 6.9 (1.2) | <0.001 |
| Fasting insulin, mean (SD), mU/L | 10.9 (8.0) | 6.0 (2.5) | 6.2 (2.5) | 13.1 (6.6) | 11.2 (5.5) | 21.7 (10.2) | <0.001 |
| HOMA-IR, mean (SD) | 2.8 (2.8) | 1.3 (0.6) | 1.3 (0.6) | 3.7 (2.0) | 2.4 (1.2) | 6.7 (4.2) | <0.001 |
| SBP, mean (SD), mmHg | 133.5 (17.7) | 117.4 (8.7) | 146.4 (13.0) | 134.5 (14.5) | 136.4 (18.6) | 133.9 (17.6) | <0.001 |
| DBP, mean (SD), mmHg | 79.0 (9.6) | 71.8 (8.0) | 82.4 (8.9) | 79.5 (8.1) | 81.7 (9.7) | 80.8 (8.6) | <0.001 |
| Liver Fat by CAP, mean (SD), dB/m | 240.1 (53.0) | 205.7 (28.6) | 207.7 (38.4) | 250.2 (49.3) | 264.0 (43.3) | 296.5 (44.4) | <0.001 |
| hsCRP, mean (SD), mg/L | 1.4 (2.2) | 1.0 (2.4) | 1.0 (1.3) | 1.4 (1.6) | 1.8 (2.6) | 2.2 (2.6) | 0.005 |
| Uric acid, mean (SD), mmol/L | 0.29 (0.06) | 0.27 (0.05) | 0.27 (0.06) | 0.31 (0.06) | 0.30 (0.05) | 0.33 (0.07) | <0.001 |
| eGFR, mean (SD), mL/min/1.73m$^2$ | 87.6 (10.8) | 89.3 (8.2) | 87.5 (11.1) | 84.9 (10.8) | 89.9 (10.3) | 85.4 (13.3) | 0.030 |
| MoCA score , mean (SD) | 25.7 (2.6) | 25.5 (2.4) | 26.2 (2.1) | 26.0 (2.3) | 25.2 (3.3) | 25.6 (2.7) | 0.222 |
| Alcohol status (%) | | | | | | | 0.517 |

| | | | | | | | |
|---|---|---|---|---|---|---|---|
| Ex-drinker | 6 ( 1.9) | 1 ( 1.3) | 1 ( 1.3) | 1 ( 1.9) | 0 ( 0.0) | 3 ( 5.7) | |
| No | 226 ( 69.8) | 54 ( 70.1) | 54 ( 70.1) | 40 ( 75.5) | 45 ( 70.3) | 33 ( 62.3) | |
| Current drinker | 92 ( 28.4) | 22 ( 28.6) | 22 ( 28.6) | 12 ( 22.6) | 19 ( 29.7) | 17 ( 32.1) | |
| Smoking status (%) | | | | | | | 0.575 |
| Ex-smoker | 6 ( 1.9) | 3 ( 3.9) | 1 ( 1.3) | 0 ( 0.0) | 1 ( 1.6) | 1 ( 1.9) | |
| No | 314 ( 96.9) | 73 ( 94.8) | 76 ( 98.7) | 53 (100.0) | 61 ( 95.3) | 51 ( 96.2) | |
| Current smoker (%) | 4 ( 1.2) | 1 ( 1.3) | 0 ( 0.0) | 0 ( 0.0) | 2 ( 3.1) | 1 ( 1.9) | |
| Metabolic syndrome–IDF (%) | 96 ( 29.0) | 4 ( 5.1) | 6 ( 7.8) | 19 ( 34.5) | 26 ( 40.0) | 41 ( 74.5) | <0.001 |
| Obesity classification by BMI (%) | | | | | | | <0.001 |
| Underweight | 14 ( 4.2) | 7 ( 8.9) | 6 ( 7.8) | 1 ( 1.8) | 0 ( 0.0) | 0 ( 0.0) | |
| Normal weight | 134 ( 40.5) | 48 ( 60.8) | 52 ( 67.5) | 23 ( 41.8) | 7 ( 10.8) | 4 ( 7.3) | |
| Overweight | 131 ( 39.6) | 24 ( 30.4) | 19 ( 24.7) | 31 ( 56.4) | 37 ( 56.9) | 20 ( 36.4) | |
| Obesity | 52 ( 15.7) | 0 ( 0.0) | 0 ( 0.0) | 0 ( 0.0) | 21 ( 32.3) | 31 ( 56.4) | |
| Obesity (%) | 52 ( 15.7) | 0 ( 0.0) | 0 ( 0.0) | 0 ( 0.0) | 21 ( 32.3) | 31 ( 56.4) | <0.001 |
| Central Obesity by WC (%) | 189 ( 57.1) | 26 ( 32.9) | 28 ( 36.4) | 22 ( 40.0) | 58 ( 89.2) | 55 (100.0) | <0.001 |
| Visceral adiposity by VAT (%) | 97 ( 29.3) | 0 ( 0.0) | 0 ( 0.0) | 6 ( 10.9) | 40 ( 61.5) | 51 ( 92.7) | <0.001 |
| low HDL (%) | 39 ( 11.8) | 3 ( 3.8) | 0 ( 0.0) | 9 ( 16.4) | 9 ( 13.8) | 18 ( 32.7) | <0.001 |
| high TG (%) | 58 ( 17.5) | 4 ( 5.1) | 0 ( 0.0) | 20 ( 36.4) | 13 ( 20.0) | 21 ( 38.2) | <0.001 |
| high LDL (%) | 161 ( 48.6) | 42 ( 53.2) | 33 ( 42.9) | 24 ( 43.6) | 41 ( 63.1) | 21 ( 38.2) | 0.039 |
| high FPG (%) | 82 ( 24.8) | 7 ( 8.9) | 1 ( 1.3) | 39 ( 70.9) | 0 ( 0.0) | 35 ( 63.6) | <0.001 |
| high HbA1c (%) | 211 ( 63.7) | 39 ( 49.4) | 30 ( 39.0) | 53 ( 96.4) | 35 ( 53.8) | 54 ( 98.2) | <0.001 |
| IR by HOMA-IR(%) | 121 ( 36.6) | 1 ( 1.3) | 1 ( 1.3) | 41 ( 74.5) | 23 ( 35.4) | 55 (100.0) | <0.001 |
| Diabetes status (%) | | | | | | | <0.001 |
| Normorglycemia | 116 ( 35.0) | 39 ( 49.4) | 45 ( 58.4) | 1 ( 1.8) | 30 ( 46.2) | 1 ( 1.8) | |
| Pre DM | 145 ( 43.8) | 31 ( 39.2) | 30 ( 39.0) | 29 ( 52.7) | 32 ( 49.2) | 23 ( 41.8) | |
| DM | 70 ( 21.1) | 9 ( 11.4) | 2 ( 2.6) | 25 ( 45.5) | 3 ( 4.6) | 31 ( 56.4) | |
| high SBP (%) | 183 ( 55.3) | 0 ( 0.0) | 77 (100.0) | 36 ( 65.5) | 42 ( 64.6) | 28 ( 50.9) | <0.001 |
| high DBP (%) | 87 ( 26.3) | 2 ( 2.5) | 29 ( 37.7) | 16 ( 29.1) | 23 ( 35.4) | 17 ( 30.9) | <0.001 |
| Hypertension (%) | 206 ( 62.2) | 9 ( 11.4) | 77 (100.0) | 41 ( 74.5) | 42 ( 64.6) | 37 ( 67.3) | <0.001 |

| | | | | | | | |
|---|---|---|---|---|---|---|---|
| Dyslipidemia (%) | 233 ( 70.4) | 53 ( 67.1) | 42 ( 54.5) | 42 ( 76.4) | 54 ( 83.1) | 42 ( 76.4) | 0.002 |
| Liver Steatosis (%) | 123 ( 37.2) | 5 ( 6.3) | 9 ( 11.7) | 21 ( 38.2) | 43 ( 66.2) | 45 ( 81.8) | <0.001 |
| high hsCRP (%) | 110 ( 33.2) | 11 ( 13.9) | 21 ( 27.3) | 19 ( 34.5) | 28 ( 43.1) | 31 ( 56.4) | <0.001 |
| Hyperuricemia (%) | 49 ( 14.8) | 3 ( 3.8) | 6 ( 7.8) | 16 ( 29.1) | 8 ( 12.3) | 16 ( 29.1) | <0.001 |
| MoCA Cognitive impairment (%) | 120 ( 39.2) | 35 ( 46.1) | 24 ( 33.3) | 12 ( 26.1) | 26 ( 42.6) | 23 ( 45.1) | 0.141 |
| Anti-diabetic drug (%) | 26 ( 7.9) | 6 ( 7.6) | 1 ( 1.3) | 8 ( 14.5) | 3 ( 4.6) | 8 ( 14.5) | 0.015 |
| Anti-hypertension drug (%) | 61 ( 18.4) | 7 ( 8.9) | 13 ( 16.9) | 9 ( 16.4) | 14 ( 21.5) | 18 ( 32.7) | 0.011 |
| Lipid-lowering drug (%) | 60 ( 18.1) | 12 ( 15.2) | 12 ( 15.6) | 14 ( 25.5) | 16 ( 24.6) | 6 ( 10.9) | 0.161 |
| Metabolic health–WC groups (%) | | | | | | | <0.001 |
| MHNWC | 98 ( 29.6) | 45 ( 57.0) | 41 ( 53.2) | 9 ( 16.4) | 3 ( 4.6) | 0 ( 0.0) | |
| MHHWC | 93 ( 28.1) | 22 ( 27.8) | 22 ( 28.6) | 3 ( 5.5) | 32 ( 49.2) | 14 ( 25.5) | |
| MUHNWC | 44 ( 13.3) | 8 ( 10.1) | 8 ( 10.4) | 24 ( 43.6) | 4 ( 6.2) | 0 ( 0.0) | |
| MUHHWC | 96 ( 29.0) | 4 ( 5.1) | 6 ( 7.8) | 19 ( 34.5) | 26 ( 40.0) | 41 ( 74.5) | |
| Metabolic health–BMI groups (%) | | | | | | | <0.001 |
| MHNW | 171 ( 51.7) | 67 ( 84.8) | 63 ( 81.8) | 12 ( 21.8) | 24 ( 36.9) | 5 ( 9.1) | |
| MHO | 20 ( 6.0) | 0 ( 0.0) | 0 ( 0.0) | 0 ( 0.0) | 11 ( 16.9) | 9 ( 16.4) | |
| MUNW | 108 ( 32.6) | 12 ( 15.2) | 14 ( 18.2) | 43 ( 78.2) | 20 ( 30.8) | 19 ( 34.5) | |
| MUO | 32 ( 9.7) | 0 ( 0.0) | 0 ( 0.0) | 0 ( 0.0) | 10 ( 15.4) | 22 ( 40.0) | |

Abbreviation: BMI, body mass index; WC, Waist circumference; VAT, Visceral adiposity tissue; TG, Triglyceride; LDL-C, Low density lipoprotein cholesterol; HDL-C, High density lipoprotein cholesterol; FPG, Fasting plasma glucose; HbA1c, Glycated Hemoglobin A1c; HOMA-IR, Homeostatic Model Assessment of Insulin Resistance; CAP, Controlled attenuation parameter (Db/m); hsCRP, High Sensitivity C-reactive Protein; SBP, systolic blood pressure; DBP, diastolic blood pressure; MHNW, Metabolically healthy normal weight; MUNW, Metabolically unhealthy normal weight; MHO, Metabolically healthy obese; MUO, Metabolically unhealthy obese; MHNWC, Metabolically healthy normal waist circumference; MUHWC, Metabolically unhealthy high waist circumference

**Table S8 Prevalence of metabolic risk factors and 5 latent classes according to fasting glucose in discovery cohort–RESET study**

| | **Overall** | **Normoglycemia** | **FPG ≥ 5.6** | **P** |
|---|---|---|---|---|
| | 1034 | 790 | 244 | |
| Obesity (%) | 198 (19.1) | 129 (16.3) | 69 ( 28.3) | <0.001 |
| Central Obesity by WC (%) | 461 (44.6) | 318 (40.3) | 143 ( 58.6) | <0.001 |
| Visceral adiposity by VAT (%) | 386 (37.3) | 253 (32.0) | 133 ( 54.5) | <0.001 |
| low HDL (%) | 85 ( 8.2) | 47 ( 5.9) | 38 ( 15.6) | <0.001 |
| high TG (%) | 178 (17.2) | 118 (14.9) | 60 ( 24.6) | 0.001 |
| high LDL (%) | 453 (43.8) | 380 (48.1) | 73 ( 29.9) | <0.001 |
| high FPG (%) | 244 (23.6) | 0 ( 0.0) | 244 (100.0) | <0.001 |
| **high HbA1c (%)** | **652 (63.1)** | **426 (53.9)** | **226 ( 92.6)** | **<0.001** |
| **IR by HOMA-IR(%)** | **376 (36.4)** | **203 (25.7)** | **173 ( 70.9)** | **<0.001** |
| **high HbA1c or IR (%)** | **727 (70.3)** | **491 (62.2)** | **236 ( 96.7)** | **<0.001** |
| high SBP (%) | 563 (54.4) | 431 (54.6) | 132 ( 54.1) | 0.958 |
| high DBP (%) | 361 (34.9) | 285 (36.1) | 76 ( 31.1) | 0.182 |
| Liver Steatosis (%) | 392 (37.9) | 264 (33.4) | 128 ( 52.5) | <0.001 |
| high hsCRP (%) | 312 (30.2) | 229 (29.0) | 83 ( 34.0) | 0.157 |
| Hyperuricemia (%) | 174 (16.8) | 121 (15.3) | 53 ( 21.7) | 0.025 |
| Latent class (%) | | | | <0.001 |
| Class 1 | 244 (23.6) | 220 (27.8) | 24 ( 9.8) | |
| Class 2 | 244 (23.6) | 234 (29.6) | 10 ( 4.1) | |
| Class 3 | 140 (13.5) | 43 ( 5.4) | 97 ( 39.8) | |
| Class 4 | 211 (20.4) | 211 (26.7) | 0 ( 0.0) | |
| Class 5 | 195 (18.9) | 82 (10.4) | 113 ( 46.3) | |

WC, Waist circumference; VAT, Visceral adiposity tissue; TG, Triglyceride; LDL-C, Low density lipoprotein cholesterol; HDL-C, High density lipoprotein cholesterol; FPG, Fasting plasma glucose; HbA1c, Glycated Hemoglobin A1c; HOMA-IR, Homeostatic Model Assessment of Insulin Resistance; hsCRP, High Sensitivity C-reactive Protein; SBP, systolic blood pressure; DBP, diastolic blood pressure;

**Table S9 Latent class analysis (LCA) model using simplified metabolic risk factors metrics for determining best classes**

| Classes | logLik | AIC | BIC | Gsq | Chisq |
|---|---|---|---|---|---|
| 2 | -3754.1581 | 7538.31611 | 7612.43397 | 352.677715 | 344.302035 |
| **3** | **-3659.2684** | **7364.5368** | **7478.18417** | **162.898396** | **184.460842** |
| 4 | -3635.6173 | 7333.23452 | 7486.41141 | 115.596123 | 132.105936 |
| 5 | -3624.0802 | 7326.16035 | 7518.86677 | 92.5219544 | 83.3984892 |
| 6 | -3616.5009 | 7327.0017 | 7559.23763 | 77.3633026 | 73.0023379 |

logLik = log-likelihood; AIC = Akaike Information Criterion; BIC = Bayesian Information Criterion; Gsq = Likelihood ratio chi-square statistic ($G^2$); Chisq = Pearson chi-square statistic

**Table S10 Sensitivity analyses to vary each binary threshold by ±5% to assess the latent classification compared with original binary threshold**

| Variable | Measure (unit) | Sex | Rule | df_minus (×0.95) | Base (×1.00) | df_plus (×1.05) |
|---|---|---|---|---|---|---|
| Obesity | BMI (kg/m2) | All | ≥ | 26.12 | 27.5 | 28.88 |
| Central obesity | Waist (cm) | Female | ≥ | 76 | 80 | 84 |
| Central obesity | Waist (cm) | Male | ≥ | 85.5 | 90 | 94.5 |
| Visceral adiposity | VAT (cm2) | All | ≥ | 95 | 100 | 105 |
| High TG | Triglycerides (mmol/L) | All | ≥ | 1.615 | 1.7 | 1.785 |
| High LDL | LDL-C (mmol/L) | All | ≥ | 3.23 | 3.4 | 3.57 |
| Low HDL | HDL-C (mmol/L) | Female | < | 1.358 | 1.29 | 1.229 |
| Low HDL | HDL-C (mmol/L) | Male | < | 1.084 | 1.03 | 0.981 |
| High Lp(a) | Lp(a) (mg/dL) | All | ≥ | 47.5 | 50 | 52.5 |
| High FPG | Fasting glucose (mmol/L) | All | ≥ | 5.32 | 5.6 | 5.88 |
| High HbA1c | HbA1c (%) | All | ≥ | 5.415 | 5.7 | 5.985 |
| Insulin resistance | HOMA-IR | All | ≥ | 2.375 | 2.5 | 2.625 |
| High hsCRP | hsCRP (mg/L) | All | ≥ | 0.95 | 1 | 1.05 |
| Hyperuricaemia | Uric acid (mmol/L) | Female | ≥ | 0.342 | 0.36 | 0.378 |
| Hyperuricaemia | Uric acid (mmol/L) | Male | ≥ | 0.399 | 0.42 | 0.441 |
| Liver steatosis | CAP (dB/m) | All | ≥ | 241.3 | 254 | 266.7 |
| High SBP | SBP (mmHg) | All | ≥ | 123.5 | 130 | 136.5 |
| High DBP | DBP (mmHg) | All | ≥ | 80.75 | 85 | 89.25 |

| Class | Base_n | New_n | Stability_within | Outflow_within | Inflow_overall | Outflow_overall | Net_change_overall | Binary Threshold Change |
|---|---|---|---|---|---|---|---|---|
| 1 | 244 | 171 | 0.62 | 0.38 | 0.02 | 0.09 | -0.07 | -5% |
| 2 | 244 | 343 | 0.94 | 0.06 | 0.11 | 0.01 | 0.10 | -5% |
| 3 | 140 | 168 | 0.66 | 0.34 | 0.07 | 0.05 | 0.03 | -5% |
| 4 | 211 | 180 | 0.68 | 0.32 | 0.04 | 0.07 | -0.03 | -5% |
| 5 | 195 | 172 | 0.74 | 0.26 | 0.03 | 0.05 | -0.02 | -5% |
| 1 | 244 | 482 | 0.94 | 0.06 | 0.24 | 0.01 | 0.23 | +5% |
| 2 | 244 | 83 | 0.23 | 0.77 | 0.03 | 0.18 | -0.16 | +5% |
| 3 | 140 | 146 | 0.55 | 0.45 | 0.07 | 0.06 | 0.01 | +5% |
| 4 | 211 | 219 | 0.66 | 0.34 | 0.08 | 0.07 | 0.01 | +5% |
| 5 | 195 | 104 | 0.52 | 0.48 | 0.00 | 0.09 | -0.09 | +5% |

The reclassification rates for five latent classes derived from the binary variables according to the original cut-off and ±5% cut-off.

**Table S11 Sensitivity analysis employed Latent profile analysis (LPA) on continuous data**

| **Variable** | Class 1 | Class 2 | Class 3 | Class 4 | Class 5 | p |
|---|---|---|---|---|---|---|
| n | 228 | 301 | 97 | 226 | 182 | |
| Age , mean (SD), y | 64.0 (4.6) | 63.3 (4.4) | 63.6 (5.3) | 62.5 (5.2) | 61.4 (6.9) | <0.001 |
| Male (%) | 131 (57.5) | 211 (70.1) | 58 (59.8) | 186 (82.3) | 117 (64.3) | <0.001 |
| **Body mass index , mean (SD), kg/m2** | 21.2 (1.8) | 23.6 (2.0) | 23.5 (2.0) | 26.8 (2.2) | 29.8 (12.8) | <0.001 |
| **Waist circumference, mean (SD), cm** | 74.1 (11.6) | 83.6 (5.8) | 83.5 (6.0) | 92.3 (5.7) | 88.0 (27.9) | <0.001 |
| **VAT area, mean (SD), cm2** | 50.5 (16.0) | 79.0 (19.8) | 82.4 (22.4) | 118.1 (26.3) | 141.8 (47.5) | <0.001 |
| **TG, mean (SD), mmol/L** | 2.0 (0.4) | 1.6 (0.3) | 1.6 (0.5) | 1.4 (0.3) | 1.3 (0.3) | <0.001 |
| **HDL-C , mean (SD), mmol/L** | 0.8 (0.2) | 1.1 (0.4) | 1.1 (0.3) | 1.5 (0.9) | 1.6 (0.8) | <0.001 |
| **LDL-C, mean (SD), mmol/L** | 3.4 (0.9) | 3.5 (1.0) | 2.6 (0.7) | 3.3 (0.9) | 3.0 (1.0) | <0.001 |
| **FPG, mean (SD), mmol/L** | 4.8 (0.4) | 4.8 (0.4) | 7.0 (2.1) | 5.1 (0.5) | 6.4 (2.0) | <0.001 |
| **HbA1c, mean (SD), %** | 5.6 (0.3) | 5.6 (0.4) | 7.0 (1.1) | 5.8 (0.3) | 6.7 (1.3) | <0.001 |
| **HOMA-IR, mean (SD)** | 1.1 (0.5) | 1.6 (0.6) | 3.2 (2.2) | 3.0 (1.3) | 6.1 (3.8) | <0.001 |
| **SBP, mean (SD), mmHg** | 128.3 (17.2) | 133.4 (15.6) | 130.3 (17.6) | 133.3 (15.8) | 134.5 (14.5) | <0.001 |
| **DBP, mean (SD), mmHg** | 77.7 (10.1) | 82.4 (8.8) | 78.3 (7.8) | 84.5 (8.2) | 83.7 (9.1) | <0.001 |
| **Liver Fat by CAP, mean (SD), dB/m** | 200.3 (31.0) | 226.3 (38.3) | 242.1 (56.2) | 266.5 (40.5) | 290.0 (48.5) | <0.001 |
| **hsCRP, mean (SD), mg/L** | 0.4 (0.2) | 1.8 (3.0) | 0.5 (0.3) | 0.8 (0.5) | 2.3 (2.8) | <0.001 |
| **Uric acid, mean (SD), mmol/L** | 0.3 (0.1) | 0.3 (0.1) | 0.3 (0.1) | 0.4 (0.1) | 0.4 (0.1) | <0.001 |
| Obesity (%) | 0 ( 0.0) | 7 ( 2.3) | 4 ( 4.1) | 80 (35.4) | 107 (58.8) | <0.001 |
| Central Obesity by WC (%) | 19 ( 8.3) | 93 (30.9) | 31 (32.0) | 175 (77.4) | 143 (78.6) | <0.001 |
| Visceral adiposity (%) | 0 ( 0.0) | 44 (14.6) | 21 (21.6) | 175 (77.4) | 146 (80.2) | <0.001 |
| low HDL (%) | 1 ( 0.4) | 8 ( 2.7) | 6 ( 6.2) | 26 (11.5) | 44 (24.2) | <0.001 |
| high TG (%) | 0 ( 0.0) | 29 ( 9.6) | 5 ( 5.2) | 74 (32.7) | 70 (38.5) | <0.001 |
| high LDL (%) | 116 (50.9) | 165 (54.8) | 15 (15.5) | 101 (44.7) | 56 (30.8) | <0.001 |
| high FPG (%) | 10 ( 4.4) | 14 ( 4.7) | 80 (82.5) | 33 (14.6) | 107 (58.8) | <0.001 |
| high HbA1c (%) | 89 (39.0) | 162 (53.8) | 89 (91.8) | 159 (70.4) | 153 (84.1) | <0.001 |
| IR by HOMA-IR(%) | 3 ( 1.3) | 27 ( 9.0) | 52 (53.6) | 138 (61.1) | 156 (85.7) | <0.001 |
| high SBP (%) | 105 (46.1) | 179 (59.5) | 43 (44.3) | 124 (54.9) | 112 (61.5) | 0.002 |
| high DBP (%) | 52 (22.8) | 114 (37.9) | 18 (18.6) | 103 (45.6) | 74 (40.7) | <0.001 |
| Liver Steatosis (%) | 12 ( 5.3) | 68 (22.6) | 36 (37.1) | 139 (61.5) | 137 (75.3) | <0.001 |
| high hsCRP (%) | 7 ( 3.1) | 127 (42.2) | 7 ( 7.2) | 62 (27.4) | 109 (59.9) | <0.001 |
| Hyperuricemia (%) | 14 ( 6.1) | 44 (14.6) | 13 (13.4) | 52 (23.0) | 51 (28.0) | <0.001 |

Latent profile analysis (LPA) on continuous data was performed to identify whether distinct phenotypes would be obtained independent of binary diagnostic cut-offs. Different LPA models using equal, zero or varying variances and covariances were compared according to the lowest BIC. The best LPA model was set as Varying variances, zero covariances. We manually choose 5 classes for LPA. The results showed that latent phenotype such as Class 3 and Class 4 still exists when using continuous variables, specifically, Class 3 represents lean but hyperglycemia and Class 4 represents obese but relatively normoglycemia. However the HOMA-IR did not show significant difference between these two classes.

**Table S12 Sensitivity analysis employed HCPC on continuous data clustering**

| **Variable** | Class 1 | Class 2 | Class 3 | Class 4 | p |
|---|---|---|---|---|---|
| n | 344 | 428 | 124 | 138 | |
| Age , mean (SD), y | 64.2 (4.4) | 62.8 (4.8) | 62.6 (6.2) | 60.8 (6.8) | <0.001 |
| Male (%) | 185 (53.8) | 336 (78.5) | 73 ( 58.9) | 109 (79.0) | <0.001 |
| **Body mass index , mean (SD), kg/m2** | 21.7 (2.1) | 25.0 (2.2) | 25.8 (3.2) | 31.4 (14.3) | <0.001 |
| **Waist circumference, mean (SD), cm** | 74.8 (14.0) | 85.4 (12.3) | 88.4 (12.3) | 99.7 (10.9) | <0.001 |
| **VAT area, mean (SD), cm2** | 57.5 (20.4) | 95.5 (25.1) | 113.1 (40.2) | 152.9 (43.2) | <0.001 |
| **TG, mean (SD), mmol/L** | 1.9 (0.4) | 1.5 (0.3) | 1.4 (0.3) | 1.3 (0.3) | <0.001 |
| **HDL-C , mean (SD), mmol/L** | 0.9 (0.3) | 1.3 (0.7) | 1.5 (0.8) | 1.7 (0.8) | <0.001 |
| **LDL-C, mean (SD), mmol/L** | 3.3 (0.9) | 3.4 (1.0) | 2.7 (0.9) | 3.1 (1.0) | <0.001 |
| **FPG, mean (SD), mmol/L** | 4.9 (0.5) | 5.0 (0.5) | 8.0 (2.3) | 5.3 (0.6) | <0.001 |
| **HbA1c, mean (SD), %** | 5.6 (0.4) | 5.8 (0.4) | 7.7 (1.3) | 6.0 (0.5) | <0.001 |
| **HOMA-IR, mean (SD)** | 1.3 (0.7) | 2.3 (1.2) | 6.0 (4.3) | 4.9 (2.5) | <0.001 |
| **SBP, mean (SD), mmHg** | 127.1 (15.9) | 136.4 (16.4) | 130.7 (14.0) | 133.1 (14.1) | <0.001 |
| **DBP, mean (SD), mmHg** | 77.0 (8.9) | 84.9 (8.3) | 79.9 (8.4) | 84.8 (8.9) | <0.001 |
| **Liver Fat by CAP, mean (SD), dB/m** | 203.2 (36.3) | 246.9 (41.5) | 275.7 (51.6) | 293.8 (40.6) | <0.001 |
| **hsCRP, mean (SD), mg/L** | 0.9 (2.1) | 1.2 (1.9) | 1.2 (2.1) | 2.2 (2.6) | <0.001 |
| **Uric acid, mean (SD), mmol/L** | 0.3 (0.1) | 0.4 (0.1) | 0.3 (0.1) | 0.4 (0.1) | <0.001 |
| Obesity (%) | 2 ( 0.6) | 55 (12.9) | 35 ( 28.2) | 106 (76.8) | <0.001 |
| Central Obesity by WC (%) | 53 (15.4) | 197 (46.0) | 79 ( 63.7) | 132 (95.7) | <0.001 |
| Visceral adiposity (%) | 9 ( 2.6) | 180 (42.1) | 72 ( 58.1) | 125 (90.6) | <0.001 |
| low HDL (%) | 3 ( 0.9) | 29 ( 6.8) | 27 ( 21.8) | 26 (18.8) | <0.001 |
| high TG (%) | 11 ( 3.2) | 78 (18.2) | 37 ( 29.8) | 52 (37.7) | <0.001 |
| high LDL (%) | 168 (48.8) | 216 (50.5) | 24 ( 19.4) | 45 (32.6) | <0.001 |
| high FPG (%) | 25 ( 7.3) | 59 (13.8) | 122 ( 98.4) | 38 (27.5) | <0.001 |
| high HbA1c (%) | 157 (45.6) | 263 (61.4) | 124 (100.0) | 108 (78.3) | <0.001 |
| IR by HOMA-IR(%) | 22 ( 6.4) | 138 (32.2) | 99 ( 79.8) | 117 (84.8) | <0.001 |
| high SBP (%) | 141 (41.0) | 280 (65.4) | 64 ( 51.6) | 78 (56.5) | <0.001 |
| high DBP (%) | 56 (16.3) | 213 (49.8) | 30 ( 24.2) | 62 (44.9) | <0.001 |
| Liver Steatosis (%) | 24 ( 7.0) | 176 (41.1) | 78 ( 62.9) | 114 (82.6) | <0.001 |
| high hsCRP (%) | 65 (18.9) | 128 (29.9) | 40 ( 32.3) | 79 (57.2) | <0.001 |
| Hyperuricemia (%) | 23 ( 6.7) | 79 (18.5) | 26 ( 21.0) | 46 (33.3) | <0.001 |

For continuous variables we performed unsupervised clustering using Hierarchical Clustering on Principal Components (HCPC) on continuous data to identify groupings independent of binary diagnostic cut-offs. 14 continuous cardiometabolic features (Bold). All variables were Z-standardized, and Principal Component Analysis (PCA) was performed to reduce dimensionality, retaining the top ten principal components sufficient to explain ~90% of the variance. The final optimal cluster numbers: k=4 was derived from the hierarchical tree and consolidated via K-means to optimize stability. Analysis was performed using the FactoMineR package

**Table S13 The predictive accuracy for the decision tree model in identifying unsupervised latent classes with 5-fold internal cross-validation**

| | Class: Class_1 | Class: Class_2 | Class: Class_3 | Class: Class_4 | Class: Class_5 |
|---|---|---|---|---|---|
| Sensitivity | 0.920948617 | 0.882783883 | 0.606060606 | 0.826347305 | 0.83732057 |
| Specificity | 0.985915493 | 0.996057819 | 0.933481153 | 0.915801615 | 0.97575758 |
| Pos Pred Value | 0.954918033 | 0.987704918 | 0.571428571 | 0.654028436 | 0.8974359 |
| Neg Pred Value | 0.974683544 | 0.959493671 | 0.941834452 | 0.964763062 | 0.95947557 |
| Precision | 0.954918033 | 0.987704918 | 0.571428571 | 0.654028436 | 0.8974359 |
| Recall | 0.920948617 | 0.882783883 | 0.606060606 | 0.826347305 | 0.83732057 |
| F1 | 0.937625755 | 0.932301741 | 0.588235294 | 0.73015873 | 0.86633663 |
| Prevalence | 0.244680851 | 0.264023211 | 0.127659574 | 0.161508704 | 0.20212766 |
| Detection Rate | 0.225338491 | 0.233075435 | 0.077369439 | 0.133462282 | 0.16924565 |
| Detection Prevalence | 0.235976789 | 0.235976789 | 0.135396518 | 0.204061896 | 0.18858801 |
| Balanced Accuracy | 0.953432055 | 0.939420851 | 0.76977088 | 0.87107446 | 0.90653907 |

The overall predictive accuracy for the decision tree model in identifying unsupervised latent classes was measured by an area under the curve (AUC) of 0.84, 95% CI:(0.81, 0.86) with 5-fold internal cross-validation.

**Table S14 Biological annotation of the 31 DEPs with |$\log_2$ fold-change| > 0.8 in PICMAN across these two phenotypes**

| Variable | Module | Full name | Tissue / main source | Biological function | category | logFC_LeanIR | logFC_ObeseIS | P.Value_LeanIR | P.Value_ObeseIS | validated_ukb |
|---|---|---|---|---|---|---|---|---|---|---|
| **ADH1B** | Hepatic metabolism & injury | Alcohol dehydrogenase 1B (class I) | Liver | NAD-dependent ethanol/alcohol oxidation | Common_Same | 0.613 | 0.951 | 1.47e-02 | 4.92e-05 | TRUE |
| **ADH4** | Hepatic metabolism & injury | Alcohol dehydrogenase 4 (class II) | Liver | NAD-dependent alcohol/retinol oxidation | Unique_ObeseIS | | 1.191 | | 3.65e-05 | TRUE |
| **CA5A** | Hepatic metabolism & injury | Carbonic anhydrase 5A (mitochondrial) | Liver (mitochondria) | Supplies bicarbonate for gluconeogenesis, ureagenesis and lipogenesis | Unique_ObeseIS | | 1.051 | | 1.41e-04 | TRUE |
| **DCXR** | Hepatic metabolism & injury | Dicarbonyl / L-xylulose reductase | Liver, kidney | Carbonyl and glucose (uronate-cycle) metabolism; osmoregulation | Unique_ObeseIS | | 0.817 | | 2.81e-04 | TRUE |
| **FTCD** | Hepatic metabolism & injury | Formimidoyltransferase cyclodeaminase | Liver | Histidine catabolism and folate metabolism | Common_Same | 0.724 | 0.961 | 2.24e-02 | 1.02e-03 | TRUE |
| **GSTA1** | Hepatic metabolism & injury | Glutathione S-transferase alpha 1 | Liver | Phase-II detoxification; glutathione conjugation of electrophiles | Common_Same | 0.751 | 1.079 | 8.28e-03 | 4.58e-05 | TRUE |
| **HAO1** | Hepatic metabolism & injury | Hydroxyacid oxidase 1 (glycolate oxidase) | Liver (peroxisomal) | Oxidises glycolate to glyoxylate; oxalate metabolism | Common_Same | 0.915 | 1.313 | 3.18e-02 | 8.57e-04 | TRUE |
| **KRT18** | Hepatic metabolism & injury | Keratin 18 | Epithelia incl. hepatocytes | Intermediate filament; hepatocyte injury marker (M30 fragment) | Unique_ObeseIS | | 0.837 | | 1.67e-03 | TRUE |
| **KRT8** | Hepatic metabolism & injury | Keratin 8 | Epithelia incl. hepatocytes | Intermediate filament; hepatocyte injury/turnover marker | Unique_ObeseIS | | 0.98 | | 8.06e-03 | TRUE |
| **IL6** | Inflammatory signaling | Interleukin-6 | Immune cells, adipose, muscle | Pleiotropic pro-inflammatory cytokine; acute-phase response | Unique_LeanIR | 0.863 | | 1.06e-02 | | TRUE |
| **PLCB2** | Inflammatory signaling | Phospholipase C beta 2 | Haematopoietic / immune cells | GPCR-downstream signalling (IP3/DAG); leukocyte activation, chemotaxis | Unique_LeanIR | 0.959 | | 1.78e-02 | | TRUE |
| **SERPINE1** | Inflammatory signaling | Serpin family E member 1 (PAI-1) | Adipose, endothelium, liver | Anti-fibrinolytic; pro-thrombotic/pro-inflammatory; adipokine | Common_Same | 0.855 | 0.776 | 3.34e-03 | 3.50e-03 | TRUE |
| **FGF21** | Insulin & endocrine signaling | Fibroblast growth factor 21 | Liver (primary), adipose | Metabolic hormone (hepatokine); insulin-sensitising; FGF21 resistance | Common_Same | 1.193 | 0.791 | 1.19e-03 | 1.73e-02 | TRUE |

| | | | | | | | | | | |
|---|---|---|---|---|---|---|---|---|---|---|
| **IGFBP1** | Insulin & endocrine signaling | Insulin-like growth factor-binding protein 1 | Liver | Insulin-suppressed IGF carrier; marker of insulin action/fasting | Common_Same | -1.416 | -1.483 | 1.18e-03 | 2.16e-04 | TRUE |
| **LEP** | Insulin & endocrine signaling | Leptin | Adipose tissue | Adipokine; regulates energy balance/appetite; leptin resistance in obesity | Common_Same | 1.438 | 0.828 | 3.86e-04 | 2.27e-02 | TRUE |
| **AIFM1** | Mitochondrial & energy metabolism | Apoptosis-inducing factor, mitochondria-associated 1 | Mitochondria, ubiquitous | Mitochondrial oxidoreductase; complex-I/redox; caspase-independent apoptosis | Common_Same | 0.623 | 0.806 | 1.71e-02 | 8.28e-04 | TRUE |
| **ECHDC3** | Mitochondrial & energy metabolism | Enoyl-CoA hydratase domain-containing 3 | Mitochondria (broad; adipose/liver) | Putative fatty-acid/lipid metabolism | Unique_ObeseIS | | 0.912 | | 1.19e-02 | TRUE |
| **ECHS1** | Mitochondrial & energy metabolism | Enoyl-CoA hydratase, short chain 1 | Mitochondria (liver, muscle) | Fatty-acid beta-oxidation; branched-chain (valine) amino-acid catabolism | Common_Same | 0.697 | 0.868 | 4.28e-02 | 6.00e-03 | TRUE |
| **RTN4IP1** | Mitochondrial & energy metabolism | Reticulon-4-interacting protein 1 | Mitochondria (broad) | Mitochondrial NADPH oxidoreductase | Unique_LeanIR | 0.819 | | 3.39e-03 | | TRUE |
| **EIF4E** | Nucleic-acid & protein regulation | Eukaryotic translation initiation factor 4E | Ubiquitous | mRNA 5' cap binding; cap-dependent translation initiation | Unique_ObeseIS | | 0.909 | | 4.31e-02 | TRUE |
| **ELOA** | Nucleic-acid & protein regulation | Elongin A (TCEB3) | Ubiquitous | RNA-Pol-II transcription elongation factor | Unique_ObeseIS | | 0.89 | | 1.27e-05 | TRUE |
| **GTPBP2** | Nucleic-acid & protein regulation | GTP-binding protein 2 | Ubiquitous | Ribosome rescue / translational quality control; GTPase | Common_Same | 0.829 | 1.032 | 3.05e-02 | 3.41e-03 | TRUE |
| **ITPA** | Nucleic-acid & protein regulation | Inosine triphosphate pyrophosphatase | Ubiquitous | Nucleotide-pool sanitisation (hydrolyses ITP/dITP) | Unique_ObeseIS | | 0.849 | | 2.15e-02 | TRUE |
| **PARP1** | Nucleic-acid & protein regulation | Poly(ADP-ribose) polymerase 1 | Ubiquitous (nuclear) | DNA-damage repair via PARylation; NAD+ metabolism | Unique_ObeseIS | | 0.909 | | 1.26e-03 | TRUE |
| **ESYT2** | Cytoskeletal & structural | Extended synaptotagmin 2 | Ubiquitous | ER–plasma-membrane contact sites; Ca2+-dependent lipid transfer | Unique_LeanIR | 1.165 | | 7.40e-03 | | TRUE |
| **GAS2** | Cytoskeletal & structural | Growth arrest-specific 2 | Broad | Actin/microtubule cytoskeleton; cell-cycle and apoptosis regulation | Unique_ObeseIS | | 0.93 | | 3.37e-03 | TRUE |
| **IGSF9** | Cytoskeletal & structural | Immunoglobulin superfamily member 9 | Neural (broad) | Cell-adhesion molecule; synapse/neurite development | Common_Same | 0.841 | 1.013 | 1.26e-03 | 2.73e-05 | TRUE |

| | | | | | | | | | | |
|---|---|---|---|---|---|---|---|---|---|---|
| **OXT** | Insulin & endocrine signaling | Oxytocin | Hypothalamus (posterior pituitary) | Neuropeptide hormone; emerging metabolic/appetite roles | Unique_ObeseIS | | 1.031 | | 1.55e-02 | TRUE |
| **DBH** | Other / unclassified | Dopamine beta-hydroxylase | Adrenal medulla, sympathetic neurons | Converts dopamine to noradrenaline; catecholamine synthesis | Unique_ObeseIS | | -0.979 | | 1.87e-02 | TRUE |
| **DKKL1** | Other / unclassified | Dickkopf-like protein 1 (SGY-1) | Testis (broad) | Secreted Wnt-related modulator; spermatogenesis | Unique_ObeseIS | | -0.896 | | 3.75e-03 | TRUE |
| **TREH** | Other / unclassified | Trehalase | Intestine, kidney (brush border), liver | Hydrolyses trehalose to glucose; carbohydrate digestion | Unique_LeanIR | 1.04 | | 2.20e-02 | | TRUE |

**Table S15 The Dominant Biological Modules, Pathways and Proteins with top fold change in each class in PICMAN study**

| Class | Dominant Biological Module | Pathway | Source | Proteins with \|log₂ fold-change\| >0.8 |
|---|---|---|---|---|
| Isolated Hypertension | Innate immune activation and inflammatory signaling | NOD-like receptor signaling pathway | KEGG | TANK/NEK7/PLCB2 |
| Isolated Hypertension | Innate immune activation and inflammatory signaling | response to virus | GO | IFIT1/TANK |
| Lean IR/ Hyperglycemia | Endocrine–inflammatory metabolic stress signaling | Cytokine-cytokine receptor interaction | KEGG | LEP/IL6 |
| Lean IR/ Hyperglycemia | Endocrine–inflammatory metabolic stress signaling | AGE-RAGE signaling pathway in diabetic complications | KEGG | PLCB2/IL6/SERPINE1 |
| Lean IR/ Hyperglycemia | Endocrine–inflammatory metabolic stress signaling | lipid transport | GO | LEP/SLC4A1/ESYT2 |
| Lean IR/ Hyperglycemia | Endocrine–inflammatory metabolic stress signaling | chemotaxis | GO | IL6/SERPINE1 |
| Lean IR/ Hyperglycemia | Endocrine–inflammatory metabolic stress signaling | lipid localization | GO | LEP/SLC4A1/ESYT2/IL6 |
| Lean IR/ Hyperglycemia | Endocrine–inflammatory metabolic stress signaling | positive regulation of MAPK cascade | GO | LEP/FGF21/IL6 |
| Lean IR/ Hyperglycemia | Endocrine–inflammatory metabolic stress signaling | cellular catabolic process | GO | LEP/MPST/HAO1 |
| Lean IR/ Hyperglycemia | Endocrine–inflammatory metabolic stress signaling | leukocyte migration | GO | LEP/IL6/SERPINE1 |
| Obese IS/Normoglycemia | Hepatic oxidative and small-molecule catabolic profile | Tyrosine metabolism | KEGG | ADH4/HPD/DBH/ADH1B |
| Obese IS/Normoglycemia | Hepatic oxidative and small-molecule catabolic profile | Pyruvate metabolism | KEGG | ADH4/ADH1B |
| Obese IS/Normoglycemia | Hepatic oxidative and small-molecule catabolic profile | response to oxidative stress | GO | HAO1/IL1A/C19orf12/PARP1/ AIFM1 |
| Obese IS/Normoglycemia | Hepatic oxidative and small-molecule catabolic profile | response to peptide hormone | GO | IGFBP1/OXT/ECHDC3/PARP1/LEP |
| Obese IS/Normoglycemia | Hepatic oxidative and small-molecule catabolic profile | cellular catabolic process | GO | HAO1/HPD/FTCD/ECHS1/LEP/DCXR |
| Obese IS/Normoglycemia | Hepatic oxidative and small-molecule catabolic profile | small molecule catabolic process | GO | HAO1/ADH4/HPD/FTCD/ ECHS1/LEP/DCXR |
| Obese IR/Hyperglycemia | Immuno-metabolic inflammation with adhesion/migration remodeling | MAPK signaling pathway | KEGG | IL1A/FGF21/ARTN |
| Obese IR/Hyperglycemia | Immuno-metabolic inflammation with adhesion/migration remodeling | Cytokine-cytokine receptor interaction | KEGG | IL31/GH1/IL1A/IL6/LEP |
| Obese IR/Hyperglycemia | Immuno-metabolic inflammation with adhesion/migration remodeling | PI3K-Akt signaling pathway | KEGG | GH1/IL6/FGF21/TCL1A/ARTN |
| Obese IR/Hyperglycemia | Immuno-metabolic inflammation with adhesion/migration remodeling | IgSF CAM signaling | KEGG | SPTBN5/WASL/MTSS2 |
| Obese IR/Hyperglycemia | Immuno-metabolic inflammation with adhesion/migration remodeling | JAK-STAT signaling pathway | KEGG | IL31/GH1/IL6/LEP/STAT5B |
| Obese IR/Hyperglycemia | Immuno-metabolic inflammation with adhesion/migration remodeling | Alcoholic liver disease | KEGG | ADH4/IL6/ADH1B |

| | | | | |
|---|---|---|---|---|
| | | Lipid and atherosclerosis | KEGG | IL6/PLCB2 |
| | | leukocyte cell-cell adhesion | GO | RIPOR2/IL1A/IGFBP2/IL6/LEP/STAT5B |
| | | axonogenesis | GO | LHX2/IGSF9/ARTN |
| | | positive regulation of cell development | GO | IL6/HIF1A/STAT5B |
| | | response to peptide hormone | GO | IGFBP1/GH1/PNPT1/OXT/LEP/GHRHR/FGF21/STAT5B/KHK |
| | | regulation of inflammatory response | GO | ESR1/IL6/SERPINE1/EXTL3/STAT5B |
| | | leukocyte migration | GO | RIPOR2/WASL/IL1A/IL6/SERPINE1/LEP/STAT5B/ARTN |

**Table S16 Cardiometabolic traits, phenotypes and events according to 5 Latent classes in validation cohort–CHARLS**

| Variable | **Overall** | **Class 1** | **Class 2** | **Class 3** | **Class 4** | **Class 5** | **p** |
|---|---|---|---|---|---|---|---|
| n | 12145 | 3036 | 1267 | 1523 | 4834 | 1485 | |
| Age , mean (SD), y | 55.4 (7.6) | 55.1 (7.5) | 58.4 (7.3) | 54.8 (7.4) | 54.9 (7.6) | 55.5 (7.5) | <0.001 |
| Male (%) | 5663 (46.6) | 2094 (69.0) | 1004 ( 79.2) | 725 ( 47.6) | 1466 ( 30.3) | 374 ( 25.2) | <0.001 |
| Waist circumference, mean (SD) | 84.5 (12.4) | 76.1 (10.1) | 77.6 (10.8) | 75.3 (13.4) | 91.6 (7.3) | 93.9 (8.0) | <0.001 |
| Body mass index , mean (SD) | 24.3 (25.0) | 21.2 (4.0) | 22.3 (14.6) | 23.0 (21.0) | 26.4 (36.6) | 26.9 (7.0) | <0.001 |
| Body mass index , mean (SD), kg/$m^2$ | 125.7 (19.6) | 112.9 (10.1) | 145.4 (14.7) | 123.6 (18.7) | 128.4 (20.1) | 128.4 (19.3) | <0.001 |
| Waist circumference, mean (SD), cm | 75.3 (12.0) | 68.5 (8.9) | 84.0 (10.3) | 74.1 (11.5) | 77.0 (11.9) | 77.4 (12.1) | <0.001 |
| TG, mean (SD), mmol/L | 1.6 (1.2) | 0.9 (0.3) | 1.0 (0.4) | 2.3 (1.4) | 1.6 (1.2) | 2.5 (1.5) | <0.001 |
| HDL-C , mean (SD), mmol/L | 1.3 (0.4) | 1.5 (0.4) | 1.5 (0.4) | 1.1 (0.3) | 1.3 (0.3) | 1.2 (0.2) | <0.001 |
| LDL-C, mean (SD), mmol/L | 2.8 (0.9) | 2.7 (0.8) | 2.8 (0.8) | 2.7 (0.9) | 2.9 (0.9) | 2.9 (0.9) | <0.001 |
| FPG, mean (SD), mmol/L | 5.9 (2.0) | 5.5 (1.3) | 5.7 (1.6) | 6.1 (2.2) | 5.8 (1.5) | 7.2 (3.5) | <0.001 |
| HbA1c, mean (SD), % | 5.5 (0.9) | 5.4 (0.7) | 5.4 (0.8) | 5.4 (0.9) | 5.3 (0.7) | 6.6 (1.3) | <0.001 |
| TG_HDL , mean (SD) | 1.4 (2.2) | 0.7 (0.3) | 0.7 (0.3) | 2.4 (3.5) | 1.5 (2.1) | 2.4 (2.8) | <0.001 |
| Uric acid, mean (SD), mmol/L | 0.27 (0.08) | 0.27 (0.07) | 0.28 (0.08) | 0.28 (0.09) | 0.27 (0.08) | 0.29 (0.08) | <0.001 |
| SBP, mean (SD), mmHg | 125.67 (19.61) | 112.88 (10.09) | 145.45 (14.66) | 123.57 (18.69) | 128.36 (20.13) | 128.36 (19.30) | <0.001 |
| DBP, mean (SD), mmHg | 75.27 (11.96) | 68.51 (8.88) | 83.99 (10.33) | 74.07 (11.48) | 76.95 (11.93) | 77.38 (12.07) | <0.001 |
| Obesity (%) | 1780 (14.7) | 14 ( 0.5) | 19 ( 1.5) | 48 ( 3.2) | 1161 ( 24.1) | 538 ( 36.4) | <0.001 |
| Central Obesity by WC (%) | 6319 (52.0) | 0 ( 0.0) | 0 ( 0.0) | 0 ( 0.0) | 4834 (100.0) | 1485 (100.0) | <0.001 |
| low HDL (%) | 4336 (35.7) | 301 ( 9.9) | 107 ( 8.4) | 873 ( 57.3) | 2118 ( 43.8) | 937 ( 63.1) | <0.001 |
| high TG (%) | 3446 (28.4) | 90 ( 3.0) | 64 ( 5.1) | 930 ( 61.1) | 1371 ( 28.4) | 991 ( 66.7) | <0.001 |
| high LDL (%) | 2758 (22.7) | 540 (17.8) | 269 ( 21.2) | 316 ( 20.8) | 1286 ( 26.6) | 347 ( 23.4) | <0.001 |
| high FPG (%) | 5519 (45.5) | 990 (32.6) | 560 ( 44.2) | 746 ( 49.1) | 2273 ( 47.1) | 950 ( 64.0) | <0.001 |
| high HbA1c (%) | 3658 (30.1) | 671 (22.1) | 302 ( 23.8) | 406 ( 26.7) | 794 ( 16.4) | 1485 (100.0) | <0.001 |
| IR by TG:HDL (%) | 5297 (43.6) | 0 ( 0.0) | 0 ( 0.0) | 1523 (100.0) | 2289 ( 47.4) | 1485 (100.0) | <0.001 |
| high SBP (%) | 4397 (36.2) | 0 ( 0.0) | 1267 (100.0) | 489 ( 32.1) | 2004 ( 41.5) | 637 ( 42.9) | <0.001 |
| high DBP (%) | 2422 (19.9) | 82 ( 2.7) | 557 ( 44.0) | 247 ( 16.2) | 1174 ( 24.3) | 362 ( 24.4) | <0.001 |
| Hyperuricemia (%) | 836 ( 6.9) | 105 ( 3.5) | 67 ( 5.3) | 129 ( 8.5) | 334 ( 6.9) | 201 ( 13.5) | <0.001 |
| Metabolic syndrome–IDF (%) | 3831 (31.6) | 0 ( 0.0) | 0 ( 0.0) | 0 ( 0.0) | 2627 ( 54.4) | 1204 ( 81.1) | <0.001 |
| event_CVD (%) | 1743 (14.4) | 273 (9.0) | 178 (14.0) | 210 (13.8) | 813 (16.8) | 269 (18.1) | <0.001 |
| event_STROKE (%) | 706 (5.8) | 104 (3.4) | 94 (7.4) | 90 (5.9) | 299 (6.2) | 119 (8.0) | <0.001 |

The five latent classes were identified by decision tree model in The China Health and Retirement Longitudinal Study (CHARLS, N=12145). In CHARLS, the insulin resistance was replaced by the surrogate indicator high TG:HDL ratio (≥1.51 mmol/L in men and 0.84 mmol/L in women), and the visceral adiposity was replaced by the surrogate indicator central obesity (waist circumference≥80cm for female and ≥90cm for male).

**Table S17 Cardiometabolic traits, phenotypes and events according to 5 Latent classes in validation cohort–UK Biobank**

| Variable | Overall | Class 1 | Class 2 | Class 3 | Class 4 | Class 5 | p |
|---|---|---|---|---|---|---|---|
| n | 344817 | 70179 | 93995 | 67811 | 91198 | 21634 | |
| Age , mean (SD), y | 56.4 (8.0) | 52.7 (7.9) | 57.8 (7.7) | 56.5 (8.0) | 56.9 (7.8) | 59.4 (6.9) | <0.001 |
| Male (%) | 157191 (45.6) | 22238 (31.7) | 48311 ( 51.4) | 39819 ( 58.7) | 37363 ( 41.0) | 9460 ( 43.7) | <0.001 |
| Body mass index , mean (SD), kg/m$^2$ | 27.3 (4.7) | 24.0 (2.8) | 25.1 (2.7) | 26.2 (2.6) | 31.6 (4.2) | 33.4 (5.0) | <0.001 |
| Waist circumference, mean (SD), cm | 90.0 (13.4) | 79.4 (9.0) | 83.9 (9.4) | 87.9 (8.6) | 102.0 (10.0) | 106.7 (11.6) | <0.001 |
| TG, mean (SD), mmol/L | 1.7 (1.0) | 1.1 (0.4) | 1.2 (0.4) | 2.6 (1.0) | 1.9 (1.0) | 2.8 (1.2) | <0.001 |
| HDL-C , mean (SD), mmol/L | 1.5 (0.4) | 1.6 (0.4) | 1.6 (0.4) | 1.2 (0.2) | 1.4 (0.3) | 1.2 (0.2) | <0.001 |
| LDL-C, mean (SD), mmol/L | 3.6 (0.9) | 3.4 (0.8) | 3.6 (0.8) | 3.8 (0.9) | 3.7 (0.9) | 3.5 (1.0) | <0.001 |
| FPG, mean (SD), mmol/L | 5.1 (1.2) | 4.8 (0.8) | 5.0 (0.9) | 5.1 (1.1) | 5.1 (0.9) | 6.3 (2.6) | <0.001 |
| HbA1c, mean (SD), % | 5.4 (0.6) | 5.3 (0.4) | 5.3 (0.4) | 5.4 (0.5) | 5.4 (0.5) | 6.4 (1.0) | <0.001 |
| TG/HDL,mean (SD) | 3.1 (2.5) | 1.6 (0.7) | 1.8 (0.7) | 5.1 (2.6) | 3.6 (2.6) | 5.8 (3.2) | <0.001 |
| SBP, mean (SD), mmHg | 138.0 (18.6) | 118.4 (8.0) | 147.6 (14.3) | 139.0 (18.0) | 141.1 (17.9) | 143.4 (17.5) | <0.001 |
| DBP, mean (SD), mmHg | 82.4 (10.1) | 73.5 (6.8) | 85.4 (8.8) | 82.7 (9.9) | 85.3 (9.8) | 84.8 (9.7) | <0.001 |
| hsCRP, mean (SD), mg/L | 2.5 (4.2) | 1.6 (3.6) | 1.9 (3.9) | 2.3 (3.7) | 3.5 (4.6) | 4.7 (5.6) | <0.001 |
| Uric acid, mean (SD), mmol/L | 307.5 (79.5) | 267.4 (68.2) | 296.1 (73.6) | 321.4 (78.3) | 329.6 (78.8) | 350.2 (80.0) | <0.001 |
| 10y ASCVD risk score,(mean (SD) | 4.4 (3.2) | 2.0 (1.6) | 4.6 (2.8) | 5.2 (3.3) | 4.8 (3.0) | 7.2 (3.9) | <0.001 |
| Metabolic syndrome–IDF (%) | 49159 (14.3) | 0 ( 0.0) | 0 ( 0.0) | 0 ( 0.0) | 35298 ( 38.7) | 13861 ( 64.1) | <0.001 |
| Obesity classification by BMI (%) | | | | | | | <0.001 |
| Underweight | 1766 ( 0.5) | 1028 ( 1.5) | 606 ( 0.6) | 128 ( 0.2) | 4 ( 0.0) | 0 ( 0.0) | |
| Normal weight | 114297 (33.1) | 44694 (63.7) | 45943 ( 48.9) | 21450 ( 31.6) | 2038 ( 2.2) | 172 ( 0.8) | |
| Overweight | 147178 (42.7) | 23017 (32.8) | 44048 ( 46.9) | 41097 ( 60.6) | 33668 ( 36.9) | 5348 ( 24.7) | |
| Obesity | 81576 (23.7) | 1440 ( 2.1) | 3398 ( 3.6) | 5136 ( 7.6) | 55488 ( 60.8) | 16114 ( 74.5) | |
| Central Obesity by WC (%) | 112832 (32.7) | 0 ( 0.0) | 0 ( 0.0) | 0 ( 0.0) | 91198 (100.0) | 21634 (100.0) | <0.001 |
| IR by TG/HDL-C (%) | 134389 (39.0) | 0 ( 0.0) | 0 ( 0.0) | 67811 (100.0) | 44944 ( 49.3) | 21634 (100.0) | <0.001 |
| Diabetes status (%) | | | | | | | <0.001 |
| Nomorglycemia | 285677 (82.8) | 64603 (92.1) | 82524 ( 87.8) | 56679 ( 83.6) | 80524 ( 88.3) | 1347 ( 6.2) | |
| Pre DM | 43327 (12.6) | 4515 ( 6.4) | 9160 ( 9.7) | 8941 ( 13.2) | 6843 ( 7.5) | 13868 ( 64.1) | |
| DM | 15813 ( 4.6) | 1061 ( 1.5) | 2311 ( 2.5) | 2191 ( 3.2) | 3831 ( 4.2) | 6419 ( 29.7) | |
| Hypertension (%) | 182788 (53.0) | 5133 ( 7.3) | 65168 ( 69.3) | 36360 ( 53.6) | 59230 ( 64.9) | 16897 ( 78.1) | <0.001 |
| Dyslipidemia (%) | 165137 (47.9) | 21137 (30.1) | 42938 ( 45.7) | 37354 ( 55.1) | 48455 ( 53.1) | 15253 ( 70.5) | <0.001 |
| Anti-diabetic drug (%) | 10341 ( 3.0) | 642 ( 0.9) | 1464 ( 1.6) | 1294 ( 1.9) | 2302 ( 2.5) | 4639 ( 21.4) | <0.001 |
| Anti-hypertension drug (%) | 63599 (18.5) | 3094 ( 4.4) | 15895 ( 16.9) | 11391 ( 16.8) | 23598 ( 25.9) | 9621 ( 44.5) | <0.001 |
| Lipid-lowering drug (%) | 50298 (14.6) | 3882 ( 5.5) | 11600 ( 12.3) | 9719 ( 14.3) | 16108 ( 17.7) | 8989 ( 41.6) | <0.001 |
| event Myocardial infarction (%) | 8631 ( 2.5) | 666 ( 0.9) | 2180 ( 2.3) | 2316 ( 3.4) | 2449 ( 2.7) | 1020 ( 4.7) | <0.001 |
| event Stroke (%) | 8031 ( 2.3) | 806 ( 1.1) | 2417 ( 2.6) | 1579 ( 2.3) | 2333 ( 2.6) | 896 ( 4.1) | <0.001 |
| low_HDL (%) | 69267 (20.1) | 4633 ( 6.6) | 3480 ( 3.7) | 23648 ( 34.9) | 25410 ( 27.9) | 12096 ( 55.9) | <0.001 |
| high_TG (%) | 137834 (40.0) | 3802 ( 5.4) | 10108 ( 10.8) | 60050 ( 88.6) | 44550 ( 48.8) | 19324 ( 89.3) | <0.001 |
| high_LDL (%) | 198656 (57.6) | 32274 (46.0) | 52583 ( 55.9) | 46409 ( 68.4) | 56145 ( 61.6) | 11245 ( 52.0) | <0.001 |

| | | | | | | | |
|---|---|---|---|---|---|---|---|
| high_FPG (%) | 49234 (14.3) | 4985 ( 7.1) | 11757 ( 12.5) | 9600 ( 14.2) | 13431 ( 14.7) | 9461 ( 43.7) | <0.001 |
| high_HbA1c (%) | 58961 (17.1) | 5291 ( 7.5) | 11144 ( 11.9) | 11165 ( 16.5) | 9727 ( 10.7) | 21634 (100.0) | <0.001 |
| high_hsCRP (%) | 119906 (34.8) | 12322 (17.6) | 22122 ( 23.5) | 22376 ( 33.0) | 48685 ( 53.4) | 14401 ( 66.6) | <0.001 |
| high_SBP (%) | 222686 (64.6) | 0 ( 0.0) | 93995 (100.0) | 45751 ( 67.5) | 66090 ( 72.5) | 16850 ( 77.9) | <0.001 |
| high_DBP (%) | 135527 (39.3) | 3142 ( 4.5) | 48017 ( 51.1) | 27230 ( 40.2) | 46506 ( 51.0) | 10632 ( 49.1) | <0.001 |
| Hyperuricemia (%) | 42346 (12.3) | 2465 ( 3.5) | 6481 ( 6.9) | 8912 ( 13.1) | 18106 ( 19.9) | 6382 ( 29.5) | <0.001 |

The five latent classes were identified by decision tree model in UK Biobank (N= 344817). In UK Biobank, the IR was replaced by the surrogate indicator high TG:HDL ratio (≥3.5 mmol/L in male and 2.5 mmol/L in female), and the visceral adiposity was replaced by the surrogate indicator central obesity (WC≥88cm for female and ≥102cm for male)

**Table S18 Cox proportional models for evaluating the association between latent classes and incident outcomes**

| Cohort | Outcome | Model | Class 2 vs 1 | Class 3 vs 1 | Class 4 vs 1 | Class 5 vs 1 |
|---|---|---|---|---|---|---|
| CHARLS | CVD | Model 1 | 1.52 (1.26–1.84) | 1.53 (1.27–1.83) | 1.72 (1.49–1.99) | 2.48 (2.08–2.95) |
| | | Model 2 | 1.32 (1.07–1.62) | 1.51 (1.24–1.85) | 1.44 (1.20–1.73) | 1.62 (1.27–2.05) |
| | Stroke | Model 1 | 1.89 (1.43–2.50) | 2.00 (1.50–2.65) | 2.18 (1.72–2.75) | 4.63 (3.51–6.10) |
| | | Model 2 | 1.14 (0.84–1.55) | 1.49 (1.09–2.04) | 1.24 (0.92–1.68) | 1.82 (1.24–2.67) |
| | CVD/Stroke | Model 1 | 1.56 (1.33–1.84) | 1.61 (1.37–1.88) | 1.79 (1.58–2.04) | 2.78 (2.38–3.25) |
| | | Model 2 | 1.24 (1.04–1.48) | 1.49 (1.25–1.78) | 1.35 (1.14–1.59) | 1.57 (1.27–1.94) |
| UK Biobank | MI | Model 1 | 1.63 (1.49–1.78) | 2.43 (2.22–2.65) | 2.18 (2.00–2.37) | 3.49 (3.16–3.85) |
| | | Model 2 | 1.23 (1.12–1.36) | 1.42 (1.28–1.57) | 1.29 (1.15–1.44) | 1.31 (1.14–1.50) |
| | Stroke | Model 1 | 1.42 (1.31–1.54) | 1.37 (1.25–1.49) | 1.58 (1.46–1.71) | 2.24 (2.03–2.46) |
| | | Model 2 | 1.03 (0.94–1.13) | 0.98 (0.89–1.09) | 1.00 (0.90–1.12) | 1.08 (0.94–1.24) |
| | MI/Stroke | Model 1 | 1.52 (1.43–1.61) | 1.86 (1.75–1.98) | 1.85 (1.75–1.97) | 2.75 (2.56–2.95) |
| | | Model 2 | 1.13 (1.06–1.21) | 1.19 (1.11–1.28) | 1.14 (1.05–1.23) | 1.16 (1.06–1.28) |

Cox proportional models for incident cardiovascular disease (CVD), stroke and CVD/stroke outcomes in 5 classes identified by the decision tree model in The China Health and Retirement Longitudinal Study (CHARLS, n=12,145); and for incident myocardial infarction (MI), stroke and MI/stroke outcomes in 5 classes identified by the decision tree model in the UK Biobank (n= 34,4817).

Model 1: age and sex; Model 2: age, sex, smoking status, WC, TG, HDL cholesterol, fasting glucose, HbA1c, anti-hypertensive, diabetic and dyslipidemic drugs

**Table S19 Events numbers and events rates per 1000 person-years for CVD, stroke and CVD/stroke in CHARLS and UK Biobank**

### CHARLS

| Class | CVD events | CVD person years | CVD rates per 1000 person years (95%CI) | Stroke events | Stroke person years | Stroke rates per 1000 person years (95%CI) | CVD/stroke events | CVD/stroke person years | CVD/stroke rates per 1000 person years (95%CI) |
|---|---|---|---|---|---|---|---|---|---|
| 1 | 273 | 20251 | 13.5 (11.9-15.2) | 104 | 20695 | 5 (4.1-6.1) | 358 | 20085 | 17.8 (16-19.8) |
| 2 | 178 | 8228 | 21.6 (18.6-25.1) | 94 | 8430 | 11.2 (9-13.6) | 243 | 8067 | 30.1 (26.5-34.2) |
| 3 | 210 | 9910 | 21.2 (18.4-24.3) | 90 | 10264 | 8.8 (7.1-10.8) | 274 | 9775 | 28 (24.8-31.6) |
| 4 | 813 | 32096 | 25.3 (23.6-27.1) | 299 | 33712 | 8.9 (7.9-9.9) | 1015 | 31674 | 32 (30.1-34.1) |
| 5 | 269 | 7813 | 34.4 (30.4-38.8) | 119 | 8065 | 14.8 (12.2-17.7) | 344 | 7675 | 44.8 (40.2-49.8) |

### UK Biobank

| Class | MI events | MI person years | MI rates per 1000 person years (95%CI) | Stroke events | Stroke person years | Stroke rates per 1000 person years (95%CI) | MI/stroke events | MI/stroke person years | MI/stroke rates per 1000 person years (95%CI) |
|---|---|---|---|---|---|---|---|---|---|
| 1 | 666 | 976864 | 0.7 (0.6-0.7) | 806 | 976837 | 0.8 (0.8-0.9) | 1445 | 973708 | 1.5 (1.4-1.6) |
| 2 | 2180 | 1285342 | 1.7 (1.6-1.8) | 2417 | 1286674 | 1.9 (1.8-2) | 4469 | 1275415 | 3.5 (3.4-3.6) |
| 3 | 2316 | 925848 | 2.5 (2.4-2.6) | 1579 | 932201 | 1.7 (1.6-1.8) | 3795 | 919133 | 4.1 (4-4.3) |
| 4 | 2449 | 1239598 | 2 (1.9-2.1) | 2333 | 1242919 | 1.9 (1.8-2) | 4645 | 1229914 | 3.8 (3.7-3.9) |
| 5 | 1020 | 286714 | 3.6 (3.3-3.8) | 896 | 288330 | 3.1 (2.9-3.3) | 1813 | 283356 | 6.4 (6.1-6.7) |

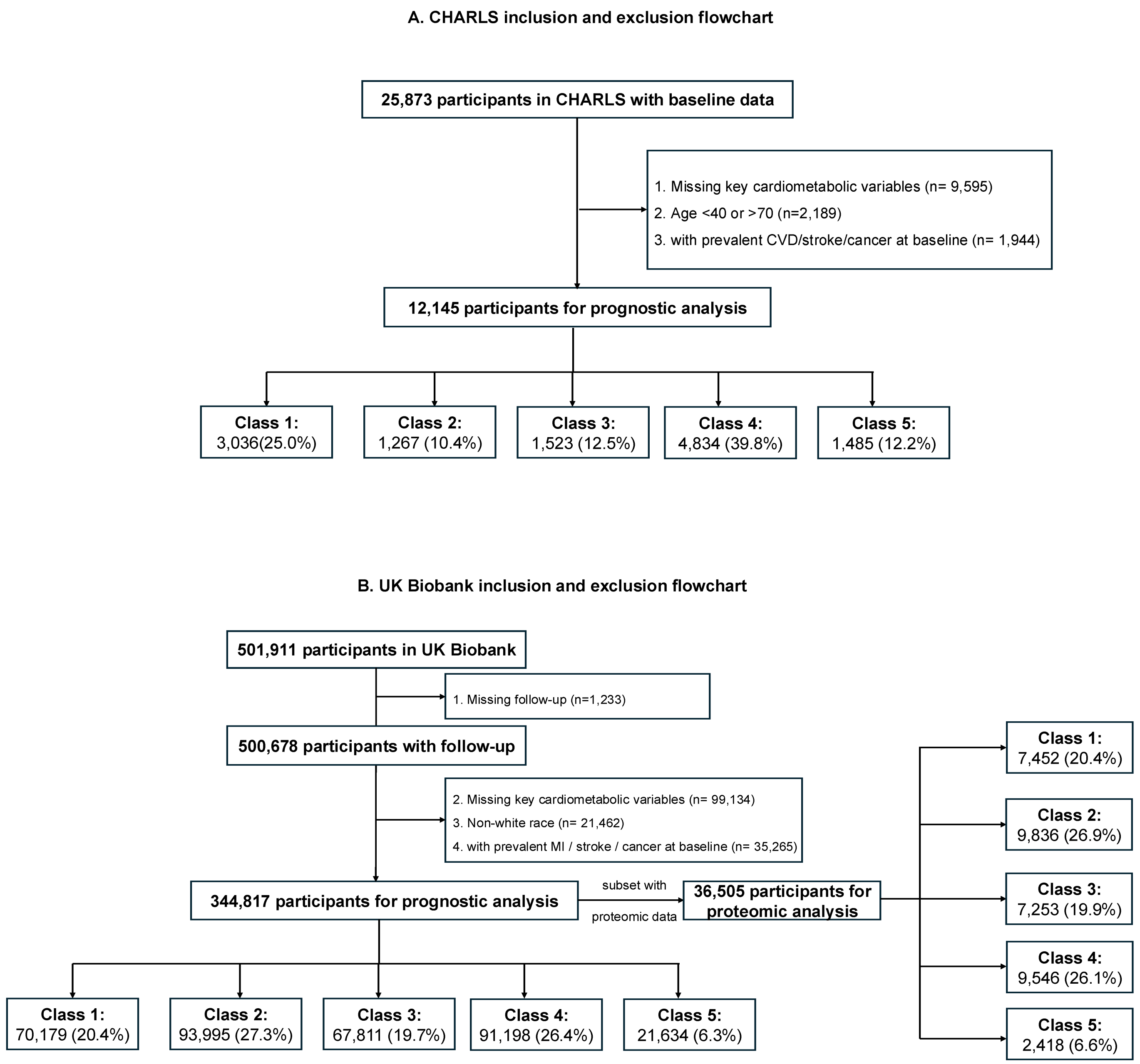


**Figure S1 Inclusion and exclusion flowcharts in CHARLS and UK Biobank**

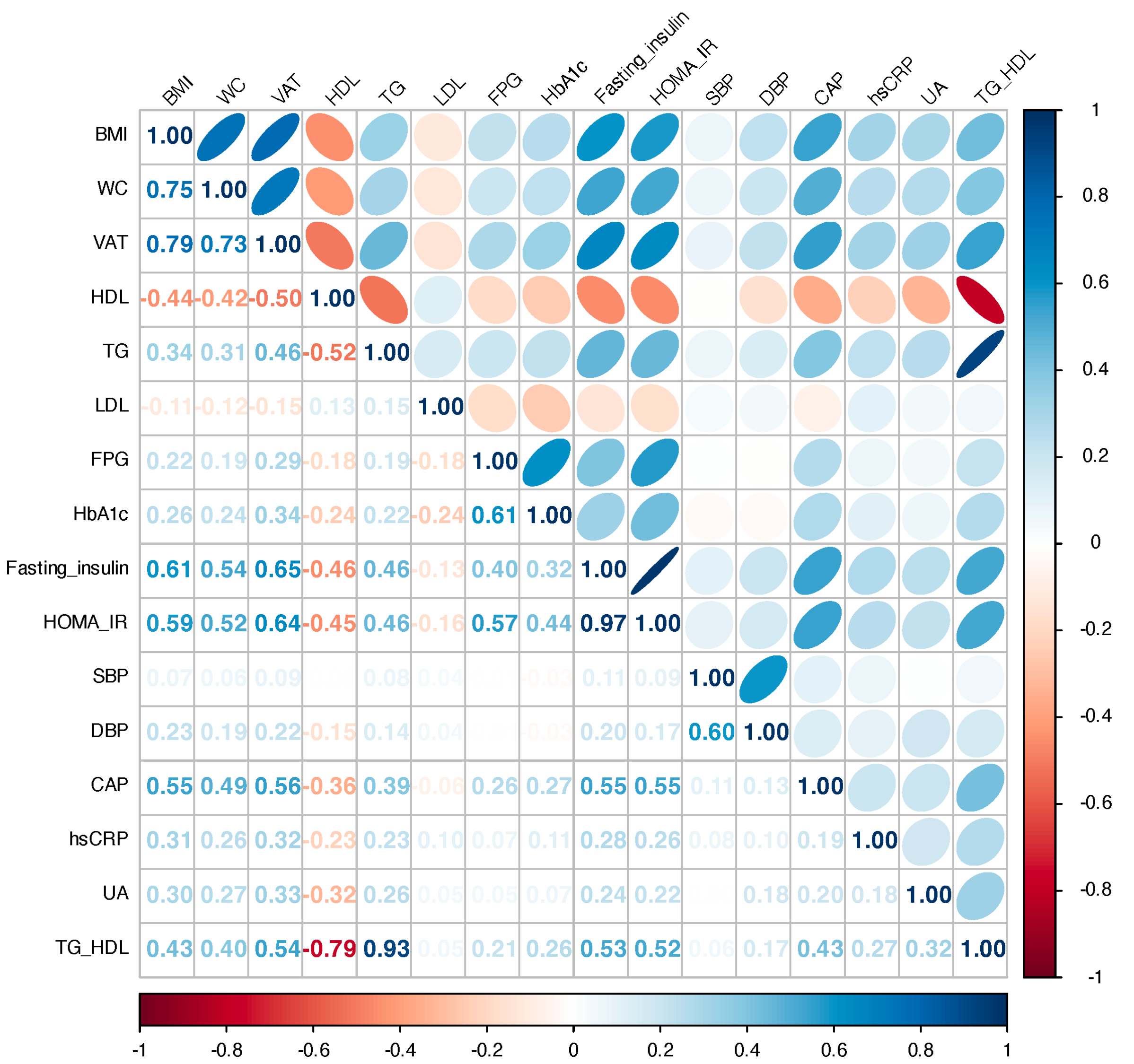


**Figure S2 The heatmap of correlation matrix for all related continuous cardiometabolic traits**

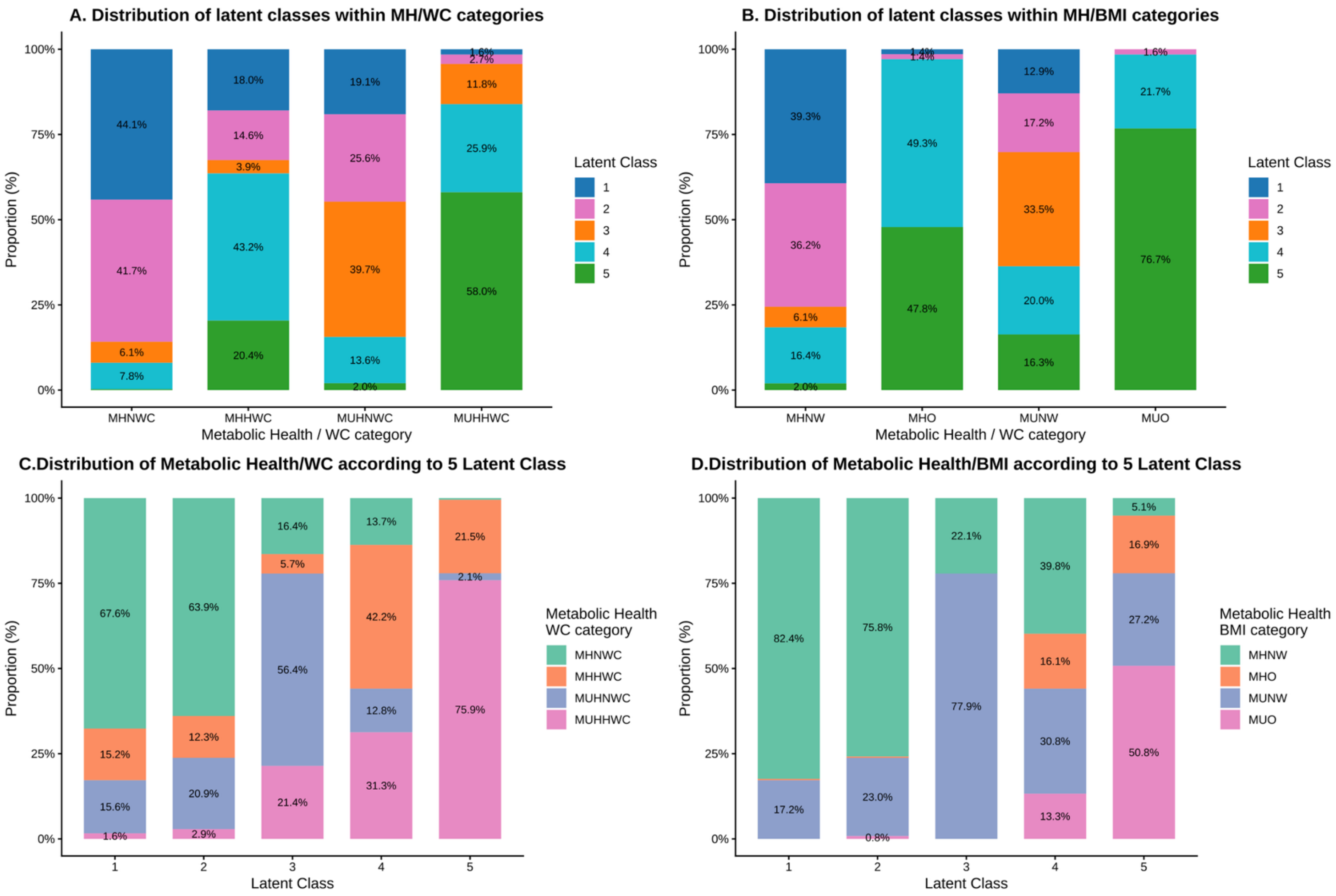


**Figure S3 Distribution of the five latent classes across categories of metabolic health and WC/BMI**

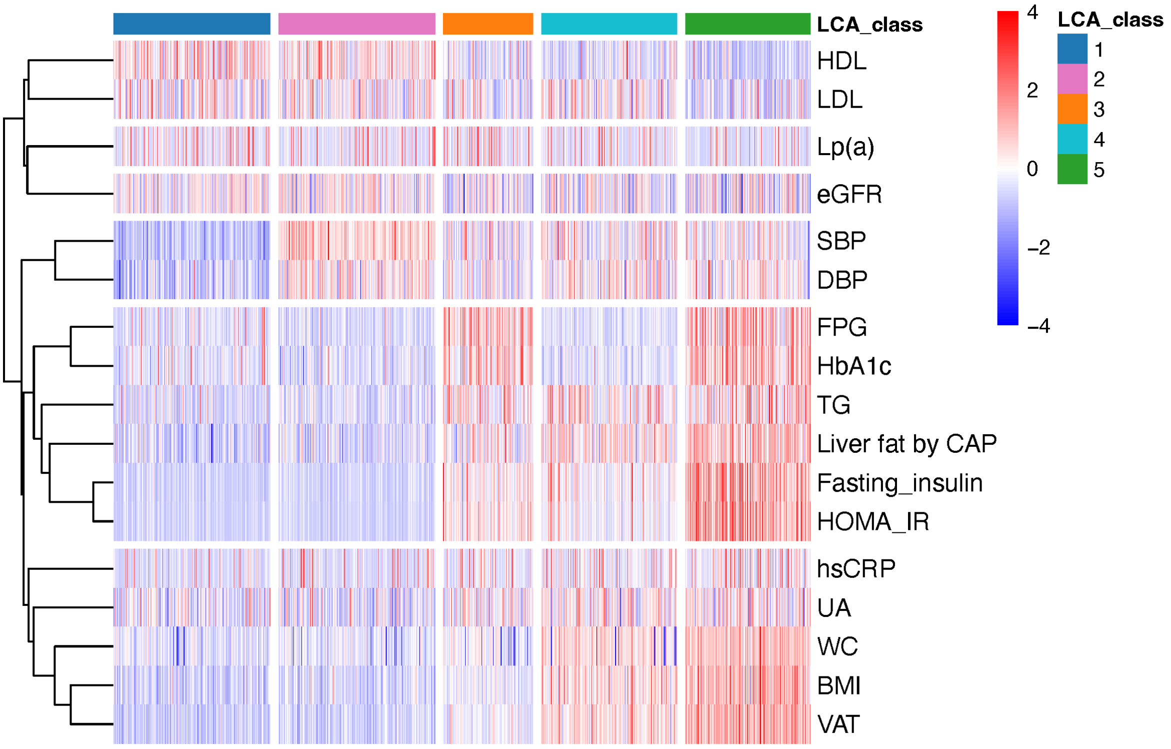


**Figure S4 Comparison of the metabolic traits as continuous variables among five latent classes from LCA in RESET**

All continuous variables were z-score scaled. Latent Class 1 : Metabolic preserved, Class 2 : Isolated Hypertension, Class 3 : Lean IR/Hyperglycemia, Class 4 : Obese IS/Normoglycemia, Class 5 : Obese IR/Hyperglycemia.

LDL, Low density lipoprotein cholesterol; HDL, High density lipoprotein cholesterol; Lp(a), Lipoprotein (a); SBP, systolic blood pressure; DBP, diastolic blood pressure; FPG, Fasting plasma glucose; HbA1c, Glycated Hemoglobin A1c; TG, triglyceride; HOMA-IR, Homeostatic Model Assessment of Insulin Resistance; CAP, Controlled attenuation parameter (Db/m); hsCRP, High Sensitivity C-reactive Protein; UA, Uric acid; WC, Waist circumference; BMI, Body mass index; VAT, Visceral adiposity tissue area by DEXA.

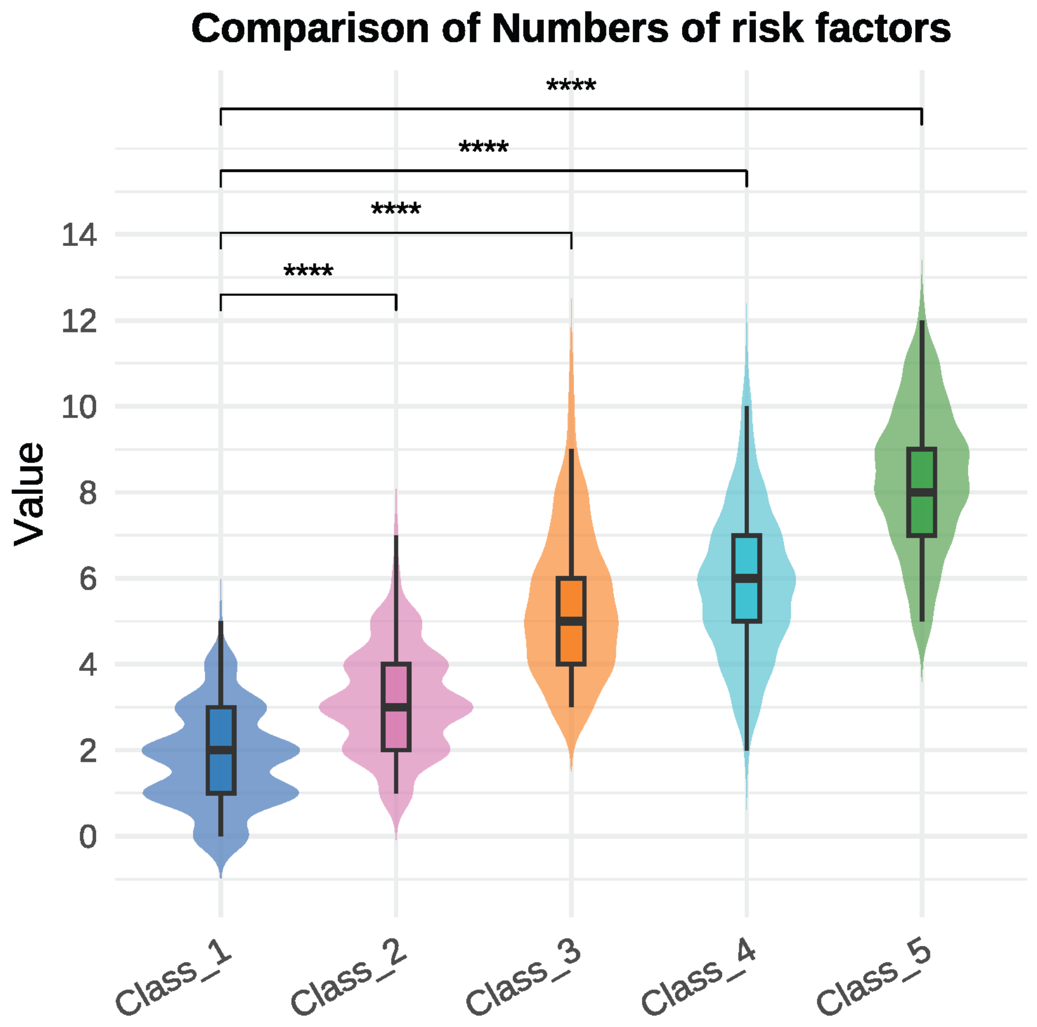


**Figure S5 Violin plot of the numbers of abnormal binary metabolic phenotypes among five latent classes**

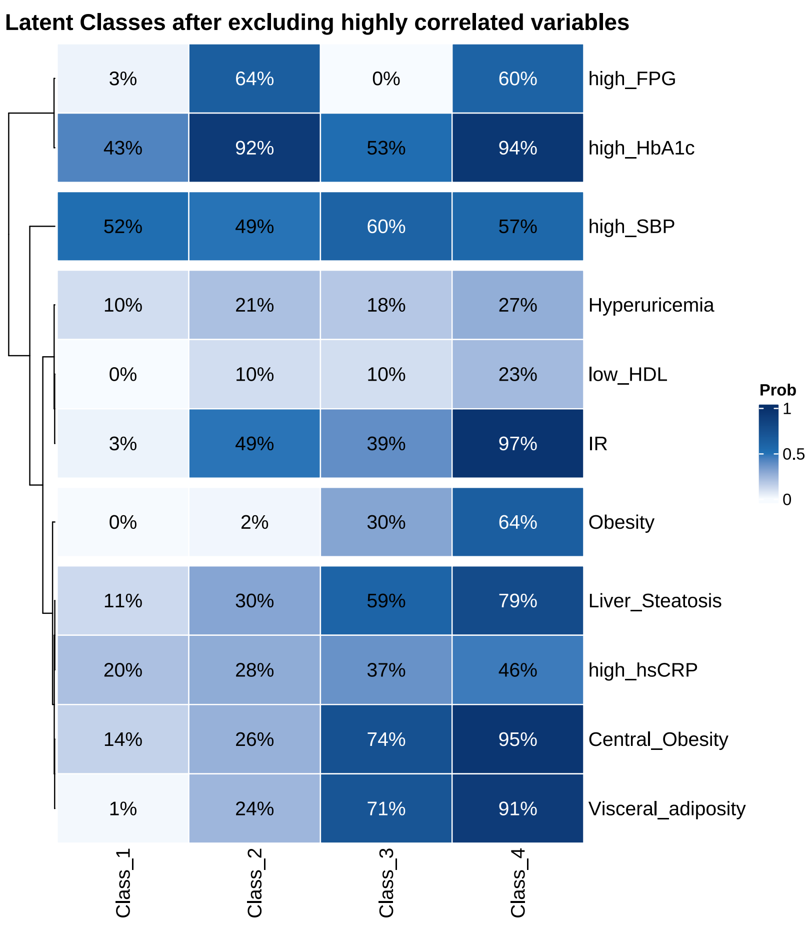


| | Class_1 | Class_2 | Class_3 | Class_4 |
|---|---|---|---|---|
| high_FPG | 3% | 64% | 0% | 60% |
| high_HbA1c | 43% | 92% | 53% | 94% |
| high_SBP | 52% | 49% | 60% | 57% |
| Hyperuricemia | 10% | 21% | 18% | 27% |
| low_HDL | 0% | 10% | 10% | 23% |
| IR | 3% | 49% | 39% | 97% |
| Obesity | 0% | 2% | 30% | 64% |
| Liver_Steatosis | 11% | 30% | 59% | 79% |
| high_hsCRP | 20% | 28% | 37% | 46% |
| Central_Obesity | 14% | 26% | 74% | 95% |
| Visceral_adiposity | 1% | 24% | 71% | 91% |

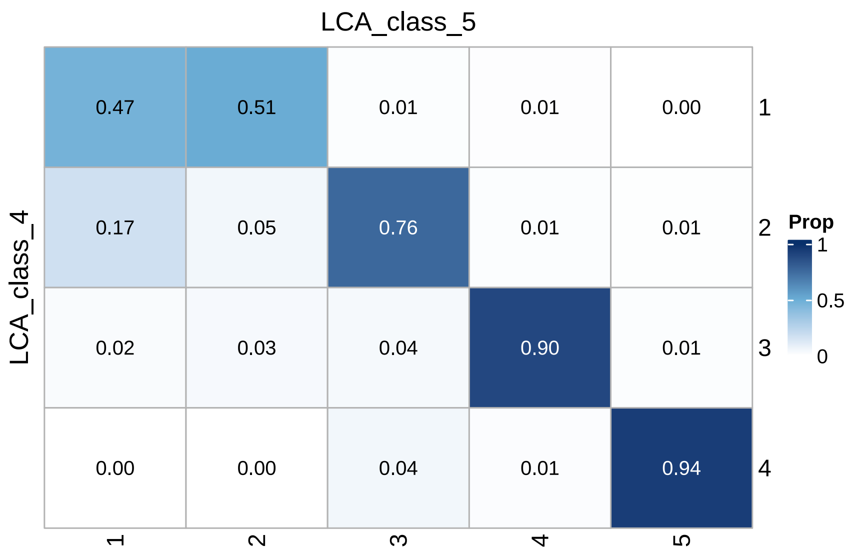


| LCA_class_4 \ LCA_class_5 | 1 | 2 | 3 | 4 | 5 |
|---|---|---|---|---|---|
| 1 | 0.47 | 0.51 | 0.01 | 0.01 | 0.00 |
| 2 | 0.17 | 0.05 | 0.76 | 0.01 | 0.01 |
| 3 | 0.02 | 0.03 | 0.04 | 0.90 | 0.01 |
| 4 | 0.00 | 0.00 | 0.04 | 0.01 | 0.94 |

**Figure S6 Sensitivity analysis of LCA model after excluding highly correlated variables.**
The sensitivity analysis of excluding highly correlated variables (exclude high TG, high LDL and high DBP and retain low HDL and high SBP) for LCA showed that subtypes similar with the original latent classes still existed. The optimal latent classes numbers for the model after excluding highly correlated variables are four according to the lowest BIC.

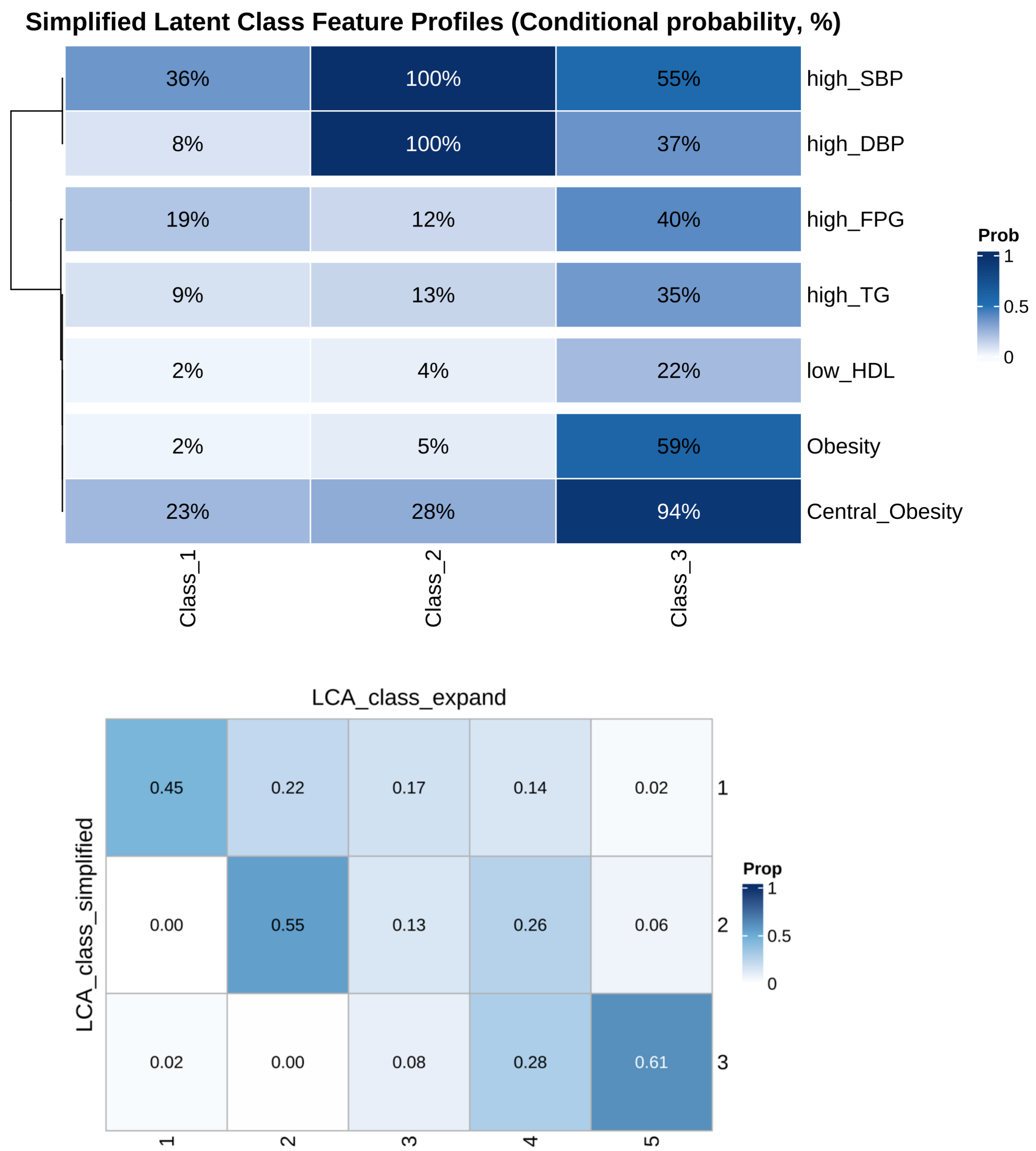


**Figure S7 Sensitivity analysis of using simplified metabolic risk factors for clustering.**
The sensitivity analysis of using simplified metabolic risk factors (included in the IDF criteria of metabolic syndrome and obesity defined by BMI) for LCA showed that reduced number of risk factors are unable to classify sub-patterns such as Lean-IR/Hyperglycemia or Obese-IS/Normoglycemia. The optimal latent classes numbers for the model using 7 variables are three according to the lowest BIC.

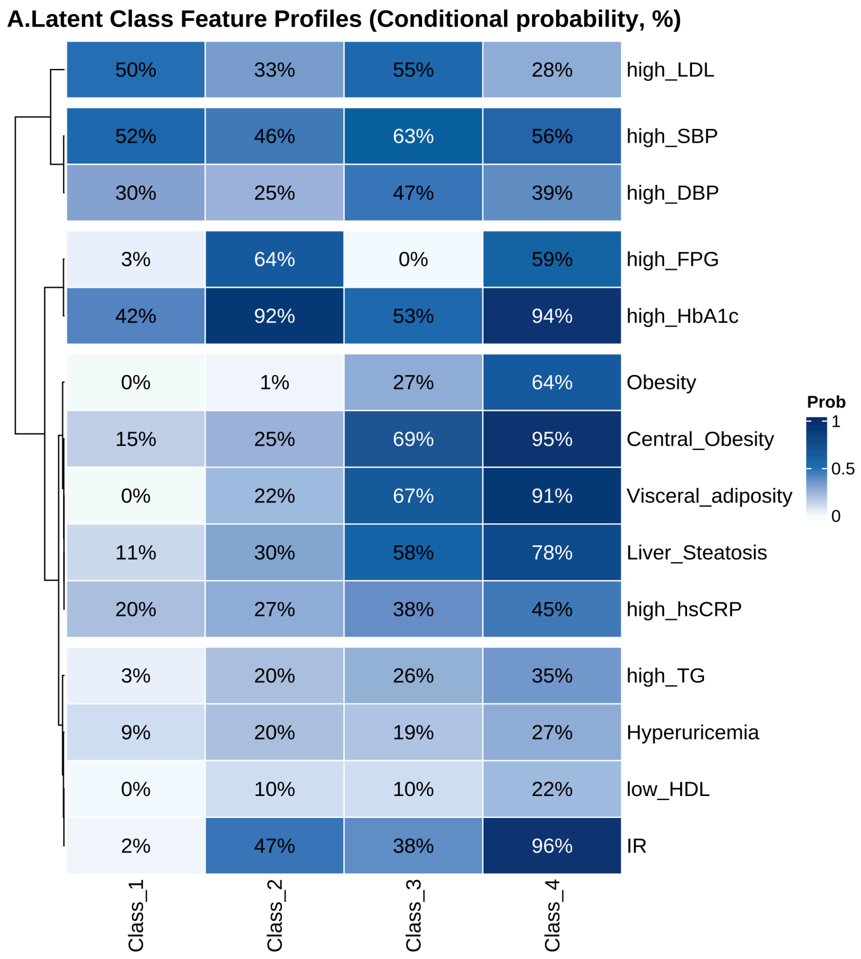


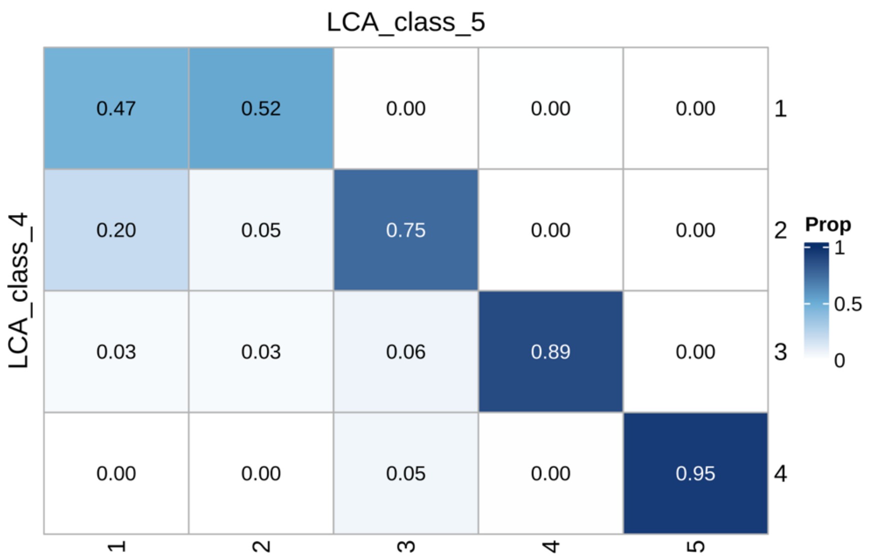


**Figure S8 Sensitivity analysis of choosing four latent classes.**
The sensitivity analysis of choosing four latent classes still shows subtypes including Class2: Lean-IR/Hyperglycemia and Class3: Obese-IS/Normoglycemia, demonstrating that the existence of adiposity and IR/Hyperglycemia discordance

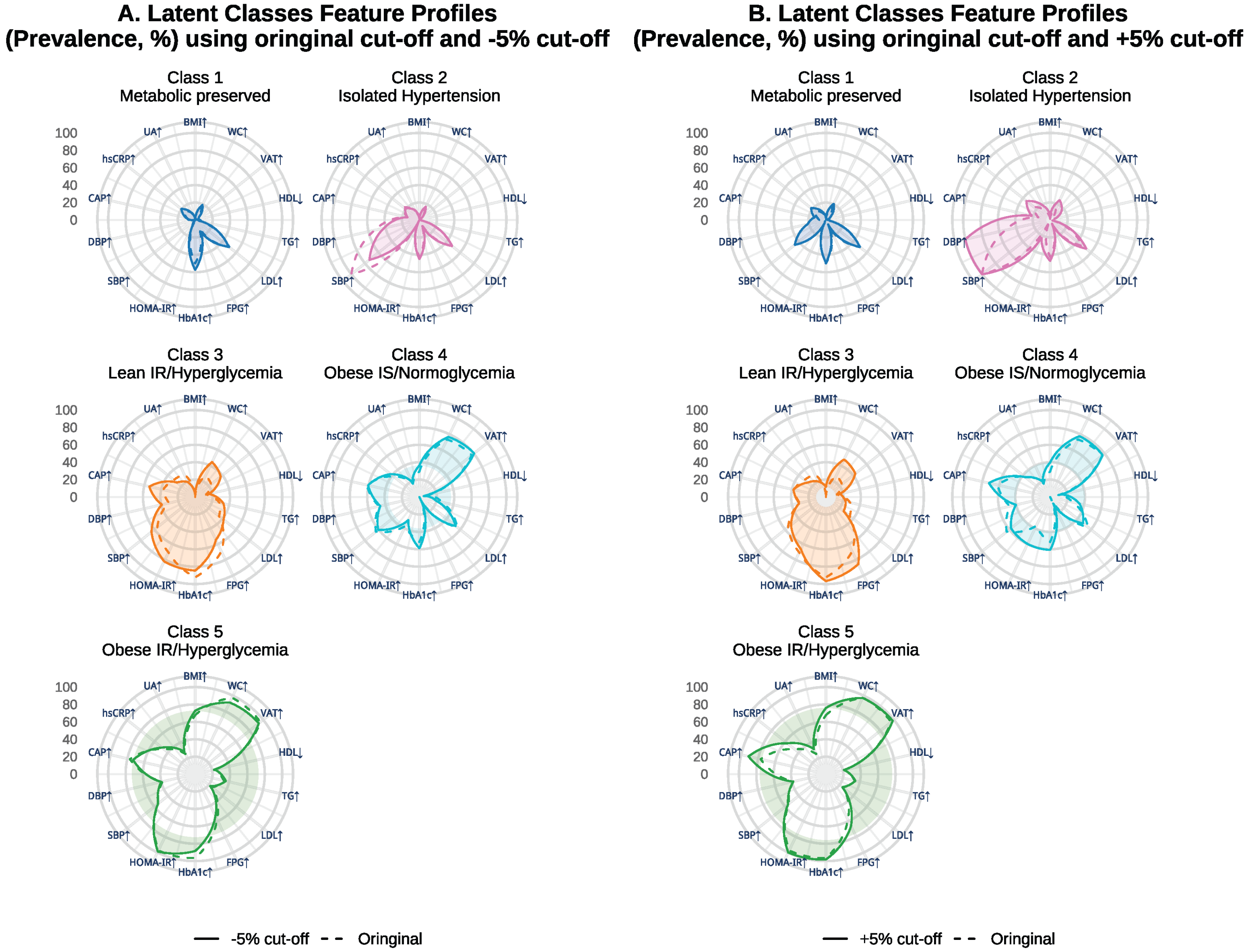


**Figure S9 Sensitivity analyses to vary each binary threshold by ±5% to assess the latent classification**

Radar plots illustrating the within-class prevalence of each metabolic phenotype among the five classes derived from the binary variables according to the original cut-off and ±5% cut-off.

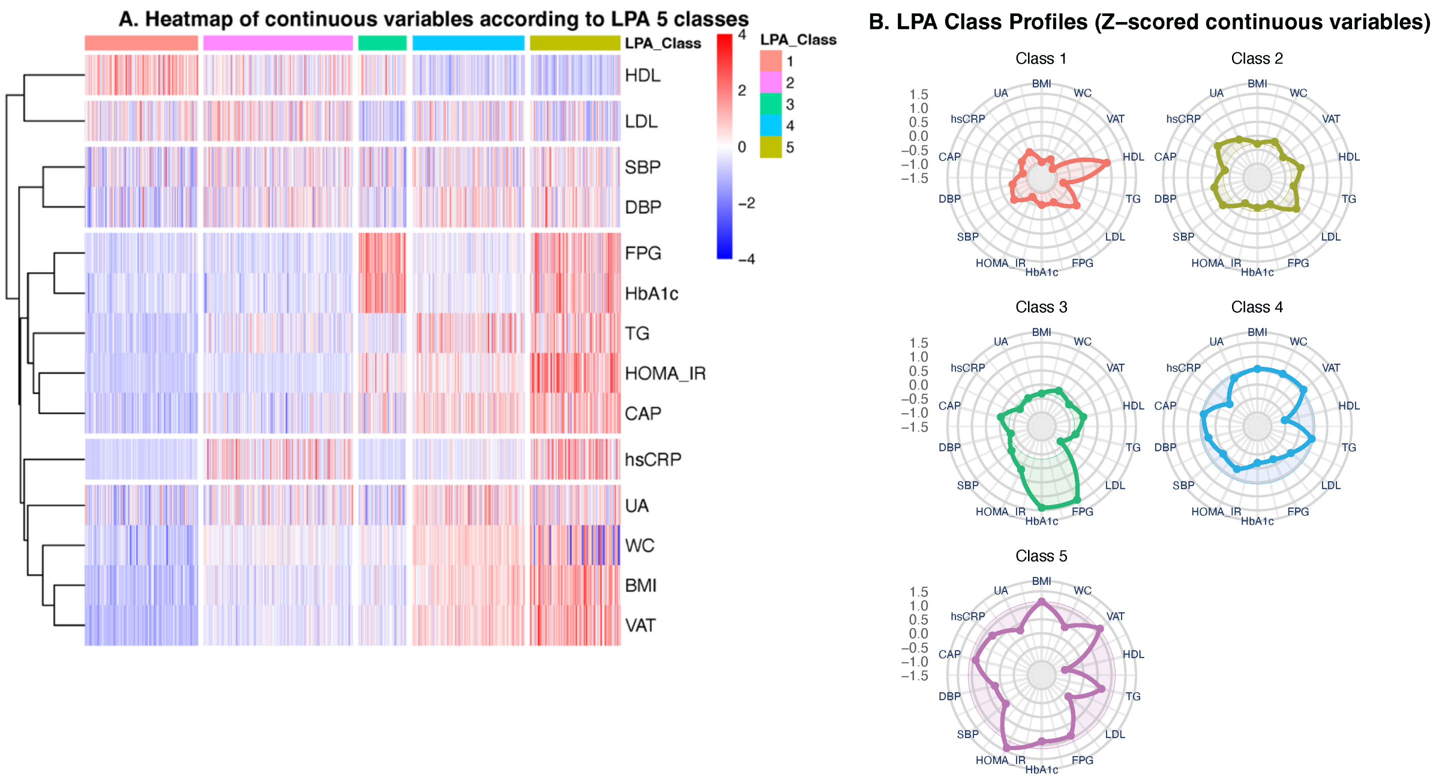


**Figure S10 Sensitivity analysis of Latent profile analysis (LPA) on continuous data**
Latent profile analysis (LPA) on continuous data was performed to identify whether distinct phenotypes would be obtained independent of binary diagnostic cut-offs. Different LPA models using equal, zero or varying variances and covariances were compared according to the lowest BIC. The best LPA model was set as "Varying variances, zero covariances". We manually choose 5 classes for LPA. The results showed that latent phenotype such as Class 3 and Class 4 still exists when using continuous variables, specifically, Class 3 represents lean but hyperglycemia and Class 4 represents obese but relatively normoglycemia. However the HOMA-IR did not show significant difference between these two classes.

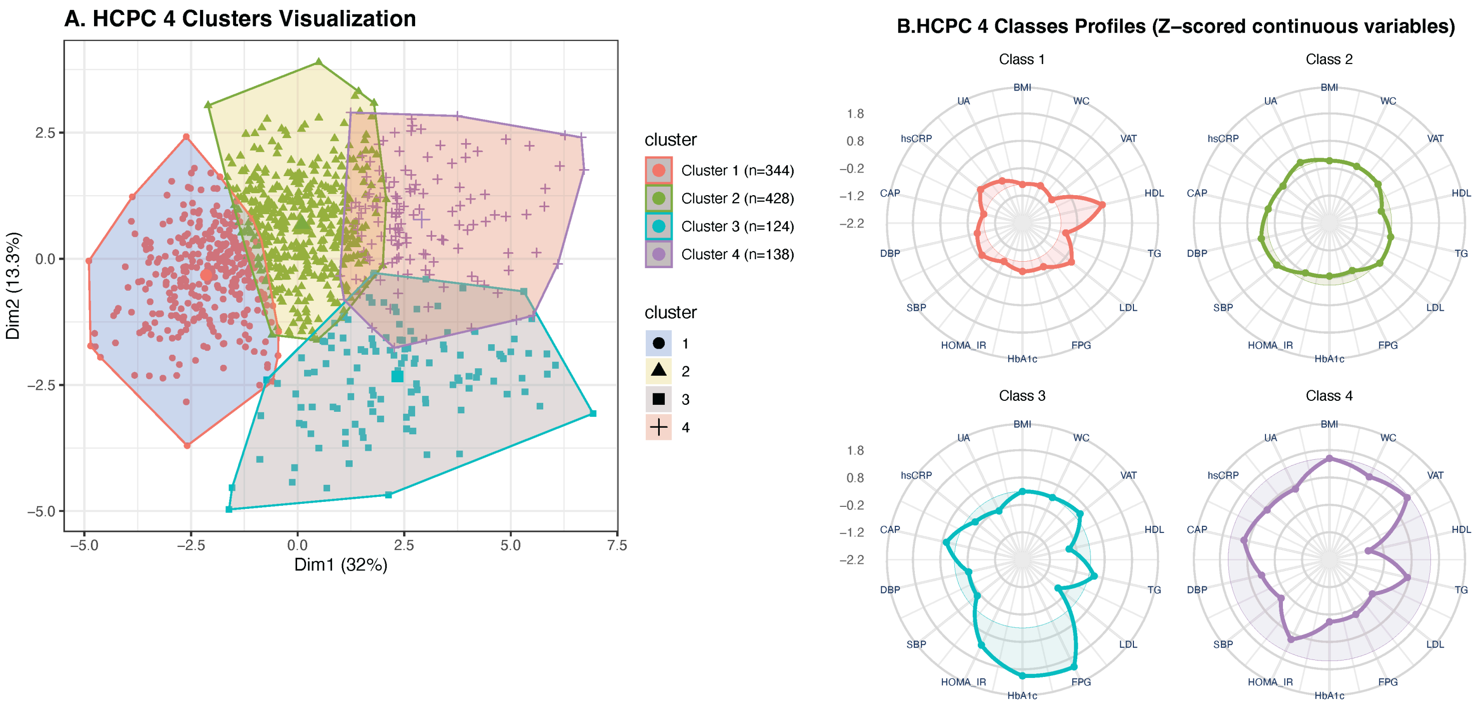


**Figure S11 Sensitivity analysis of Hierarchical Clustering on Principal Components (HCPC) on continuous data**

For continuous variables we performed unsupervised clustering using Hierarchical Clustering on Principal Components (HCPC) on continuous data to identify groupings independent of binary diagnostic cut-offs. All variables were Z-standardized, and Principal Component Analysis (PCA) was performed to reduce dimensionality, retaining the top ten principal components sufficient to explain ~90% of the variance. The final optimal cluster numbers: k=4 was derived from the hierarchical tree and consolidated via K-means to optimize stability. Analysis was performed using the FactoMineR package.

The results showed that latent phenotype such as Class 3 and Class 4 still exists when using continuous variables, specifically, Class 3 represents lean but hyperglycemia and Class 4 represents obese but relatively normoglycemia. However the HOMA-IR did not show significant difference between these two classes.

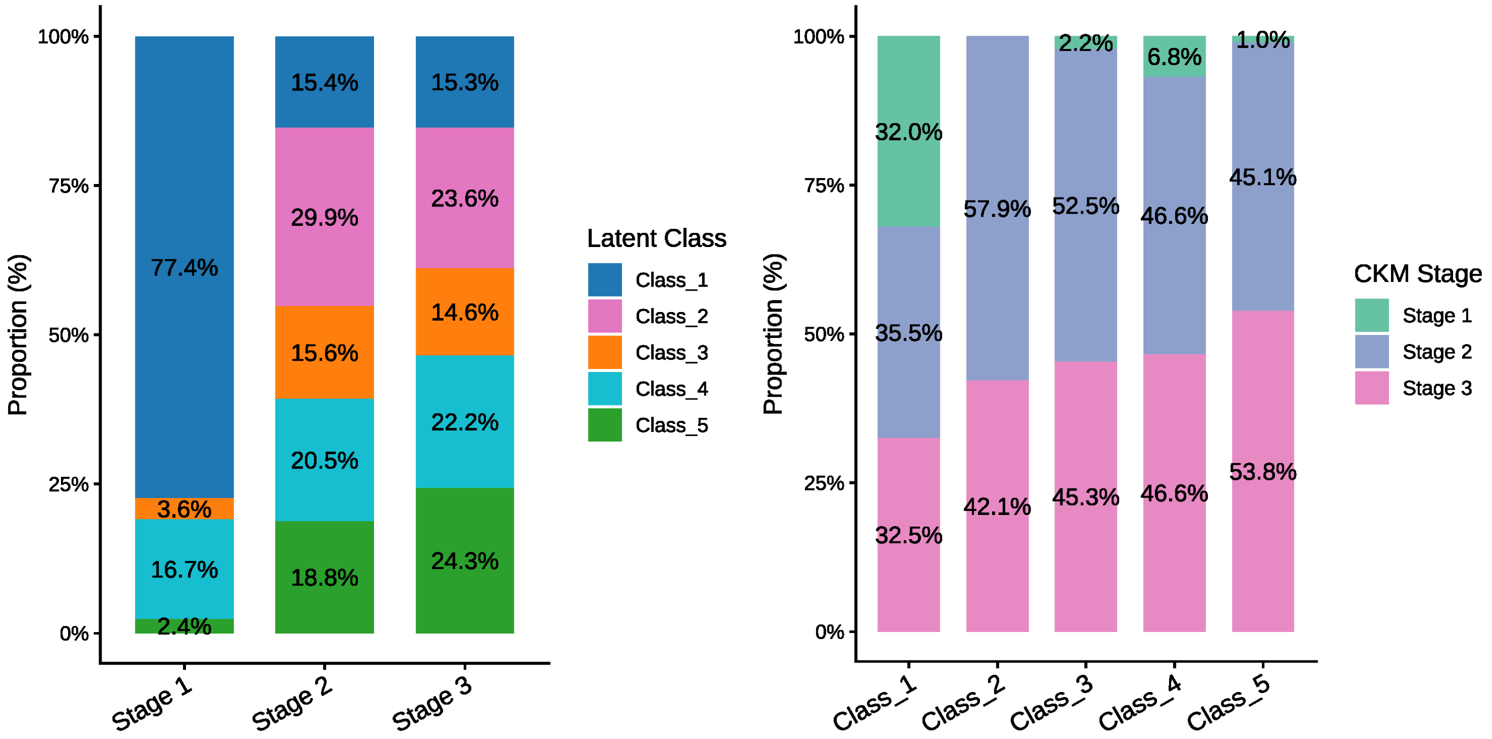


**Figure S12 Distribution of five latent classes within CKM stages**

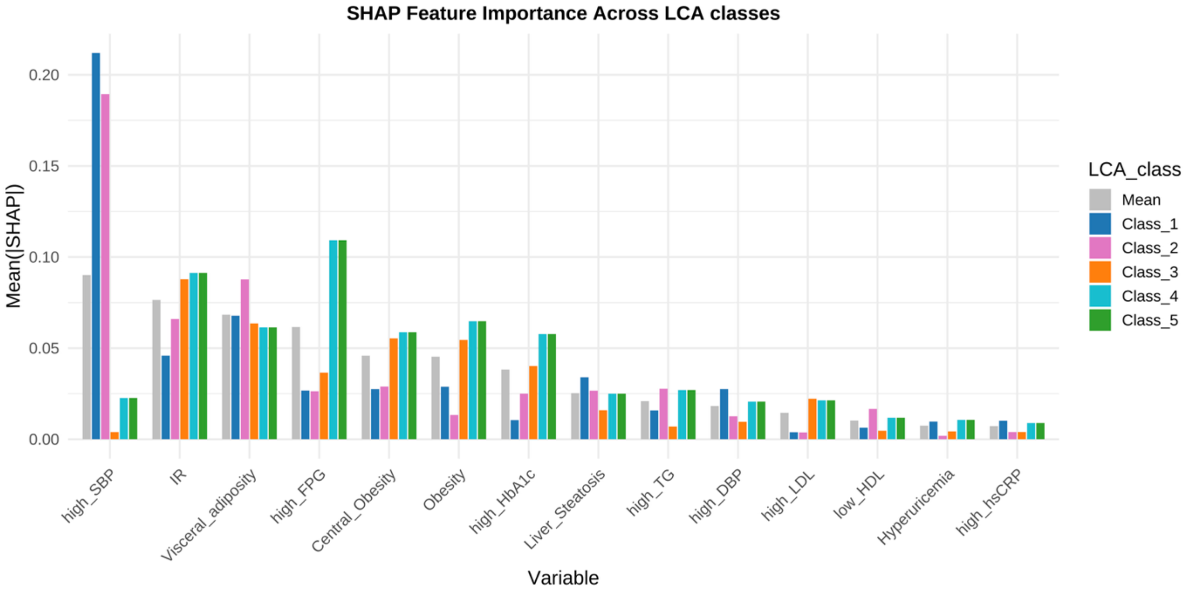


**Figure S13 SHAP identifying the importance of features in a multinomial logistic regression model for predicting five latent classes in discovery cohort–RESET study**

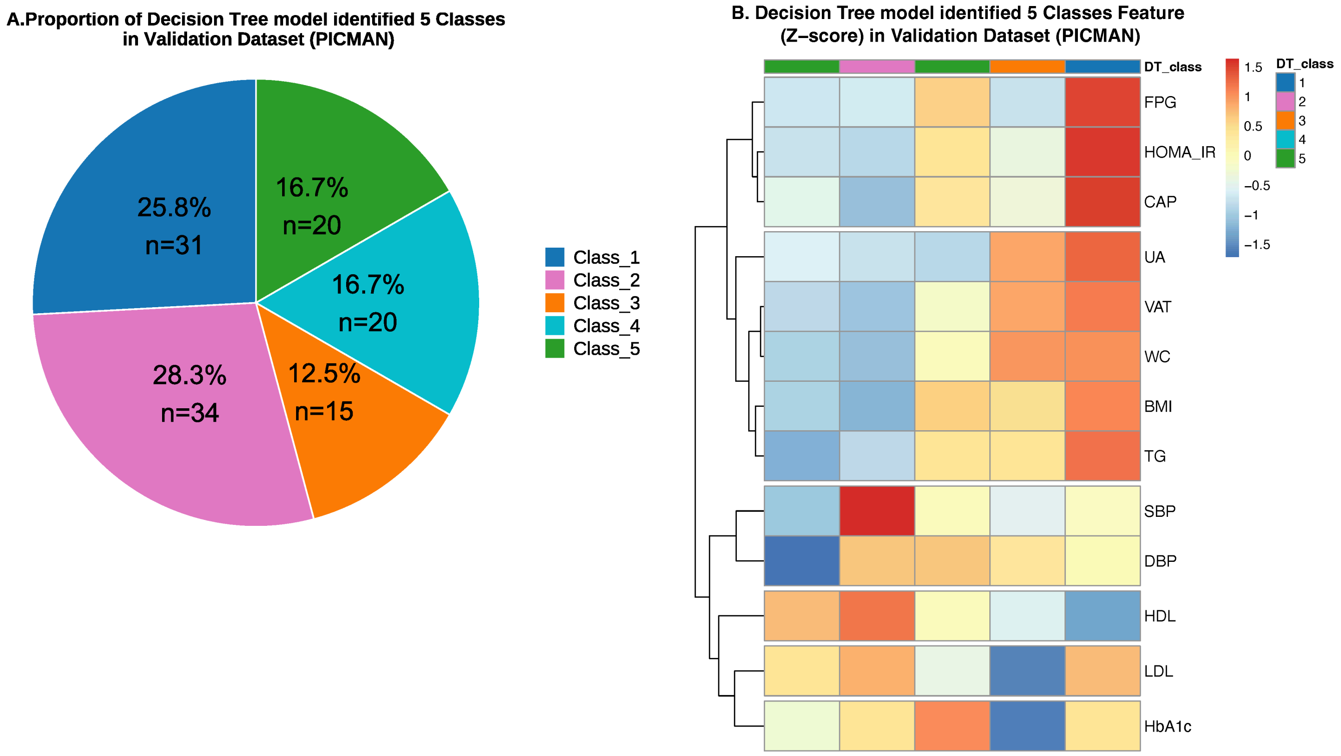


**Figure S14 Validation of decision tree model identified five classes in PICMAN study**

A. Pie plot showing the prevalence of five classes identified by decision tree model in PICMAN (N=120). In PICMAN, the insulin resistance was defined as HOMA-IR>66.6th percentile of the distribution, and the visceral adiposity was defined as visceral adipose tissue (VAT) volume by MRI> 66.6th percentile of the distribution.

B. Comparison of the median expression value of metabolic traits among across the five latent classes in PICMAN.

IR, insulin resistance; LDL, Low density lipoprotein cholesterol; HDL, High density lipoprotein cholesterol; SBP, systolic blood pressure; DBP, diastolic blood pressure; FPG, Fasting plasma glucose; HbA1c, Glycated Hemoglobin A1c; TG, triglyceride; HOMA-IR, Homeostatic Model Assessment of Insulin Resistance; CAP, Controlled attenuation parameter (Db/m); hsCRP, High Sensitivity C-reactive Protein; UA, Uric acid; WC, Waist circumference; BMI, Body mass index; VAT, Visceral adiposity tissue area by MRI

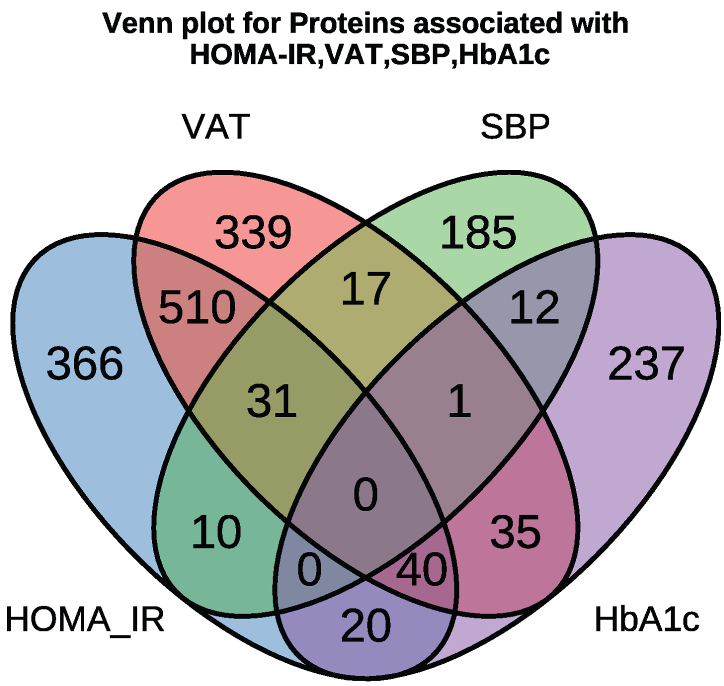


**Figure S15 Venn plot for proteins associated with HOMA-IR, VAT, SBP and HbA1c in the validation dataset (PICMAN)**

In PICMAN, the visceral adipose tissue (VAT) volume was determined by MRI.

Limma test was used to select the proteins significantly correlated to 4 continuous variables: HOMA-IR, VAT, SBP and HbA1c ($P<0.05$). Proteins significantly correlated to these 4 continuous variables were used for next step pairwise differential analysis among 5 latent classes identified by decision tree model

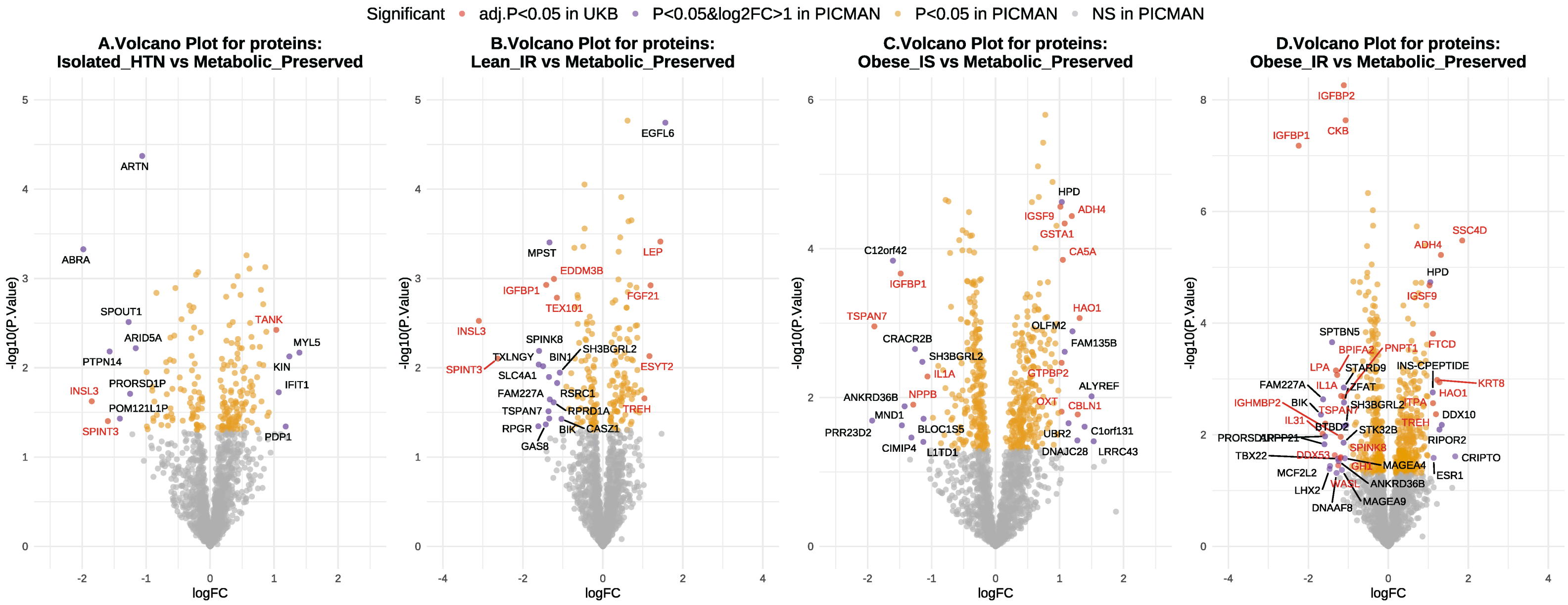


**Figure S16 Proteomic signatures of the 5 classes in the PICMAN study and the UK Biobank.**
Volcano plots showing differentially expressed proteins (DEPs) for: A. Metabolically isolated hypertension (Class2), B. Lean IR/hyperglycemia (Class3), C. Obese IS/normoglycemia (Class4), and D. Obese IR/hyperglycemia (Class5), each compared to the Metabolically preserved Class1. Proteins associated with HOMA-IR, VAT by MRI, SBP and HbA1c as continuous traits were first identified and subsequently subjected to pairwise differential expression analysis across latent classes. Proteins with $P < 0.05$ and log 2 fold change (FC)>1 in PICMAN were labelled on the Volcano plot, and proteins also with FDR-adjusted $P < 0.05$ in UK Biobank were labelled with red.

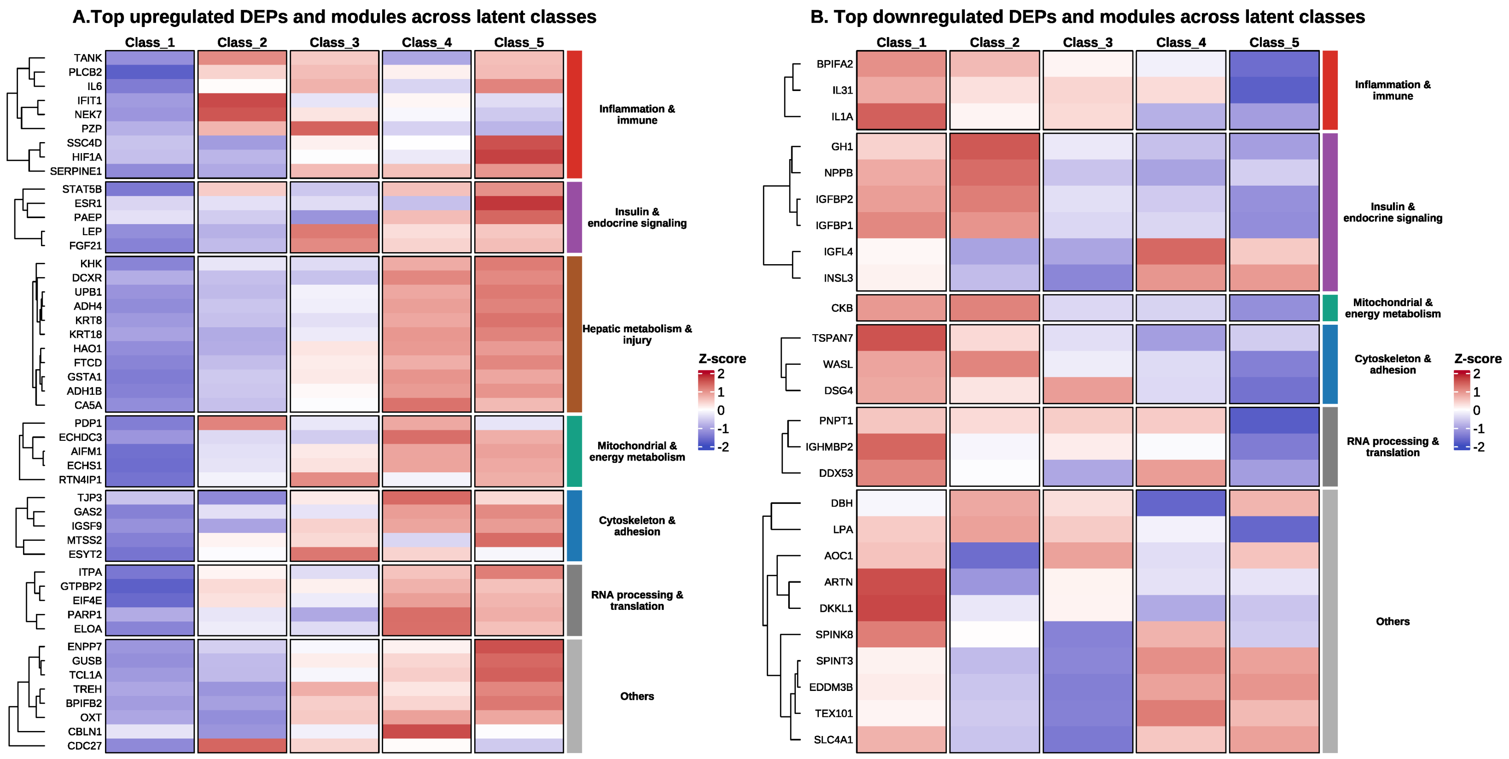


**Figure S17 Top differential proteomic signatures of the 5 classes in the PICMAN study and the UK Biobank.**
Proteins meeting significance thresholds in PICMAN ($P<0.05$ and $|\log_2$ fold change$| >0.8$) and replicated in the UK Biobank (FDR-adjusted $P<0.05$) are labelled.

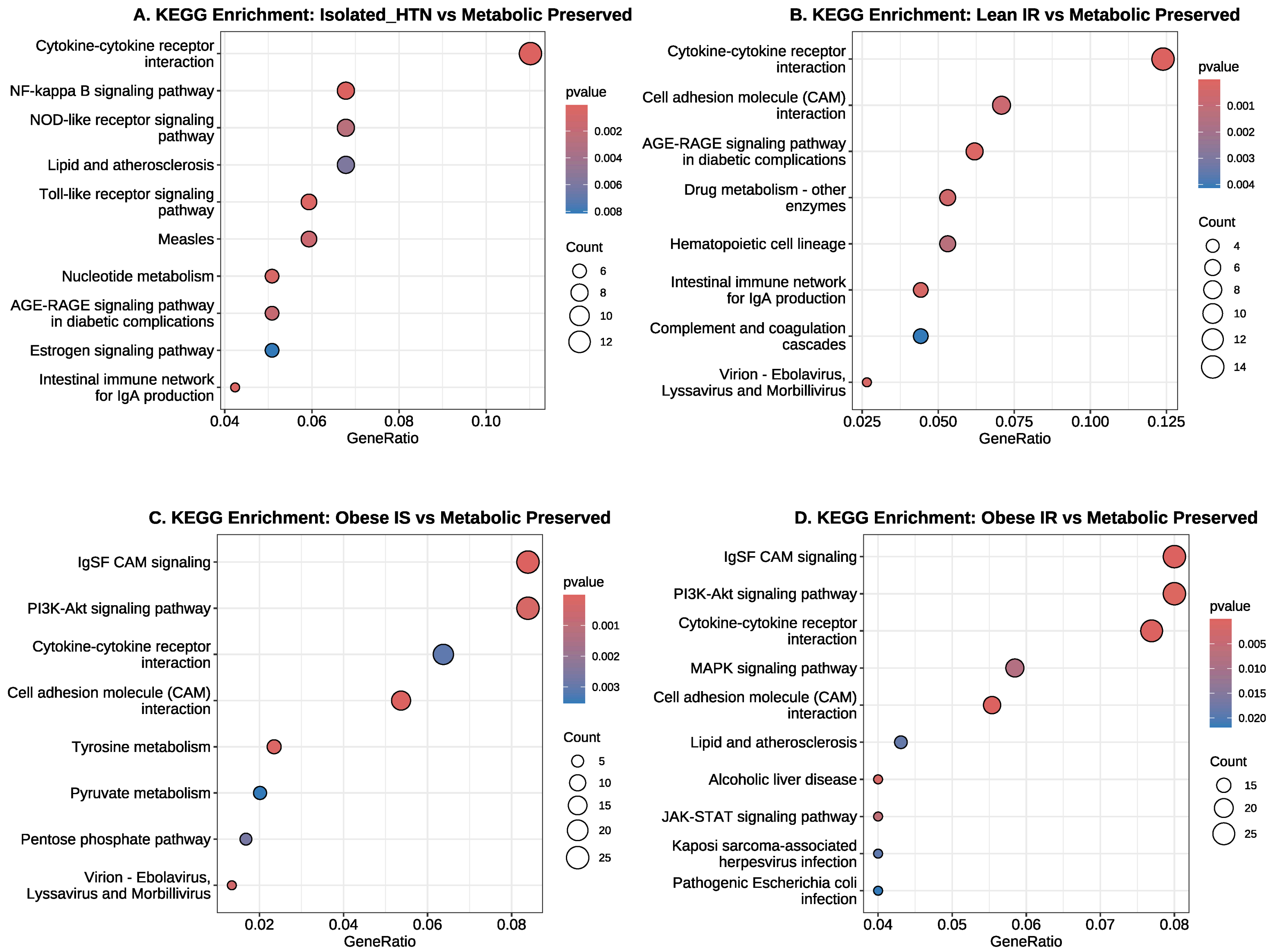


**Figure S18 KEGG over-representation analysis of differentially expressed proteins corresponding to each class compared with Metabolically Preserved (Class1).**

Dot plots showing KEGG pathways enriched by differentially expressed proteins (DEPs) for: A. Isolated Hypertension (Class2), B. Lean IR/hyperglycemia (Class3), C. Obese IS/normoglycemia (Class4), and D. Obese IR/hyperglycemia (Class5), each compared with the Metabolic Preserved group (Class1). Top 10 pathways with highest GeneRatio were plotted.

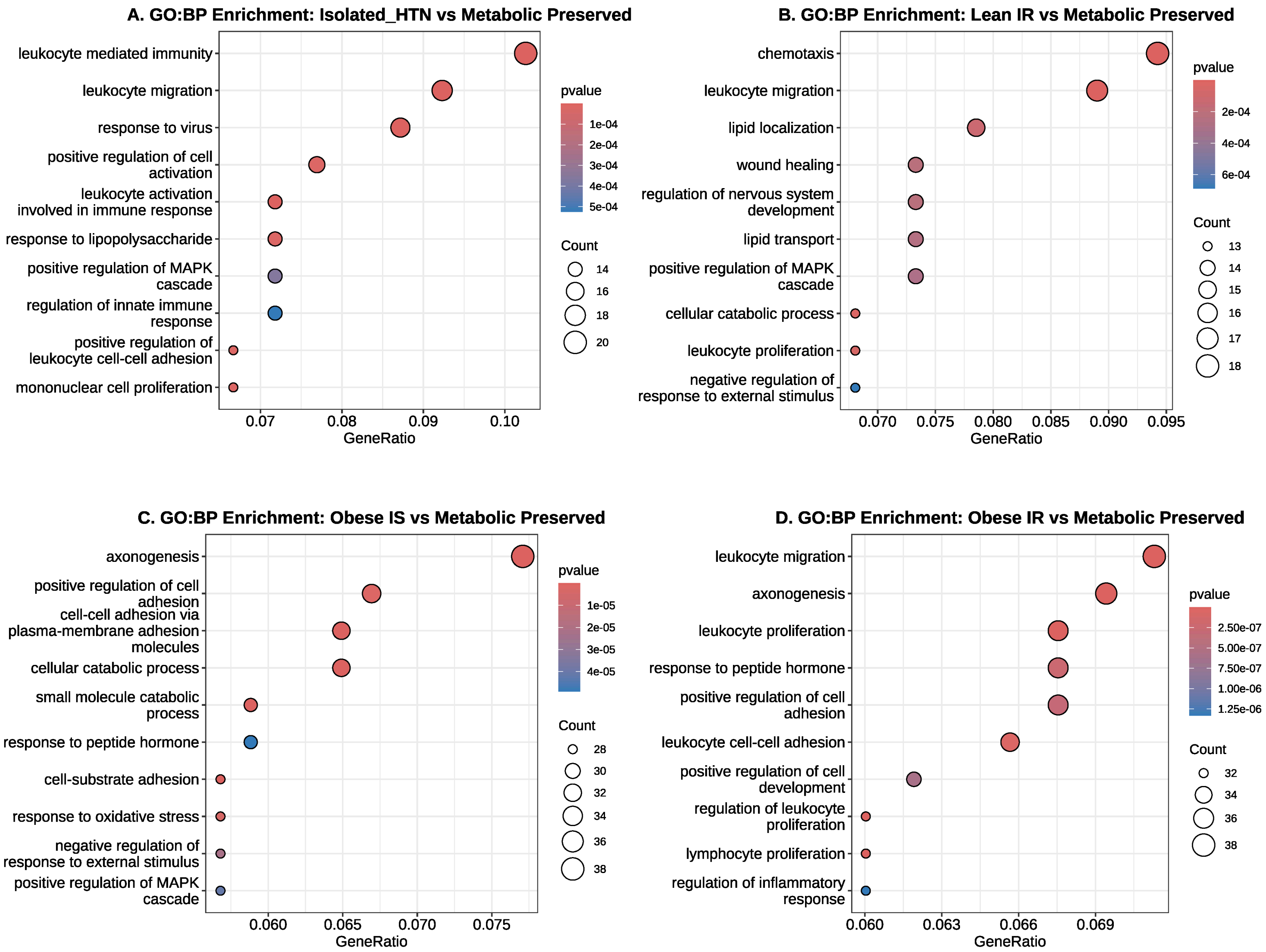


**Figure S19 Gene Ontology Biological Pathway (GO:BP) over-representation analysis of differentially expressed proteins corresponding to each class compared with Metabolically Preserved (Class1).**

Dot plots showing GO:BP pathways enriched by differentially expressed proteins (DEPs) for: A. Isolated Hypertension (Class2), B. Lean IR/hyperglycemia (Class3), C. Obese IS/normoglycemia (Class4), and D. Obese IR/hyperglycemia (Class5), each compared with the Metabolic Preserved group (Class1). Top 10 pathways with highest GeneRatio were plotted.

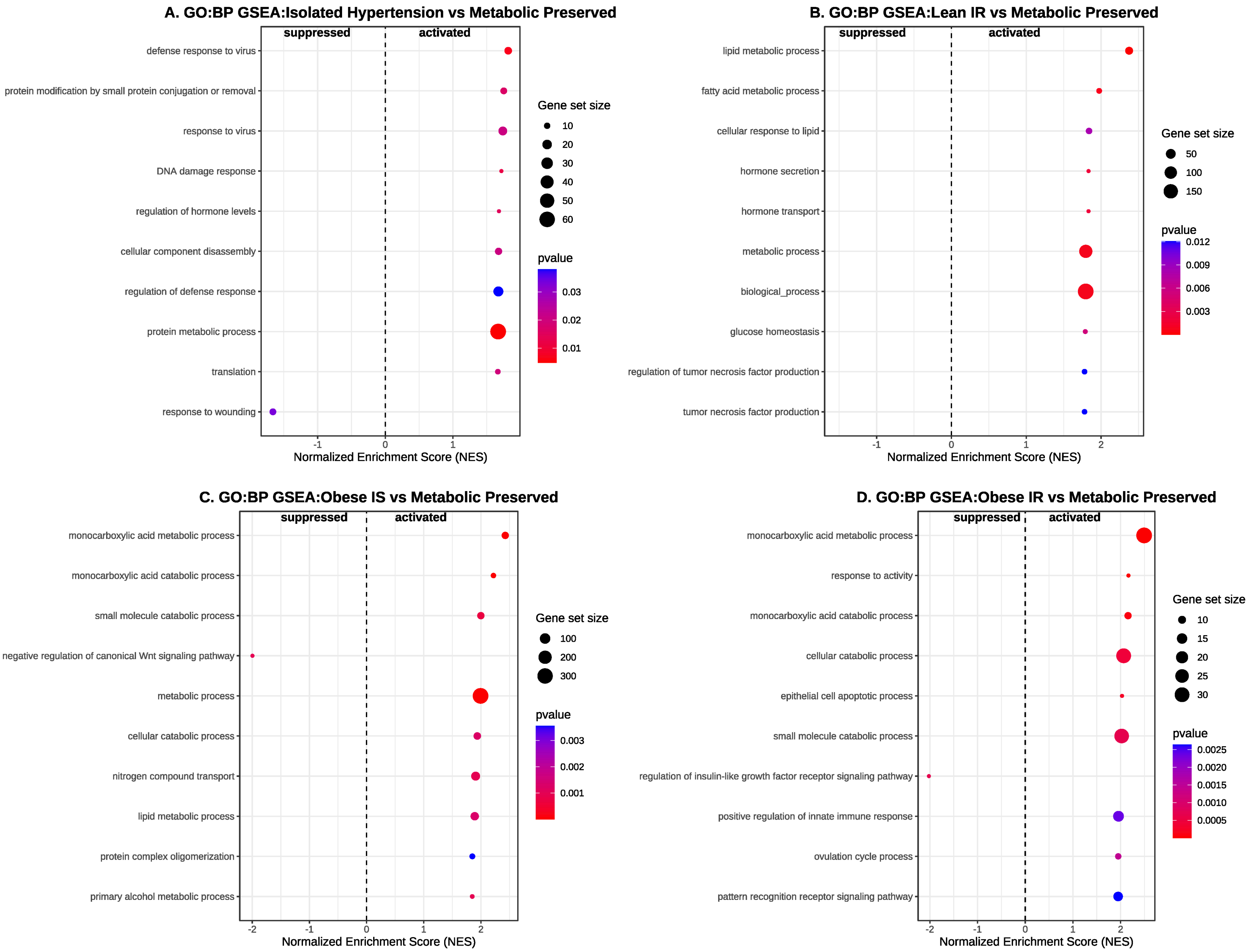


**Figure S20 Gene Set Enrichment Analysis (GSEA) of Gene Ontology Biological Pathway (GO:BP) among five latent classes in the validation dataset (PICMAN)**

Gene Set Enrichment Analysis (GSEA) of Gene Ontology Biological Pathway (GO:BP) for the differential proteins between: A. Isolated Hypertension versus Metabolic Preserved; B. Lean IR versus Metabolic Preserved; C. Obese IS versus Metabolic Preserved; D. Obese IR versus Metabolic Preserved. Top 10 GO:BP terms for activated or suppressed pathways were plotted descending by Normalized Enrichment Score (NES)

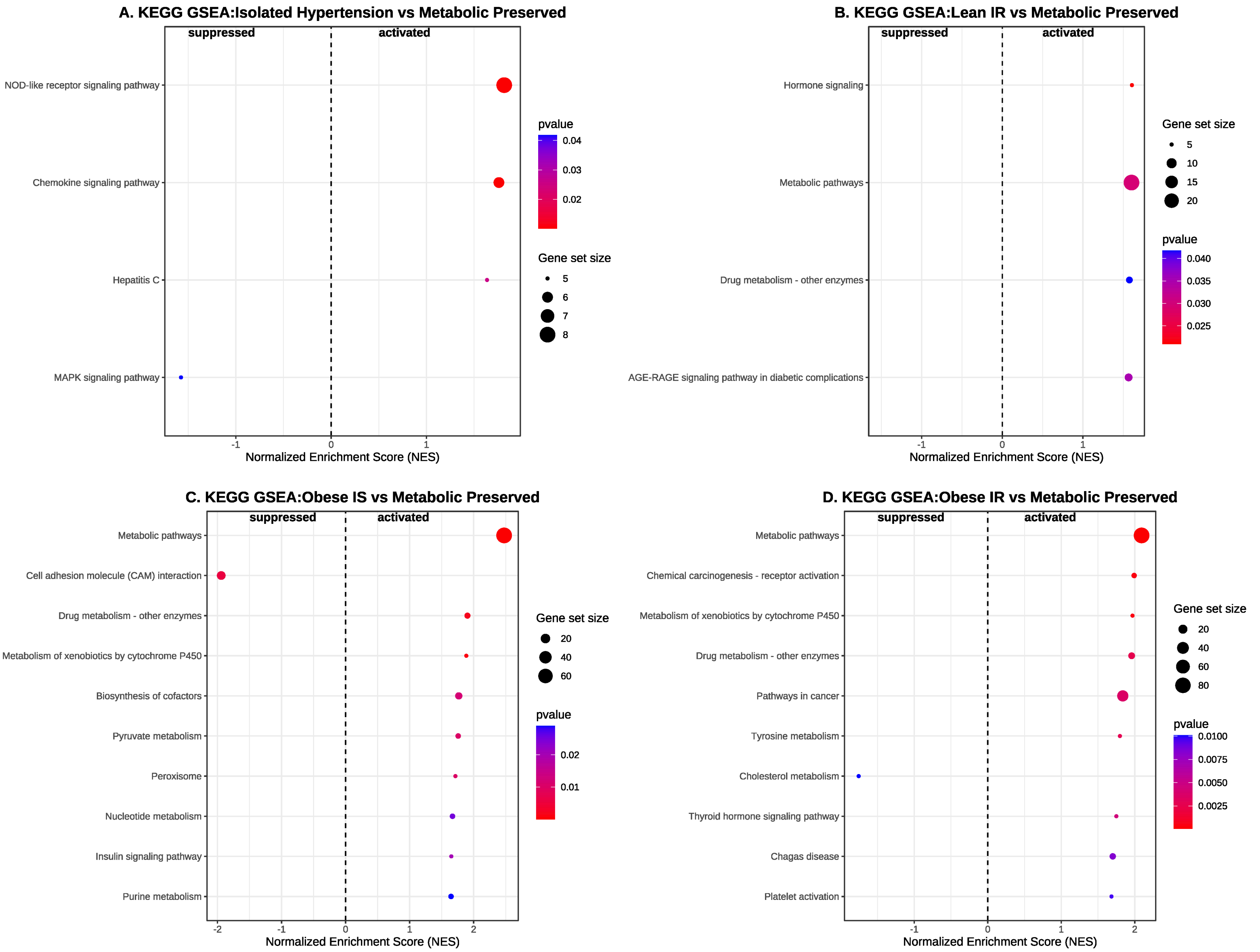


**Figure S21 Gene Set Enrichment Analysis (GSEA) of KEGG pathways among five latent classes in the validation dataset (PICMAN)**

Gene Set Enrichment Analysis (GSEA) of KEGG for the differential proteins between: A. Isolated Hypertension versus Metabolic Preserved; B. Lean IR versus Metabolic Preserved; C. Obese IS versus Metabolic Preserved; D. Obese IR versus Metabolic Preserved. Top 10 GO:BP terms for activated or suppressed pathways were plotted descending by Normalized Enrichment Score (NES)

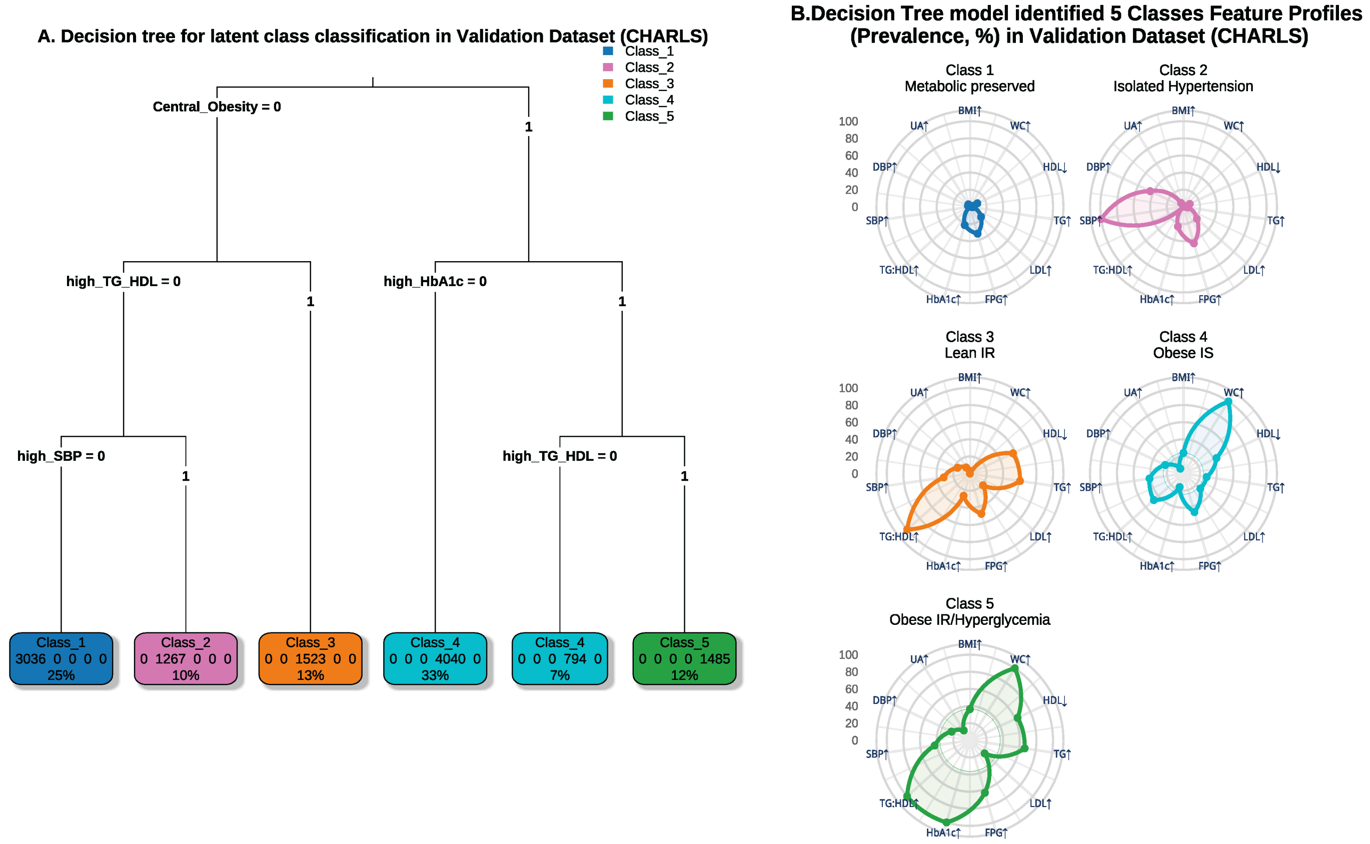


**Figure S22 Validation of decision tree identified latent phenotypes in CHARLS**

A. Tree graph showing the prevalence of five classes identified by decision tree model in The China Health and Retirement Longitudinal Study (CHARLS, N=12145). In CHARLS, the insulin resistance was replaced by the surrogate indicator high TG:HDL ratio (≥1.51 mmol/L in men and 0.84 mmol/L in women), and the visceral adiposity was replaced by the surrogate indicator central obesity (waist circumference≥80cm for female and ≥90cm for male).

B. Radar plots illustrating the within-class prevalence of each metabolic phenotype among the five classes derived from decision tree model in CHARLS.

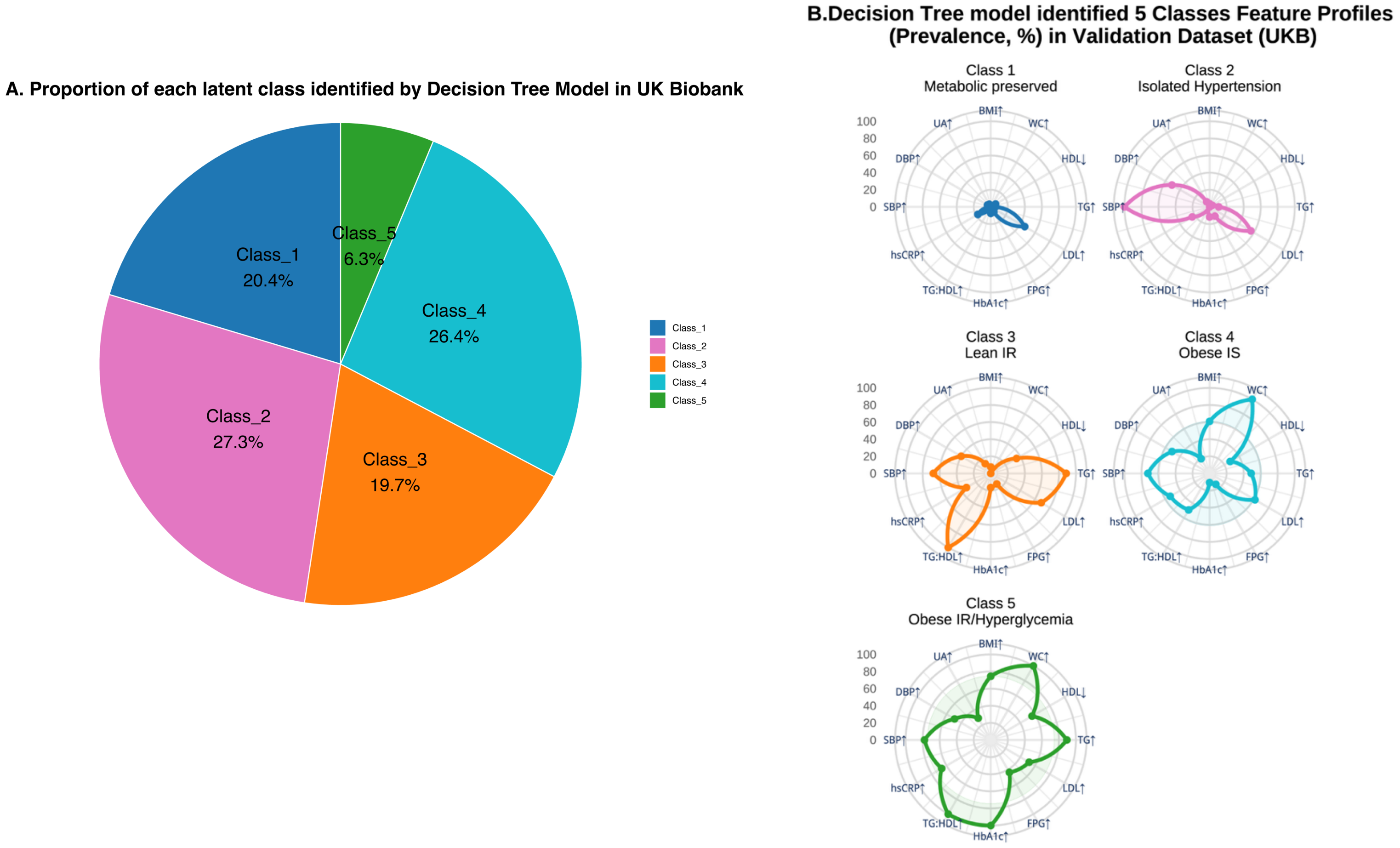


**Figure S23 Validation of decision tree identified latent phenotypes in UK Biobank**

A. Pie graph showing the prevalence of five classes identified by decision tree model in UK Biobank (N= 344817). In UK Biobank, the IR was replaced by the surrogate indicator high TG:HDL ratio (≥3.5 mmol/L in male and 2.5 mmol/L in female), and the visceral adiposity was replaced by the surrogate indicator central obesity (WC≥88cm for female and ≥102cm for male)

B. Radar plots illustrating the within-class prevalence of each metabolic phenotype among the five classes derived from decision tree model in UK Biobank.
